# The association between the gut microbiome and 24-hour blood pressure measurements in the SCAPIS study

**DOI:** 10.1101/2023.12.08.23299598

**Authors:** Yi-Ting Lin, Sergi Sayols-Baixeras, Gabriel Baldanzi, Koen F. Dekkers, Ulf Hammar, Diem Nguyen, Nynne Nielsen, Aron C. Eklund, Georgios Varotsis, Jacob B. Holm, H. Bjørn Nielsen, Lars Lind, Göran Bergström, J. Gustav Smith, Gunnar Engström, Johan Ärnlöv, Johan Sundström, Marju Orho-Melander, Tove Fall

## Abstract

**Background and Aims:** Large population-based studies on gut microbiota and hypertension have been conducted using methods with low taxonomic resolution and office blood pressure. This study aims to investigate the relationship between specific characteristics of the gut microbiome and 24-hour blood pressure measurements.

**Methods and results:** The association of gut microbial species, determined by shotgun metagenomic sequencing of fecal samples, with 24-hour ambulatory blood pressure measurements was assessed in 4063 participants without antihypertensive medication from the Swedish CArdioPulmonary bioImage Study. Multivariable-adjusted models identified 140 microbial species associated with at least one of the 24-hour blood pressure traits. Notably, *Roseburia faecis*, *R. inulinivorans*, and *Dorea longicatena* were strongly positively associated with mean systolic and diastolic blood pressure, while *Alistipes communis* and *A. shahii* were inversely associated with diastolic blood pressure. An enrichment of threonine degradation I, *Bifidobacterium* shunt, and lactate production was observed in species associated with mean systolic and diastolic blood pressure. Species positively associated with blood pressure were in general also positively associated with secondary bile acids but negatively associated with primary bile acids and vitamin A-related metabolites.

**Conclusions:** In this large cross-sectional analysis, we identified a group of gut microbial species and microbial functions associated with blood pressure. Our findings provide insights into the relation of the gut microbiome and blood pressure, which can lead to a new understanding of the etiology of hypertension.

## Introduction

Hypertension is estimated to have contributed to 10.8 million deaths globally in 2019 through its impact on cardiovascular and renal disease development.^1^ Yet, our understanding of hypertension pathophysiology is incomplete, although it has been hypothesized that the gut microbiota affects the blood pressure (BP) regulation mediated through microbiota-derived metabolites and interactions with the immune system.^2^ Evidence supporting the role of the microbiota in hypertension is based on experimental and observational studies. Germ-free mice transplanted with fecal material from hypertensive human donors showed higher systolic (SBP) and diastolic BP (DBP) than mice transplanted with material from a normotensive donor.^3^ Plasma metabolites derived or modified by the gut microbiota, including trimethylamine N-oxide (TMAO) and short-chain fatty acids (SCFA), have been suggested to affect blood pressure through chemosensory receptors.^4–6^

In 4672 participants from the HEalthy Life In an Urban Setting (HELIUS) study, where the gut microbiota was characterized using the 16S amplicon method, inverse associations of SCFA-producing bacteria with SBP were observed.^7^ However, findings were inconsistent across ethnicities. Another large study based on shallow metagenomics sequencing in samples from 6953 participants of the FINRISK 2002 study identified mainly positive associations between 45 microbial genera and blood pressure. Studies applying methods resulting in a higher resolution of the microbiome are anticipated to provide more specific results.

Moreover, while SBP and DBP measured at the test centers constitute a snapshot of BP, ambulatory blood pressure monitoring (ABPM) provides a more representative measurement of SBP and DBP. ABPM is not affected by “white coat hypertension” and it captures nocturnal BP. ABPM can also be used to estimate the variability of BP, often defined as the standard deviation (SD) of SBP and DBP, and has been reported as an independent risk factor for cardiovascular disease.^8^ However, there is a lack of population-based studies investigating the association between the gut microbiota and BP using high-resolution methods to assess the microbiota composition and functional profile, as well as ABPM for capturing BP and BP variation.

Therefore, we aimed to investigate the association of the gut microbiome composition measured at high resolution and functional profile with ABPM measurements, as well as the association of plasma metabolites with species related to ABPM measurements, in a large sample of participants without antihypertensive medication.

## Methods

### Study population

The Swedish CArdioPulmonary bioImage Study (SCAPIS) cohort is a prospective observational study with the baseline investigation conducted in 2013-2018, including 30 154 individuals invited from a random extract from the population register of individuals aged 50–64 living in six counties across Sweden including 2-3 visits to the respective study centers.^9^ We used data from the baseline investigation from the Uppsala (n=5036) and Malmö (n=6251) centers, of which 4839 individuals in Uppsala and 4980 in Malmö had available fecal metagenomics data, and where a subset were provided with equipment for ABPM (**Supplementary Figure S1**). We excluded 2459 participants who had a prescription for antihypertensive medication in the last 12 months in the Swedish Prescribed Drug Register (ATC codes in **Supplementary methods**) before the first baseline visit and 30 participants with missing information in the country of birth. The remaining 7330 participants were split into two groups. The “ABPM subsample” included those 4063 participants (3261 from Uppsala, 802 from Malmö) who had 24-hour BP records that passed quality control.^10^ There were 312 individuals whose ABPM data did not pass the quality control. After removing 6 participants without any office BP measurement, the final “non-ABPM subsample”, consisted of 395 participants from Uppsala and 2866 from Malmö who had valid office BP measurements. All participants provided written informed consent. The study was conducted in accordance with the Declaration of Helsinki. The Swedish Ethical Review Authority approved the Swedish CardioPulmonary bioImage Study (DNR 2010-228-31M) and the present study (DNR 2018-315).

### Blood pressure measurements

Participants of the ABPM subsample were instructed to apply the ABPM device (Labtech EC-3H/ABP, Labtech Ltd, Debrecen, Hungary) in the morning and to remove it 24 hours later. SBP and DBP were measured automatically every 30 minutes for participants from the Malmö center. SBP and DPB were measured every 30 minutes during the day, and every 90 minutes during the night in participants from the Uppsala center (**Supplementary methods**).^10^ Office BP was measured after five minutes of rest in supine position using an automatic device at the brachial artery (Omron M10-IT, Omron Healthcare Co. Ltd., Kyoto, Japan).

### Microbiome analysis

Fecal samples were collected for microbiome analysis, as previously described.^11^ At the first visit, participants received a fecal sample collection kit with barcoded tubes, gloves, re-sealable plastic bags, paper collection bowls, and instructions from the SCAPIS test center. Participants were instructed to collect fecal samples at home and to store the tubes in sealed plastic bags in their home freezer until the second visit to the test center, where they were stored for a maximum of 7 days at -20°C until transferred to the -80°C freezers at the central biobank. The samples were shipped on dry ice to Clinical Microbiomics A/S (Copenhagen, Denmark), where DNA extraction, metagenomic shotgun sequencing, and bioinformatics analyses were performed.^11^ In brief, the process involved sequencing with Illumina Novaseq 6000 platform yielding on average 26 million read pairs per sample. Non-host reads and previously published data were utilized to construct nonredundant gene catalogues for fecal samples, where metagenomic species (MGS) and their corresponding signature gene sets were identified. Species abundance was based on the abundance of signature gene sets adjusting for effective gene length, and taxonomic annotation was based on the NCBI RefSeq database.^12^ The Shannon diversity index, an alpha diversity metric that provides information about richness and evenness in the gut microbiota,^13^ was estimated with R package *vegan* v2.5-7 using rarefied data. The functional potential profile of the gut microbiota was determined by assigning genes to 103 metabolic pathways comprising the gut metabolic modules (GMM).^14^ A species was considered to carry a GMM if it carried at least two-thirds of the KEGG (Kyoto Encyclopedia of Genes and Genomes) Orthology of a module. For modules with three or fewer steps, all steps were required. For modules with alternative paths, only one path had to fulfill the criterion.

### Metabolomics

The GUTSY Atlas (https://gutsyatlas.serve.scilifelab.se/)^11^ was used to investigate the associations between 24-hour BP-related species and plasma metabolites in SCAPIS. The methodological details are described in Dekkers et al.^11^

### Other phenotypes

Information on smoking, country of birth, previous diagnosis of diabetes mellitus and inflammatory bowel diseases (Crohn’s disease and ulcerative colitis), use of antidiabetic and antihyperlipidemic drugs, and diet were based on questionnaire information. Smoking was categorized as either current smoker or non-smoker.Country of birth was categorized as Scandinavia (Sweden, Norway, or Finland), non-Scandinavian Europe, Asia, and others. The MiniMeal-Q food frequency questionnaire^15^ was used to estimate total energy intake and energy-adjusted fiber intake. Energy-adjusted fiber intake was determined by calculating the amount of consumed fiber per 1000 kcal of energy. Diet data from women reporting energy intake values <500 or >5000 kcal/day, and men reporting <550 or >6000 kcal/day were excluded as they were likely misreported.

Urinary sodium (Architect c16000; Abbott Laboratories, Abbott Park, IL, USA) and creatinine (enzymatic method) were analyzed by Clinical Chemistry at Uppsala University Hospital, Uppsala, Sweden from the morning fasting spot urine samples collected at first visit.^16, 17^ As a surrogate for sodium intake, the 24-hour sodium excretion was estimated based on the spot urinary sodium using the Kawasaki formula.^18^

Information on antibiotic use was extracted from the Swedish Prescribed Drug Register 6 months preceding the visit 1. Proton pump inhibitor (PPI) use was defined as participants with measurable omeprazole and/or pantoprazole levels in plasma from metabolomics data.

### Statistical analysis

#### Main analysis

All the statistical methods were performed using R (version 4.1.1). Four different 24-hour BP outcomes were assessed: mean SBP and DBP and variability of SBP and DBP, measured as the SD of the respective trait. A series of linear regression models were applied in the ABPM subsample assessing the association of microbiome diversity (Shannon diversity index) and species with these four outcomes, one at a time. The species relative abundance was ln(x+1) transformed, where x denotes the species relative abundance, followed by a z-transformation to set the mean 0 and the SD to 1. A false discovery rate (FDR) of 5% using Benjamini-Hochberg method was applied based on *P*-values from all four phenotypes together.^19^

We first applied a model with adjustment for age, sex, country of birth, and technical source of variation stemming from DNA extraction plates where the samples from the two study centers were analyzed on different plates (Model 1). We assessed the enrichment for genera and GMMs using the gene set enrichment analysis method^20^ from the *fgsea* R package. For the enrichment analysis, the ranked *P*-values of the associations between species and 24-hour BP outcomes were stratified by effect direction from Model 1.

Associations with FDR <5% in Model 1 were selected for further assessment in Model 2. Model 2 was additionally adjusted for smoking, fiber intake, total energy intake, estimated sodium intake, use of antidiabetic drug, and use of antihyperlipidemic drug. We used a complete case approach, analyzing the 3694 individuals with complete data on these phenotypes with one sample excluded due to low read counts in the analyses using rarefied data. The selection of these covariates was made using d-separation criteria applied on a directed acyclic graph (DAG) assisted by the DAGitty, version 3.0, software (www.dagitty.net; **Supplementary Figure S2A**).^21^

We considered BMI as a potential mediator or confounder. Therefore, to identify species associated with BP independent of BMI, we also analyzed the associations in Model 3, which included additional adjustment for BMI (**Supplementary Figure S2B)**.

To ensure the associations were not driven by influential observations in Model 2 and Model 3, unscaled dfbeta values were calculated using dfbetas R function for each association. For an association to be considered reliable, the *P*-value after excluding the most influential value had to be <0.05 and the direction of the regression coefficient had to remain unchanged. To validate the findings and assess the generalizability of associated species, we repeated the analysis in Model 2 using office BP measurements in the ABPM and non-ABPM subsamples separately.

From the GUTSY Atlas, we retrieved the associations between 24-h BP-associated species in Model 2 and plasma metabolites. The three associations with lowest *P*-values between the species and known plasma metabolites were selected for visualization. Hierarchical clustering using the Euclidean distance on the partial Spearman correlation coefficients was performed on the unique subset of 50 selected metabolites for the mean 24-hour BP and 32 metabolites for variability of 24-hour BP.

#### Sensitivity analysis

The following sensitivity analyses were performed in Model 2: (1) exclusion of participants with a dispensed antibiotic prescription within 6 months before the visit 1; (2) exclusion of participants with inflammatory bowel diseases; and (3) adjustment for PPI usage.

## Results

### Baseline characteristics

The ABPM subsample included 3261 participants recruited in Uppsala (mean age 57.3 years, mean 24-hour SBP/DBP 122/76 mmHg) and 802 participants recruited in Malmö (mean age 57.0 years, mean 24-hour SBP/DBP 122/75 mmHg) (**Table 1**). The non-ABPM subsample consisted of 395 participants from Uppsala (mean age 57.3, office SBP/DBP, 124/77) and 2866 participants from Malmö (mean age 56.9 years, office SBP/DBP, 120/74 mmHg) (**Table 1**). Self-reported comorbidities were similar in Uppsala and Malmö, but a higher proportion of current smokers was noted in Malmö (14.5% in ABPM and 18.9% in non-ABPM) than in Uppsala (8.5% in ABPM and 16.4% in non-ABPM).

**Table 1.** Descriptive characteristics of participants in the Swedish CArdioPulmonary bioImage Study (SCAPIS) study in the Uppsala and Malmö centers. Participants had metagenomics data and were not taking blood pressure medication. Blood pressure measurements were conducted in two subsamples, the ABPM and non-ABPM subsamples.

|  | **ABPM subsample** |  | **non-ABPM subsample** |  |
| --- | --- | --- | --- | --- |
|  | **Uppsala (n=3261)** | **Malmö (n=802)** | **Uppsala (n=395)** | **Malmö (n=2866)** |
| Age, years | 57.3 (4.37) | 57.0 (4.31) | 57.3 (4.45) | 56.9 (4.21) |
| Female, n (%) | 1735 (53.2) | 424 (52.9) | 195 (49.4) | 1564 (54.6) |
| Country of birth, n (%) |  |  |  |  |
| Scandinavia | 2915 (89.4) | 599 (74.7) | 354 (89.6) | 2276 (79.4) |
| Europe | 136 (4.17) | 137 (17.1) | 15 (3.8) | 367 (12.8) |
| Asia | 135 (4.14) | 47 (5.86) | 18 (4.56) | 154 (5.37) |
| Others | 75 (2.30) | 19 (2.37) | 8 (2.03) | 69 (2.41) |
| Current smoker, n (%) | 272 (8.5) | 116 (14.5) | 44 (16.4) | 526 (18.9) |
| Body mass index, kg/m<sup>2</sup> | 26.4 (4.11) | 26.8 (4.37) | 27.1 (4.14) | 26.7 (4.37) |
| Fiber intake, g/1000kcal | 18.6 (13.2–25.5) | 17.9 (12.5–25.7) | 19.8 (13.5–26.7) | 18.1 (12.5–25.2) |
|  |  | 1613 (1236– | 1713 (1296– |  |
| Total energy intake, kcal/day | 1631 (1292–2053) | 2123) | 2161) | 1604 (1251–2097) |
| Diabetes mellitus, n (%) | 65 (2.00) | 13 (1.62) | 6 (2.80) | 67 (2.48) |
| Medication for diabetes, n (%) | 52 (1.60) | 12 (1.50) | 3 (1.40) | 49 (1.81) |
| Antihyperlipidemic medication, n (%) | 93 (2.86) | 22 (2.75) | 8 (3.74) | 94 (3.48) |
| Proton pump inhibitor, n (%) | 64 (1.98) | 25 (3.42) | 6 (1.53) | 72 (3.44) |
| Antibiotics treatment, n (%) | 309 (9.48) | 75 (9.35) | 41 (10.4) | 319 (11.1) |
| Inflammatory bowel diseases, n (%) | 42 (1.29) | 1 (0.13) | 4 (1.87) | 31 (1.16) |
| **Clinical blood pressure** |  |  |  |  |
| 24-hour blood pressure record |  |  |  |  |
| SBP (mmHg) | 122 (10.8) | 122 (11.8) | — | — |
| DBP (mmHg) | 75.7 (7.4) | 75.3 (8.0) | — | — |
| Variability of SBP (mmHg) | 14.7 (3.98) | 14.0 (3.70) | — | — |
| Variability of DBP (mmHg) | 12.2 (3.34) | 11.1 (3.02) | — | — |
| Office blood pressure measurement |  |  |  |  |
| SBP (mmHg) | 123 (15.2) | 121 (16.4) | 124 (16.1) | 120 (15.8) |
| DBP (mmHg) | 75.6 (9.53) | 73.8 (9.90) | 76.9 (9.88) | 73.5 (9.43) |
| **Sodium intake evaluation** |  |  |  |  |
| Urine sodium (mmol/L) | 55.3 (33.3–86.1) | 47.6 (24.7–76.9) | 56.9 (32.5–86.7) | 46.1 (24.5–78.8) |
| Creatinine (mmol/L) | 124 (73.5–180) | 113 (55.1–163) | 141 (78.1–191) | 109 (54.3–165) |
| Estimated 24-hour urine sodium (mg/day)<sup>a</sup> | 8.05 (7.77–8.29) | 8.05 (7.78–8.31) | 8.02 (7.74–8.28) | 8.05 (7.78–8.28) |

ABPM, ambulatory blood pressure monitoring; SBP, systolic blood pressure; DBP, diastolic blood pressure; n, number of participants
Continuous variables are presented as mean (standard deviation) or median (interquartile range) as appropriate. The median was presented for fiber intake, energy intake, urine sodium, creatinine, and estimated 24-hour urine sodium.
<sup>a</sup>Estimated 24-hour urine sodium was based on Kawasaki formula.

### Decreased gut microbiota diversity is associated with higher systolic and diastolic blood pressure

We observed a negative association between Shannon diversity index and 24-hour BP phenotypes, after adjusting for age, sex, country of birth, and technical source of variation (**Table 2**; Model 1). The associations remained after further adjustment for smoking, fiber intake, total energy intake, sodium intake, usage of antidiabetic drugs, and usage of antihyperlipidemic drugs (**Table 2**, Model 2). The associations were clearly attenuated, and in most cases no longer significant at *P*<0.05 after adjusting for BMI, suggesting that BMI may act either as a confounder or mediator of these associations (**Table 2**, Model 3). Results were similar when using office BP as the alternative outcome (**Table 2**).

**Table 2.** Association between alpha diversity (Shannon diversity index) and blood pressure (24-hour BP outcomes and office BP) in the ABPM and non-ABPM subsamples.

| Outcome | Model 1 |  | Model 2 |  | Model 3 |  |
| --- | --- | --- | --- | --- | --- | --- |
|  | Size | Estimate (95% CI) | Size | Estimate (95% CI) | Size | Estimate (95% CI) |
| **ABPM subsample** |  |  |  |  |  |  |
| 24-hour SBP | 4062 | -1.94 (-2.78, -1.10) | 3694 | -1.61 (-2.49, -0.72) | 3694 | -0.40 (-1.24, 0.44) |
| 24-hour DBP | 4062 | -1.10 (-1.67, -0.53) | 3694 | -0.87 (-1.48, -0.27) | 3694 | -0.22 (-0.80, 0.36) |
| Variability of 24-hour SBP | 4062 | -0.60 (-0.91, -0.30) | 3694 | -0.52 (-0.84, -0.20) | 3694 | -0.34 (-0.66, -0.02) |
| Variability of 24-hour DBP | 4062 | -0.61 (-0.87, -0.35) | 3694 | -0.55 (-0.83, -0.28) | 3694 | -0.39 (-0.66, -0.12) |
| Office SBP | 4060 | -1.92 (-3.09, -0.75) | 3693 | -1.30 (-2.53, -0.07) | 3693 | -0.01 (-1.21, 1.19) |
| Office DBP | 4060 | -1.40 (-2.15, -0.65) | 3693 | -0.94 (-1.72, -0.16) | 3693 | 0.06 (-0.69, 0.80) |
| **Non-ABPM subsample** |  |  |  |  |  |  |
| Office SBP | 3261 | -3.24 (-4.53, -1.95) | 2772 | -2.67 (-4.10, -1.23) | 2772 | -1.20 (-2.61, 0.21) |
| Office DBP | 3260 | -1.98 (-2.78, -1.18) | 2771 | -1.52 (-2.41, -0.64) | 2771 | -0.34 (-1.20, 0.52) |

Abbreviations: BP, blood pressure; ABPM, ambulatory blood pressure monitoring; SBP, systolic blood pressure; DBP, diastolic blood pressure; CI, confidence interval
Model 1: Adjusted for age, sex, country of birth, and technical source of variation
Model 2: Adjusted for Model 1 covariates and additionally for smoking, fiber intake, total energy intake, sodium intake, usage of antidiabetic drugs, and usage of antihyperlipidemic drugs
Model 3: Adjusted for Model 2 covariates and additionally for BMI

### Specific metagenomics species were associated with blood pressure in minimally adjusted models

We further investigated the associations between 1692 species and the 24-hour BP outcomes. In Model 1, adjusting for age, sex, country of birth, and technical source of variation, we found 41 species positively associated with 24-hour SBP, 41 with 24-hour DBP, 27 with the variability of SBP, and 17 with the variability of DBP (**Supplementary Table S1**). We identified 52 species negatively associated with 24-hour SBP, 39 with 24-hour DBP, 42 with variability of SBP, and 25 with variability of DBP. No genera were enriched for associations (**Supplementary Table S2**) after accounting for multiple testing.

### Gut metabolic functional pathways were enriched among the associations between gut microbial species and 24-hour BP measurements

To investigate the potential functional relationship between species and 24-hour BP, we performed an enrichment analysis for GMM based on results from Model 1 (**Supplementary Table S3**). Out of the 103 GMM tested, 22 were enriched in those associations with a positive coefficient. However, no negative association was found. Specifically, 10 GMM were involved in amino acid degradation, 5 in carbohydrate degradation, 5 in central metabolism, and 2 in organic acid metabolism. Threonine degradation I and *Bifidobacterium* shunt were enriched among the associations between the species and all 24-hour BP outcomes (**Supplementary Figure S3**).

### Specific metagenomics species linked to blood pressure in additionally adjusted models

A total of 140 unique species, with the exception of *Eubacteriales* sp. (HG3A.0335) with a non-significant result in Model 1, were taken forward to Models 2 and 3. These 139 species were associated with the corresponding 24-hour BP outcomes in Model 2 at FDR <5%, which was additionally adjusted for smoking, fiber intake, total energy intake, sodium intake, usage of antidiabetic drugs, and usage of antihyperlipidemic drugs (**Supplementary Table S4, Figure 1)**. However, the associations between *Eubacterium ramulus, Eggerthellales* sp. (HG3A.1570), and *Eubacteriales* sp. (HG3A.0447) with 24-hour SBP, 24-hour DBP, and variability of SBP, respectively, were considered unreliable because the relationships were driven by a single influential observation (**Supplementary Table S4, Supplementary Figure S4**). Thus, we deemed 136 species to be robustly associated with at least one blood pressure trait.

**Figure 1.**
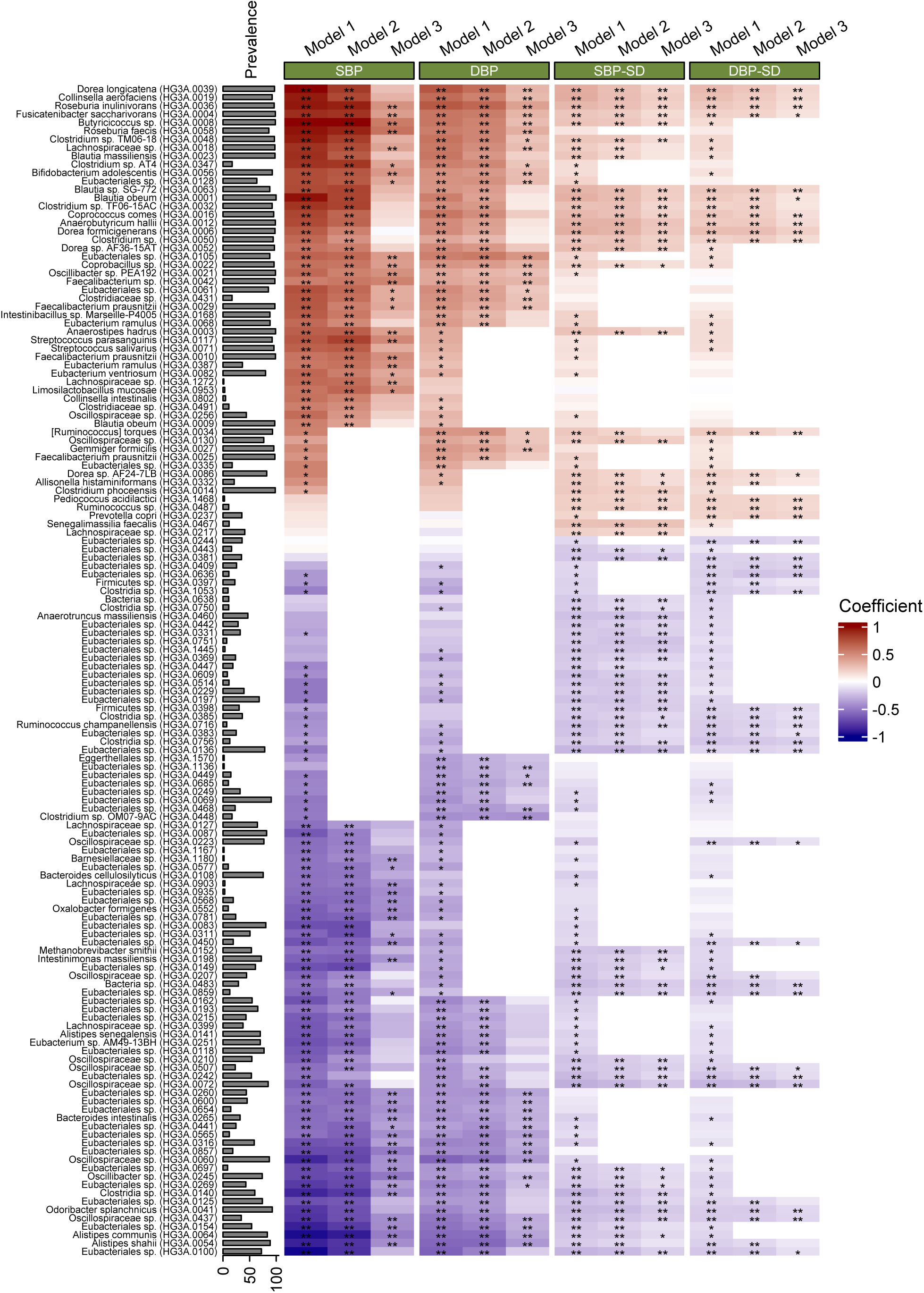
Association between metagenomic species and 24-hour blood pressure measurements and their variability from multivariable regression analyses. In Model 1, adjustment was made for age, sex, country of birth, and DNA extraction plate. Model 2 was adjusted for Model 1 covariates and additionally for smoking, fiber intake, total energy intake, sodium intake, usage of antidiabetic drugs, and usage of antihyperlipidemic drugs. In Model 3, additional adjustment for BMI was included. Y-axis shows the species name, and the internal identification number (HG3A.number) in parenthesis. Abbreviations: SBP: systolic blood pressure, DBP: diastolic blood pressure, SBP-SD: variability of 24-hour SBP, DBP-SD: variability of 24-hour DBP. ** indicates associations with FDR <5%, while * indicates associations with *P*-value <0.05.

Species with a prevalence >70% showed consistent results across 24-hour BP and office BP **(Figure 2**). In this group, the species with the largest positive effect estimates in Model 2 were *Streptococcus parasanguinis*, *Butyricicoccus* sp. (HG3A.0008), *Clostridium* sp. TM06−18, *Clostridium* sp. TF06−15AC (HG3A.0032), *S. salivarius*, *Collinsella aerofaciens*, and *Dorea longicatena.* The largest negative effect estimates were observed for *Alistipes communis, A. shahii*, and *Eubacteriales* sp. (HG3A.0100) (**Figure 2**). For species detected in ≤70% of participants, the results were somewhat less consistent (**Supplementary Figure S5**).

**Figure 2.**
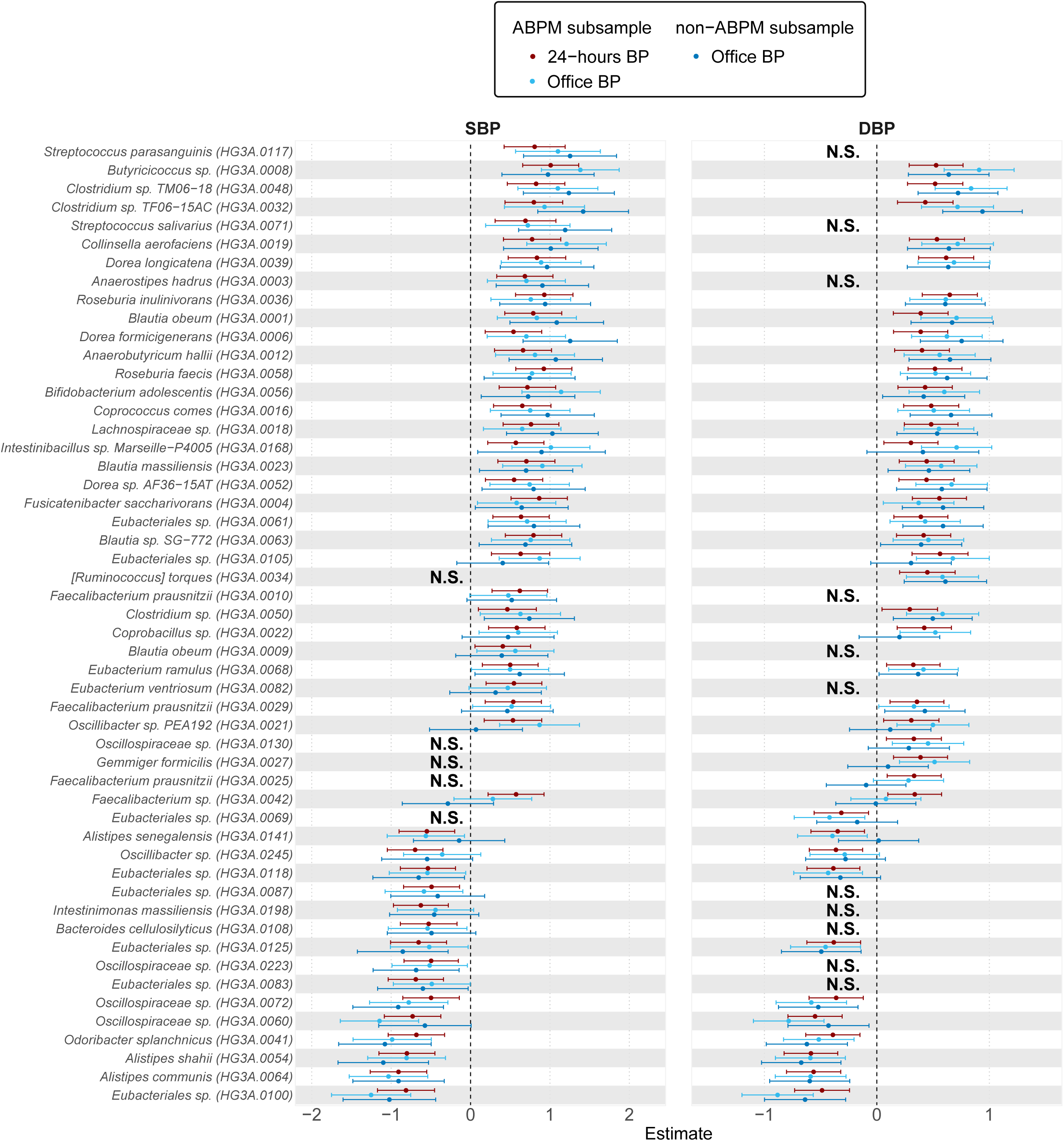
Forest plot of the association between metagenomic species (MGS, prevalence >70%) and blood pressure in two subsamples, ABPM and non-ABPM. Forest plot of the association between MGS (prevalence rate >70%) and (left panel) mean 24-hour SBP (ABPM subsample), mean office SBP (ABPM subsample), and mean office SBP (non-ABPM cohort) in Model 2. The association between MGS and (right panel) mean 24-hour DBP (ABPM subsample), mean office DBP (ABPM subsample), and mean office DBP (non-ABPM subsample) in Model 2. Model 2 was adjusted for age, sex, country of birth, DNA extraction plate, smoking, fiber intake, total energy intake, sodium intake, usage of antidiabetic drugs, and usage of antihyperlipidemic drugs. N.S. referred to the non-significant associations between MGS and BP in Model 1.

*Dorea formicigenerans*, *D. longicatena*, *Anaerobutyricum hallii*, *Blautia* sp. SG-772, *B. obeum,* and *Roseburia inulinivorans* were the species with the largest positive association with variability of 24-hour BP in Model 2, while *Odoribacter splanchnicus*, *Oscillospiraceae* sp. (HG3A.0072), *Eubacteriales* sp. (HG3A.0100, 0136) and *Clostridia* sp. (HG3A.0140) were the largest negatively associated species (**Supplementary Table S4**).

Associations between individual species and BP were attenuated after adjusting for BMI (**Supplementary Table S5**). We observed evidence for BMI-independent associations at FDR <5% with 24-hour BP for 50 species, of which 14 were deemed to be driven by a single observation associated to either 24-hour SBP or 24-hour DBP. Among these species, there were nine that had a reliable association with the other trait of 24-hour BP (**Supplementary Table S5, Figure 1**). Therefore, among the remaining 36 species encompassing 45 reliable associations, the largest BMI-independent positive effect estimates were observed for *R. faecis, R. inulinivorans, Eubacteriales* sp. (HG3A.0105), *Coprobacillus* sp. (HG3A.0022), *Faecalibacterium* sp. (HG3A.0042), and *Fusicatenibacter saccharivorans*. The largest negative effect estimates were observed for *A. shahii and A. communis.* There were 57 species associated with BP variability after adjusting for BMI of which 7 were driven by a single observation. Among the remaining 50 species *Senegalimassilia faecalis, R. inulinivorans* and *A. butyriciproducens* were examples of species positively associated with variability of 24-hour BP. *Anaerotruncus massiliensis* was an example of a species negatively associated with variability of 24-hour BP.

### Sensitivity analyses confirmed robust associations between gut microbial species and blood pressure

We performed sensitivity analyses to assess the robustness of the associations between species and 24-hour BP outcomes. First, medication usage and certain medical conditions can significantly influence the gut microbiota and the outcomes. We conducted sensitivity analyses to assess the robustness of the associations between species and 24-hour BP outcomes by repeating the analyses while excluding participants who had been exposed to antibiotics within the previous 6 months or who had inflammatory bowel diseases. Additionally, we introduced an adjustment for the use of PPIs to account for their potential impact (**Supplementary Table S6**). In these analyses, estimates and *P*-values were consistent (**Supplementary Figure S6**).

### 24-hour BP-associated species are associated with many plasma metabolites, and their clustering reflects the directionality of the associations between species and 24-hour BP outcomes

We assessed the associations between the 136 24-hour BP-associated species with plasma metabolites using the GUTSY Atlas^11^. Each of these species were associated with at least 73 metabolites. (**Supplementary Table S7**).

The species associated with 24-hour BP formed two clusters based on their strongest associations with metabolites, closely corresponding to their respective positive and negative associations with 24-hour SBP (**Supplementary Figure S7**). With the exception of a subgroup of 12 species, the species in the cluster positively associated with 24-hour SBP were positively correlated with secondary bile acids and negatively associated with primary bile acids, vitamin A metabolites, and acetylated peptides, among others. Conversely, the other cluster showed an inverse pattern for these associations. The previously observed inverse clustering pattern for the associations between 24-hour BP-associated species and metabolites was remarkably similar to the associations between variability of 24-hour BP species and metabolites (**Supplementary Figure S8**).

## Discussion

The current study of 4063 participants is the first large population-based study to describe the relationship of the human gut microbiota with 24-hour ABPM. We identified 140 species associated with at least one of the four 24-hour ABPM measurements. Among common species, *Agathobaculum butyriciproducens, Roseburia faecis, R. inulinivorans,* and *Dorea longicatena*, had the largest positive effect estimates on SBP and DBP, whereas the largest negative effect estimates were observed for *Alistipes communis, A. shahii*, and *Eubacteriales* sp. (HG3A.0100) in our adjusted Model 2. *D. formicigenerans*, *D. longicatena*, *Anaerobutyricum hallii*, *Blautia* sp. SG-772, and *Collinsella aerofaciens* were the species with a larger positive association with variability of 24-hour BP, while *Odoribacter splanchnicus*, *Oscillospiraceae* sp. (HG3A.0072), *Eubacteriales* sp. (HG3A.0100) and *Clostridia* sp. (HG3A0756) were the species with largest negative effect estimates.

The gold standard for determining the mean BP in an individual is with 24-hour ABPM, as it can account for, and quantify the substantial within-person variation in blood pressure over time. Its clinical practical application is widely acknowledged, and its usefulness for population research has recently been also recognized.^10^ Prior to this study, the association between gut microbiota and within-person variation in BP had not been investigated.

Two large studies have evaluated the association between the gut microbiota and office BP.^7, 22^ The HELIUS study used 16S rRNA gene amplicon sequencing to examine the gut microbiota of 4672 participants from the municipality of Amsterdam.^8^ After applying machine learning to identify the microbial predictors of BP, higher abundance of *Streptococcus* spp. was found to be associated with higher SBP. Similarly, *S. parasanguinis* was associated with higher BP in the present study. In the HELIUS study, increased *Roseburia* spp. abundance was inversely associated with SBP. However, in the present study, *R. faecis* and *R. inulinivorans* were positively associated with BP measurements. This divergence in findings between the HELIUS study and ours could be due to differences in the ethnic groups under investigation and differences between individual species within the genus *Roseburia*.

In the FINRISK 2002 study, 6953 participants had their gut microbiota profiled using shallow metagenomic sequencing.^22^ They found that, after multivariable adjustment including BMI and antihypertensive medication, 45 microbial genera were associated with BP. Among these, 20 genera were positively associated with prevalent hypertension, including the genus *Dorea*. *Dorea* spp. are gram positive obligatory anaerobic bacteria belonging to the family *Lachnospiraceae*.^23^ In our study, we found that *D. formicigenerans* and *D*. *longicatena* were positively associated with SBP and DBP. Higher abundance of *D. formicigenerans* was correlated with higher BMI in a previous study.^24^ However, the subspecies of *D*. *longicatena* have varied associations with BMI.^25^ When we adjusted our analysis for BMI, we no longer observed an association between *D. formicigenerans* and SBP or DBP, nor between *D. longicatena* and SBP. There are two possible explanations for these results. First, BMI could be a marker of poor lifestyle or other unmeasured confounders that affect both microbial species and BP.^26^ Second, given that certain species might cause changes in adiposity,^27^ BMI could be a mediator of the species associations with BP. Given that increased BMI is a well-known contributing cause of hypertension,^28–30^ longitudinal studies with comprehensive information are needed to better understand whether specific gut microbiota species might have an impact on BP mediated by changes in BMI.

In our study, *R. faecis*, also known as *Agathobacter faecis*, was positively associated with SBP and DBP, even after BMI adjustment. Conversely, a higher abundance of *R. faecis* has been described in subjects with a higher adherence to the Mediterranean diet ^31^, which was shown to be protective against hypertension in clinical trials^32, 33^. One possible reason for these contradictory findings is that certain components of the Mediterranean diet might have a greater impact on hypertension while other components are more associated to the *R. faecis* abundance. For instance, high intake of fiber and fruits have been associated with *R. faecies*,^31^ while the strongest lowering effects on 24-hour SBP and DBP have been found in the Mediterranean diet supplemented with extra virgin olive oil.^31^

The production of SCFA by the gut microbiota has been suggested to affect the host BP.^34–37^ In the present study, one of the SCFA, specifically butyrate/isobutyrate (4:0), was found to be linked to 59 species associated with BP. In addition to the butyrate-producing *R. faecis*, we also found that *Faecalibacterium prausnitzii* and *Bifidobacterium adolescentis* were positively associated with SBP and DBP. An *in vitro* study has shown that co-cultures of *F. prausnitzii* with *B. adolescentis* result in higher butyrate production than monocultures.^34^ Of note is that we had six within-species variants of *F*. *prausnitzii* in the dataset, but only three of these variants (HG3A.0010, 0025, 0029) were associated with BP, indicating that there are differences on substrain level. Increased fecal concentrations of SCFA have been described in subjects with elevated SBP in the HELIUS study^8^ as well as in other studies^36, 37^. Furthermore, the serum concentration of the SCFA acetate was positively associated with SBP in the longitudinal FINRISK 2007 study (n=4197).^35^ However, in the cross-sectional analysis of FINRISK (n=36 985), serum acetate was inversely associated with hypertension in women.^35^ Despite the positive associations between serum and fecal SCFA and BP in some observational studies, animal studies have produced opposite results. Oral supplementation of acetate led to a reduction in SBP and DBP in hypertensive mice models^38^ and intracolonic or intravenous administration of butyrate produced a reduction in BP in rats.^39^ Furthermore, although we found certain butyrate-producing bacteria positively associated with BP, the butyrate absorption might be reduced in individuals with elevated BP, similar to that reported in a cross-sectional study (n=61) that reported a low plasma SCFA concentration concomitant with high fecal concentrations in hypertensive individuals.^37^

We found several species that were inversely associated with BP even after adjustment for BMI, including *Oxalobacter formigenes*, *Odoribacter splanchnicus*, *A. communis,* and *A. shahii*. *O. formigenes* depends on oxalate as an energy-and carbon source and individuals colonized with this bacterium show a lower urinary oxalate excretion and thereby may have a lower risk of kidney lesion by calcium oxalate stones.^40^ *O. splanchnicus*, an anaerobic gram-negative bacterium,^41^ has been associated with lower vascular stiffness measured with pulse wave velocity,^42^ which is a predictor for cardiovascular diseases.^43^ In line with our findings, a combined analysis of the genus *Alistipes* previously showed an inverse association with cardiovascular diseases such as atrial fibrillation,^44^ atherosclerotic cardiovascular diseases,^45^ and heart failure.^46^ Moreover, an animal study suggested that *A. putredinis* might contribute to alleviate obesity induced by the fecal microbiota transplantation from obese humans to mice.^47^ Conversely, Kim et al. described a positive correlation of two *Alistipes* species, *A. finegoldii* and *A. indistinctus*, with SBP.^48^ In our study, these two species were not associated with BPs in Model 1. Together, these studies indicated that the associations of *Alistipes* species with BP might be heterogeneous.

We found that the functional pathways of threonine degradation I and *Bifidobacterium* shunt were enriched among the species positively associated with BP or BP variability. In a large prospective study (n=4288), a pattern of higher dietary intake of threonine, serine, branched chain, and aromatic amino acids was associated with incident hypertension.^49^ In contrast, an inverse association between threonine intake and BP has been described in a longitudinal post-myocardial infarction study (n=100)^50^ and in a cross-sectional study of middle-aged men (n=92).^51^ Given these conflicting results of dietary amino acids and BP, together with our current results, future investigations should examine whether the gut microbiota might act as an effect modifier on the relationship of threonine with BP.

*Bifidobacterium* shunt is a fermentation pathway unique to *Bifidobacterium* strains that yields acetate and lactate as end products.^52–54^ Although, we observed a positive association between *Bifidobacterium* shunt and BP, a reduction in the abundance of genus *Bifidobacterium* has been linked to elevated blood pressure in children with type 1 diabetes mellitus.^55^ Additionally, *B. breve* has been found to prevent deoxycorticosterone acetate (DOCA)-salt hypertension in rats possibly mediated by increased acetate production.^53, 56^ More studies are needed to investigate the association of *Bifidobacterium* species with blood pressure and explore potential contributing mechanisms.

The lactate production pathway was also enriched in species associated with DBP in our study. A higher abundance of lactate-producing bacteria has also been described in spontaneously hypertensive rats.^57^ Plasma lactate has been associated with incident hypertension in women in a large epidemiological study (n=5554). The authors conjectured that the higher lactate reflected decreased oxidative capacity or insufficient vascular capacity leading to increased BP.^58^ Although the human gut microbiota contribution to plasma lactate levels is unknown, germ-free mice fed lactate-producing *Lactobacillus* had higher circulating lactate levels than controls fed *Lactobacillus* strains lacking the lactate production pathway.^59^ Instead of crossing into the blood stream, a usual route for the lactate produced in the colon is the conversion into butyrate,^60^ which may correspond to the increased fecal SCFA observed in individuals with hypertension.^37^

## Strengths and limitations

The present study has several strengths. This is the first large population-based study to investigate the relationship of the gut microbiota with 24-hour ABPM measurements. As elaborated above, 24-hour ABPM provides a more precise and comprehensive assessment of BP compared to office measurements. Additionally, the gut microbiota was assessed using deep shotgun metagenomics enabling high resolution detection of gut microbiota species and the putative functional pathways carried by these species. Moreover, the extensive survey of covariates made it possible to adjust for several potential confounders. We also excluded participants with antihypertensive medication usage, defined by ATC codes, to avoid identifying associations confounded by hypertension treatment.

There are, however, limitations that need to be addressed. First, this study has a cross-sectional design and causality cannot be inferred. Moreover, we cannot rule out collider bias by selection on health-seeking behavior, residual confounding, or reverse causation. For instance, participants with higher blood pressure might have received recommendations of lifestyle modifications that could affect the gut microbiota. Second, formal mediation analysis for BMI was not possible due to lack of temporal precedence in the present study. We expect to be able to address the longitudinal associations between gut microbiota and BP in follow up examinations of SCAPIS. Third, the dietary assessment was based on a self-reported questionnaire that may be affected by recall and report bias. Misclassification of dietary intake has been related to factors that are relevant for the gut microbiota composition, such as sex, age or obesity.^61^ Therefore, we cannot exclude the possibility of differential misclassification of the dietary covariates dependent on the exposure, which would result in residual confounding or additional distortion of estimates. Fourth, our findings would need replication in an independent cohort with complete 24-hour ABPM and gut microbiota data. We were able to use an independent non-ABPM subsample to validate the results from the ABPM subsample, and it showed the generalizability of the results in office BP. Fifth, although we estimated salt intake using urine sodium from spot samples, this method has been shown to have individual-level inconsistencies when compared with 24-hour urine samples.^62^ Sixth, fecal metagenomics, while powerful for describing a snapshot of the microbial community, does not accurately capture the gut microbiota species that reside closer to the mucosal membrane^63^ or in the small intestines,^64^ which is a limitation of most population-based studies on gut microbiota.

## Conclusion

In our large population-based sample, the associations between the gut microbiota, plasma metabolites, and 24-hour BP were investigated. We identified 140 gut microbiota species associated with 24-hour ABPM. Moreover, we found indications of involvement of gut microbial threonine degradation I, *Bifidobacterium* shunt, and lactate production, and 24-hour BP phenotypes in BP regulation. Our findings provide a starting point for further studies on the role of human gut microbiota and downstream metabolites in BP regulation, which can potentially be targets for future studies.

## Data Availability

De-identified de-hosted metagenomic sequencing data for SCAPIS samples can be accessed from the European Nucleotide Archive under accession number PRJEB51353 (https://www.ebi.ac.uk/ena/browser/view/prjeb51353). Access to pseudonymized SCAPIS phenotype data requires ethical approval from the Swedish Ethical Review Board and approval from the SCAPIS Data access board (https://www.scapis.org/data-access/). The source code and the summary data underlying all figures used to generate the results for the analysis are available at https://github.com/MolEpicUU/24hBP-mgs.

## Author contributions

J.Ä., J.S., G.Be, G.E., J.G.S., M.O-M. and T.F. obtained the funding. Y-T.L., U.H., S.S-B., J.S., J.Ä., M.O-M. and T.F. designed the study and developed the concept. L.L., G.E., J.G.S., J.S., J.Ä., M.O-M. and T.F. collected the data. N.N., A.C.E., J.B.H., H.B.N. performed metagenomic analysis and bioinformatics, Y-T.L., K.F.D., S.S-B. performed quality control and/or data management, Y-T.L. and S.S-B. performed the association analyses, Y-T.L., K.F.D., S.S-B. and G.V. visualized the results, and M.O-M. and T.F. coordinated the study. Y-T.L., G.Ba., D.N., and T.F. wrote the first draft of the manuscript. All authors contributed with the interpretation of the results and critical revision of the manuscript.

## Acknowledgments

The computations and data handling were enabled by resources in project sens2019512 provided by the National Academic Infrastructure for Supercomputing in Sweden (NAISS) and the Swedish National Infrastructure for Computing (SNIC) at Uppsala Multidisciplinary Center for Advanced Computational Science (UPPMAX) partially funded by the Swedish Research Council through grant agreements no. 2022-06725 and no. 2018-05973. The main funding body of the Swedish CArdioPulmonary bioImage Study (SCAPIS) is the Swedish Heart-Lung Foundation. The study is also funded by the Knut and Alice Wallenberg Foundation; the Swedish Research Council, and VINNOVA (Sweden’s Innovation agency); the University of Gothenburg and Sahlgrenska University Hospital; Karolinska Institutet and Stockholm county council; Linköping University and University Hospital; Lund University and Skåne University Hospital; Umeå University and University Hospital; and Uppsala University and University Hospital.

