## Supplementary Methods and Figures S1-S8 for "The association between the gut microbiome and 24-hour blood pressure measurements in the SCAPIS study"

**Blood pressure measurements**

Participants wore the ABPM device (Labtech EC-3H/ABP, Labtech Ltd, Debrecen, Hungary) for one day. SBP and DBP were measured automatically every 30 minutes in the participants from the Malmö center and every 30 minutes during the day and every 90 minutes during the night in participants from the Uppsala center. This nighttime restriction in Uppsala was applied because of a simultaneous sleep registration. Individuals with fewer than 10 readings during daytime or fewer than 5 readings during nighttime were not considered for further analyses. Daytime and nighttime were defined using narrow fixed clock-time periods as 10 AM to 8 PM and 12 AM to 6 AM, respectively, to eliminate the influence of BP variation during the sleep-awake transition.[1] To avoid an overestimation of mean 24-hour BP due to a higher number of readings per hour during daytime, we performed an inverse weighted estimate based on the time interval between measurements.[2,3] Office BP was measured placing the cuff at the upper arm at the heart level, and the cuff size was adjusted according to individual arm circumference. Measurements were repeated until two subsequent results were within ±10 mmHg with a maximum number of attempts of four. The mean of the two measurements in each arm was used as office BP.

**Phenotypes**

The participants who had a prescription for antihypertensive medications for the previous 12 months before the visit 1 were obtained from the Swedish Prescribed Drug Register and included the Anatomical Therapeutic Chemical Classification System (ATC) codes C02, C03A, C03EA01, C07, C08C, and C09. Prescriptions for antibiotics for the previous 6 months before the visit 1 included J01CE02, J01CF05, J01EA01, J01FA01, J01FA06, J01FA09, J01FA10, J01FF01, J01XC01, J01XE01, J01AA02, J01AA04, J01AA06, J01AA07, J01CA04, J01CA08, J01CR02, J01DB05, J01DD14, J01EE01, J01MA02, J01MA06, J01MA12, J01MA14, J01XX05, and J01XX08. Participants with prescriptions for narrow-spectrum and/or for broad-spectrum were considered as participants treated with antibiotics.

### Supplementary Figures

#### Figure S1. Flowchart of study design

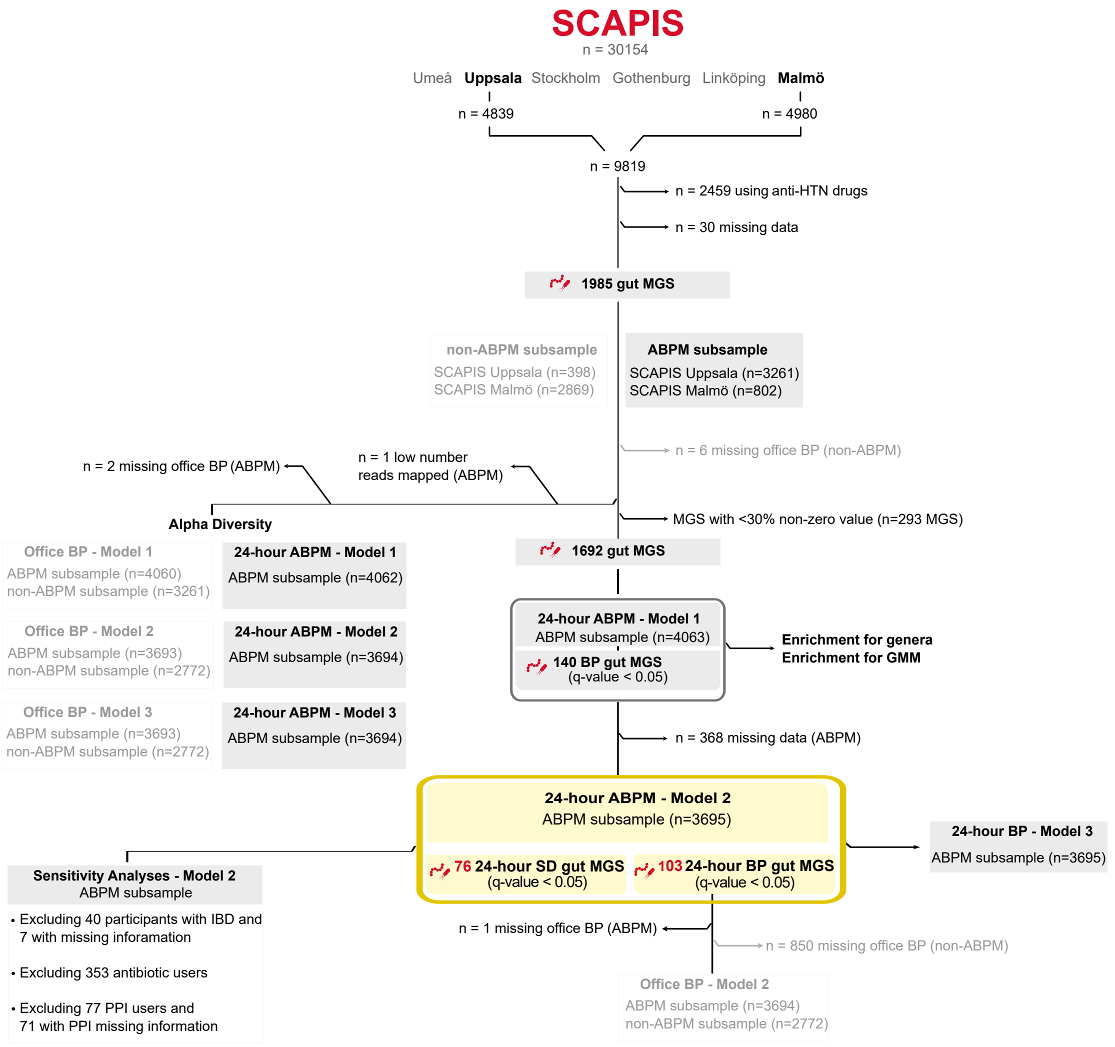

#### Figure S2. Directed acyclic graph

Proposed directed acyclic graph for the association between gut microbiome and blood pressure. (A) BMI is considered a mediator. (B) BMI is considered a confounding factor. A directed edge (or “arrow”) from one node to another represents a direct effect between these two nodes. The exposure (gut microbiome) is denoted by the green oval with the play symbol. The outcome (blood pressure) is denoted by the blue oval with the bar. The causal paths are noted in green lines and the biasing paths are in magenta. Blue oval (empty) represented a mediator. Pink oval represented the potential confounding factors.

A

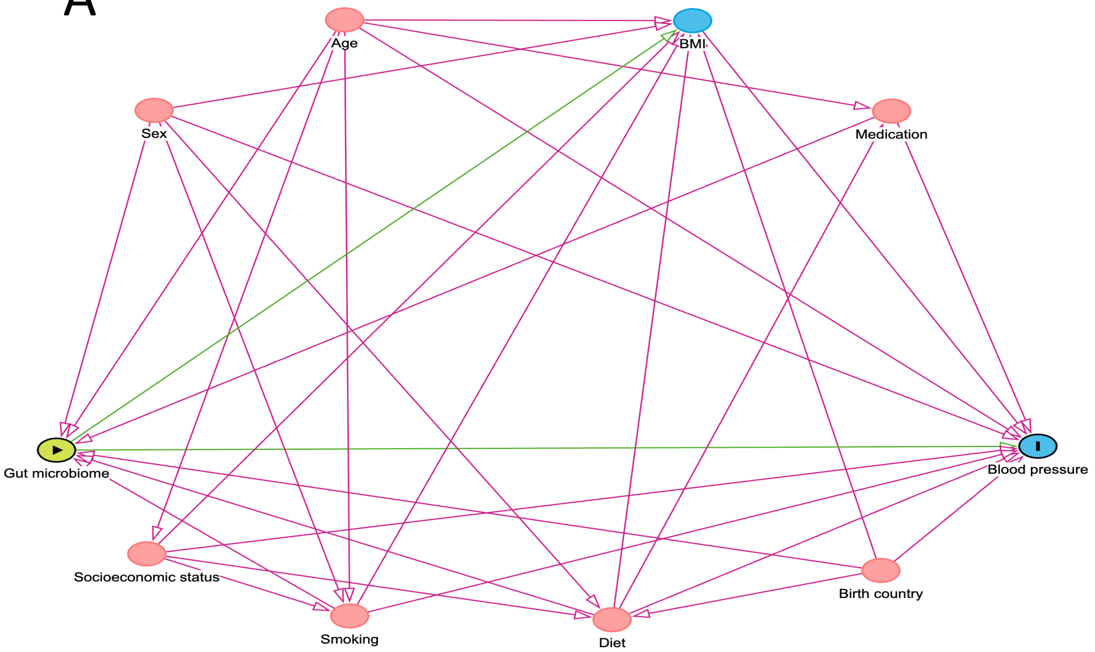
Age
BMI
Sex
Medication
Gut microbiome
Blood pressure
Socioeconomic status
Birth country
Smoking
Diet

B

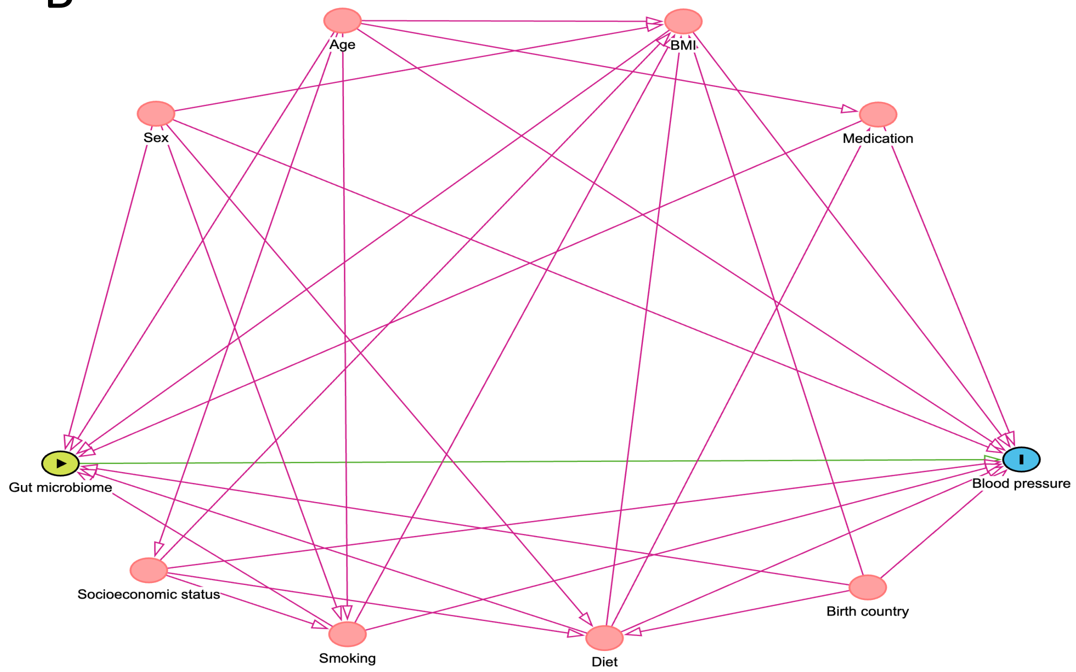
Age
BMI
Sex
Medication
Gut microbiome
Blood pressure
Socioeconomic status
Birth country
Smoking
Diet

#### Figure S3. Venn diagrams of enriched gut metabolic modules

Venn diagram displaying the enriched GMM using taxon set enrichment analysis. This analysis was based on the ranked *P*-values of the associations between metagenomics species and mean 24-hour BP outcomes stratified by effect direction from Model 1 in the ABPM subsample.

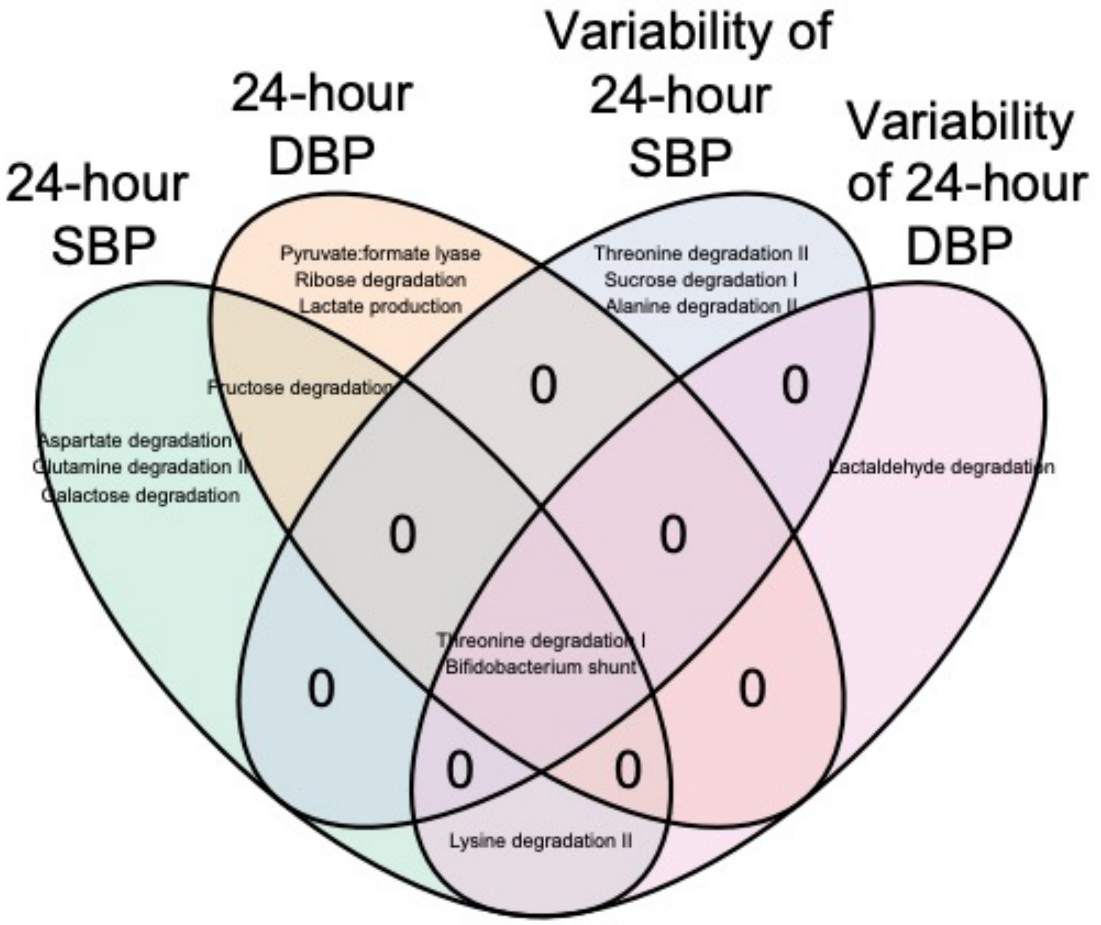

Figure S4. Venn diagram of 24-hour blood pressure outcomes in Model 2

Venn diagram displaying the metagenomic species associated with mean 24-hour SBP, mean 24-hour DBP, and their variability in the ABPM subsample using Model 2.

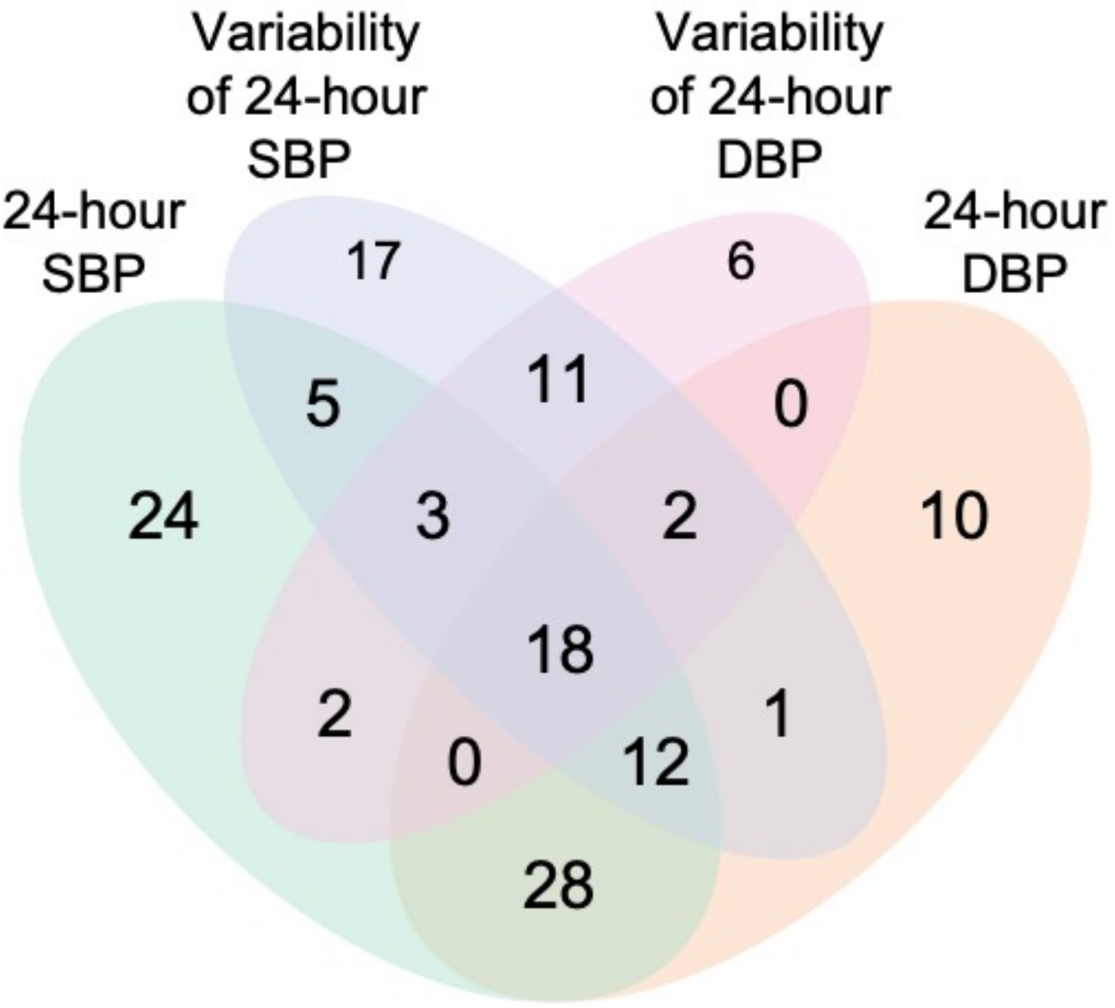

**Figure S5:** Forest plot of the association between metagenomic species (MGS, prevalence rate ≤70%) and blood pressure in two subsamples, ABPM and non-ABPM.

Forest plot of the association between MGS (prevalence rate ≤70%) and (left panel) mean 24-hour SBP (ABPM subsample), mean office SBP (ABPM subsample), and mean office SBP (non-ABPM subsample) in Model 2. The association between MGS and (right panel) mean 24-hour DBP (ABPM subsample), mean office DBP (ABPM subsample), and mean office DBP (non-ABPM subsample) in Model 2. Model 2 was adjusted for Model 1 covariates and additionally for smoking, fiber intake, total energy intake, sodium intake, usage of antidiabetic drugs, and usage of antihyperlipidemic drugs. N.S. referred to the non-significant associations between MGS and BP in Model 1.

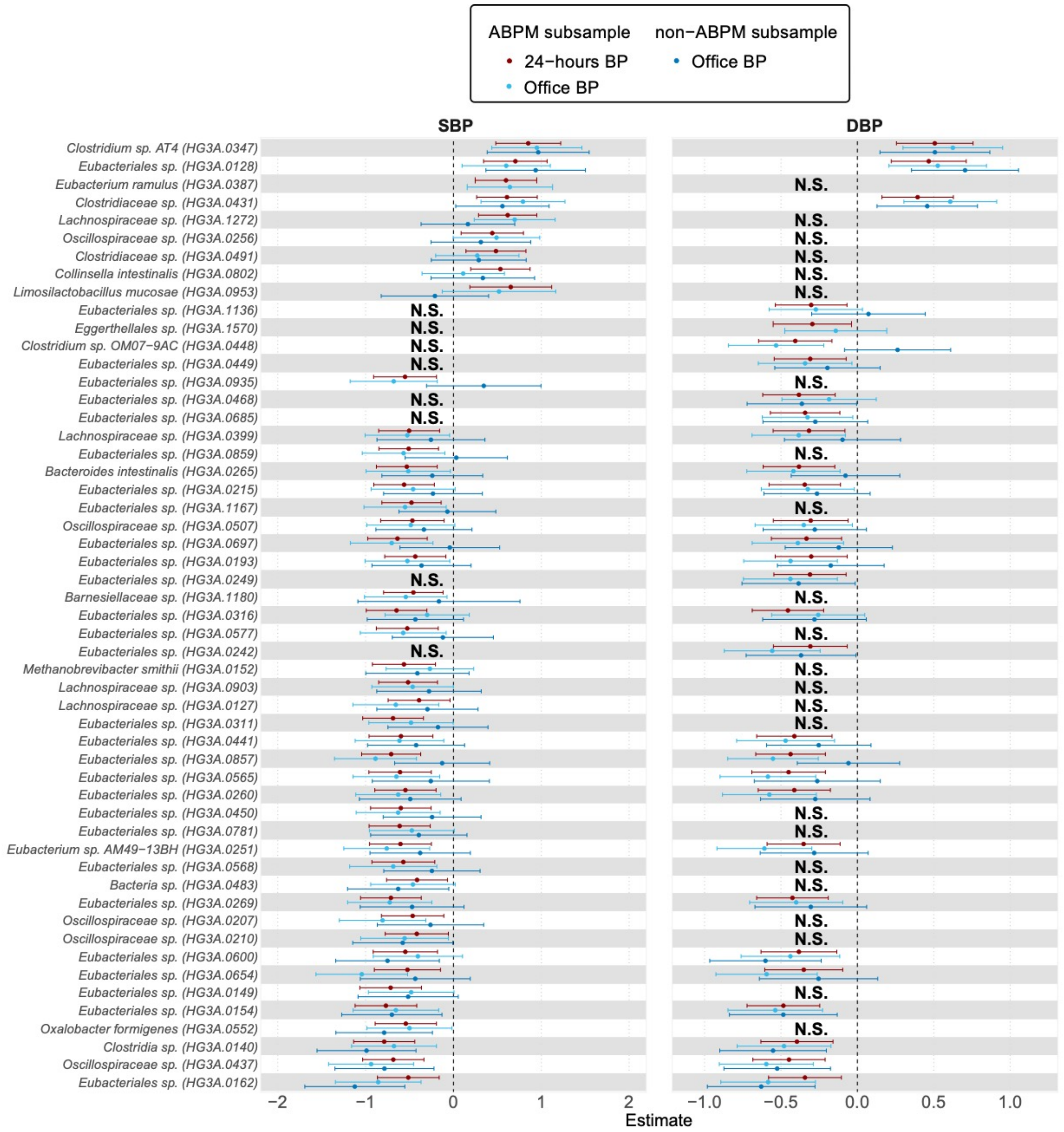
ABPM subsample
24−hours BP
Office BP
non−ABPM subsample
Office BP
SBP
DBP
Clostridium sp. AT4 (HG3A.0347)
Eubacteriales sp. (HG3A.0128)
Eubacterium ramulus (HG3A.0387)
Clostridiaceae sp. (HG3A.0431)
Lachnospiraceae sp. (HG3A.1272)
Oscillospiraceae sp. (HG3A.0256)
Clostridiaceae sp. (HG3A.0491)
Collinsella intestinalis (HG3A.0802)
Limosilactobacillus mucosae (HG3A.0953)
Eubacteriales sp. (HG3A.1136)
Eggerthellales sp. (HG3A.1570)
Clostridium sp. OM07−9AC (HG3A.0448)
Eubacteriales sp. (HG3A.0449)
Eubacteriales sp. (HG3A.0935)
Eubacteriales sp. (HG3A.0468)
Eubacteriales sp. (HG3A.0685)
Lachnospiraceae sp. (HG3A.0399)
Eubacteriales sp. (HG3A.0859)
Bacteroides intestinalis (HG3A.0265)
Eubacteriales sp. (HG3A.0215)
Eubacteriales sp. (HG3A.1167)
Oscillospiraceae sp. (HG3A.0507)
Eubacteriales sp. (HG3A.0697)
Eubacteriales sp. (HG3A.0193)
Eubacteriales sp. (HG3A.0249)
Barnesiellaceae sp. (HG3A.1180)
Eubacteriales sp. (HG3A.0316)
Eubacteriales sp. (HG3A.0577)
Eubacteriales sp. (HG3A.0242)
Methanobrevibacter smithii (HG3A.0152)
Lachnospiraceae sp. (HG3A.0903)
Lachnospiraceae sp. (HG3A.0127)
Eubacteriales sp. (HG3A.0311)
Eubacteriales sp. (HG3A.0441)
Eubacteriales sp. (HG3A.0857)
Eubacteriales sp. (HG3A.0565)
Eubacteriales sp. (HG3A.0260)
Eubacteriales sp. (HG3A.0450)
Eubacteriales sp. (HG3A.0781)
Eubacterium sp. AM49−13BH (HG3A.0251)
Eubacteriales sp. (HG3A.0568)
Bacteria sp. (HG3A.0483)
Eubacteriales sp. (HG3A.0269)
Oscillospiraceae sp. (HG3A.0207)
Oscillospiraceae sp. (HG3A.0210)
Eubacteriales sp. (HG3A.0600)
Eubacteriales sp. (HG3A.0654)
Eubacteriales sp. (HG3A.0149)
Eubacteriales sp. (HG3A.0154)
Oxalobacter formigenes (HG3A.0552)
Clostridia sp. (HG3A.0140)
Oscillospiraceae sp. (HG3A.0437)
Eubacteriales sp. (HG3A.0162)
N.S.
−2
−1
0
1
2
−1.0
−0.5
0.0
0.5
1.0
Estimate

#### Figure S6. Sensitivity analyses

Association between results from the Model 2 with 24-hour blood pressure outcomes in which participants who underwent antibiotic treatment (Antib) during the previous 6 months before visit 1, had inflammatory bowel diseases (IBD), had proton-pump inhibitors (PPI) measured in plasma, were excluded.

[Please see separate PDF file for Figure S6; there are 31 pages.]

##### Figure S7. Heatmap of associations between 24hour mean BP-associated species and plasma metabolites.

Data were downloaded from the GUTSY Atlas (Supplementary Tables 4 and 6, https://gutsyatlas.serve.scilifelab.se/), which is based on the same metagenomics data as the current study. For each 24-hour BP-associated species, the three strongest associations with known metabolites were selected based on their *P* value, and the Spearman's rank correlation coefficient of the unique subset is shown. Hierarchical clustering was performed based on the Euclidian distance. Column labeled as 24-hour SBP and 24-hour DBP are the linear regression coefficient of the species with 24-hour SBP and DBP, respectively. The color bar in the bottom depicts the annotated metabolite subclass of the metabolites.

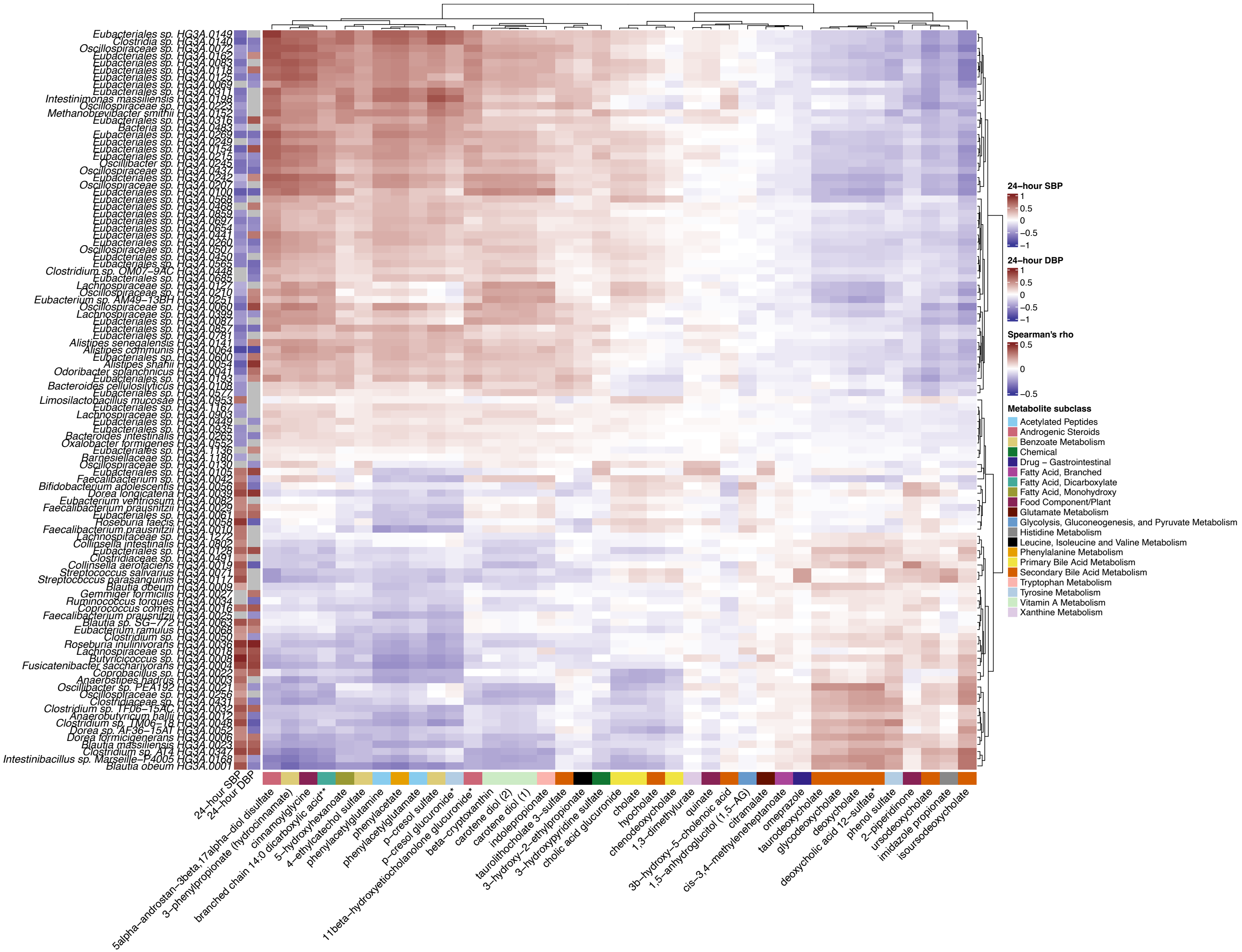

Eubacteriales sp. HG3A.0149
Clostridia sp. HG3A.0140
Oscillospiraceae sp. HG3A.0072
Eubacteriales sp. HG3A.0162
Eubacteriales sp. HG3A.0083
Eubacteriales sp. HG3A.0118
Eubacteriales sp. HG3A.0125
Eubacteriales sp. HG3A.0069
Eubacteriales sp. HG3A.0311
Intestinimonas massiliensis HG3A.0198
Oscillospiraceae sp. HG3A.0223
Methanobrevibacter smithii HG3A.0152
Eubacteriales sp. HG3A.0316
Bacteria sp. HG3A.0483
Eubacteriales sp. HG3A.0269
Eubacteriales sp. HG3A.0249
Eubacteriales sp. HG3A.0154
Eubacteriales sp. HG3A.0215
Oscillibacter sp. HG3A.0245
Oscillospiraceae sp. HG3A.0437
Eubacteriales sp. HG3A.0242
Oscillospiraceae sp. HG3A.0207
Eubacteriales sp. HG3A.0100
Eubacteriales sp. HG3A.0568
Eubacteriales sp. HG3A.0468
Eubacteriales sp. HG3A.0859
Eubacteriales sp. HG3A.0697
Eubacteriales sp. HG3A.0654
Eubacteriales sp. HG3A.0441
Eubacteriales sp. HG3A.0260
Oscillospiraceae sp. HG3A.0507
Eubacteriales sp. HG3A.0450
Eubacteriales sp. HG3A.0565
Clostridium sp. OM07−9AC HG3A.0448
Eubacteriales sp. HG3A.0685
Lachnospiraceae sp. HG3A.0127
Oscillospiraceae sp. HG3A.0210
Eubacterium sp. AM49−13BH HG3A.0251
Oscillospiraceae sp. HG3A.0060
Lachnospiraceae sp. HG3A.0399
Eubacteriales sp. HG3A.0087
Eubacteriales sp. HG3A.0857
Eubacteriales sp. HG3A.0781
Alistipes senegalensis HG3A.0141
Alistipes communis HG3A.0064
Eubacteriales sp. HG3A.0600
Alistipes shahii HG3A.0054
Odoribacter splanchnicus HG3A.0041
Eubacteriales sp. HG3A.0193
Bacteroides cellulosilyticus HG3A.0108
Eubacteriales sp. HG3A.0577
Limosilactobacillus mucosae HG3A.0953
Eubacteriales sp. HG3A.1167
Lachnospiraceae sp. HG3A.0903
Eubacteriales sp. HG3A.0449
Eubacteriales sp. HG3A.0935
Bacteroides intestinalis HG3A.0265
Oxalobacter formigenes HG3A.0552
Eubacteriales sp. HG3A.1136
Barnesiellaceae sp. HG3A.1180
Oscillospiraceae sp. HG3A.0130
Eubacteriales sp. HG3A.0105
Faecalibacterium sp. HG3A.0042
Bifidobacterium adolescentis HG3A.0056
Dorea longicatena HG3A.0039
Eubacterium ventriosum HG3A.0082
Faecalibacterium prausnitzii HG3A.0029
Eubacteriales sp. HG3A.0061
Roseburia faecis HG3A.0058
Faecalibacterium prausnitzii HG3A.0010
Lachnospiraceae sp. HG3A.1272
Collinsella intestinalis HG3A.0802
Eubacteriales sp. HG3A.0128
Clostridiaceae sp. HG3A.0491
Collinsella aerofaciens HG3A.0019
Streptococcus salivarius HG3A.0071
Streptococcus parasanguinis HG3A.0117
Blautia obeum HG3A.0009
Gemmiger formicilis HG3A.0027
Ruminococcus torques HG3A.0034
Coprococcus comes HG3A.0016
Faecalibacterium prausnitzii HG3A.0025
Blautia sp. SG−772 HG3A.0063
Eubacterium ramulus HG3A.0068
Clostridium sp. HG3A.0050
Roseburia inulinivorans HG3A.0036
Lachnospiraceae sp. HG3A.0018
Butyricicoccus sp. HG3A.0008
Fusicatenibacter saccharivorans HG3A.0004
Coprobacillus sp. HG3A.0022
Anaerostipes hadrus HG3A.0003
Oscillibacter sp. PEA192 HG3A.0021
Oscillospiraceae sp. HG3A.0256
Clostridiaceae sp. HG3A.0431
Clostridium sp. TF06−15AC HG3A.0032
Anaerobutyricum hallii HG3A.0012
Clostridium sp. TM06−18 HG3A.0048
Dorea sp. AF36−15AT HG3A.0052
Dorea formicigenerans HG3A.0006
Blautia massiliensis HG3A.0023
Clostridium sp. AT4 HG3A.0347
Intestinibacillus sp. Marseille−P4005 HG3A.0168
Blautia obeum HG3A.0001
24−hour SBP
24−hour DBP
5alpha−androstan−3beta,17alpha−diol disulfate
3−phenylpropionate (hydrocinnamate)
cinnamoylglycine
branched chain 14:0 dicarboxylic acid**
5−hydroxyhexanoate
4−ethylcatechol sulfate
phenylacetylglutamine
phenylacetate
phenylacetylglutamate
p−cresol sulfate
p−cresol glucuronide*
11beta−hydroxyetiocholanolone glucuronide*
beta−cryptoxanthin
carotene diol (2)
carotene diol (1)
indolepropionate
taurolithocholate 3−sulfate
3−hydroxy−2−ethylpropionate
3−hydroxypyridine sulfate
cholic acid glucuronide
cholate
hyocholate
chenodeoxycholate
1,3−dimethylurate
quinate
3b−hydroxy−5−cholenoic acid
1,5−anhydroglucitol (1,5−AG)
citramalate
cis−3,4−methyleneheptanoate
omeprazole
taurodeoxycholate
glycodeoxycholate
deoxycholate
deoxycholic acid 12−sulfate*
phenol sulfate
2−piperidinone
ursodeoxycholate
imidazole propionate
isoursodeoxycholate
24−hour SBP
1
0.5
0
−0.5
−1
24−hour DBP
1
0.5
0
−0.5
−1
Spearman's rho
0.5
0
−0.5
Metabolite subclass
Acetylated Peptides
Androgenic Steroids
Benzoate Metabolism
Chemical
Drug − Gastrointestinal
Fatty Acid, Branched
Fatty Acid, Dicarboxylate
Fatty Acid, Monohydroxy
Food Component/Plant
Glutamate Metabolism
Glycolysis, Gluconeogenesis, and Pyruvate Metabolism
Histidine Metabolism
Leucine, Isoleucine and Valine Metabolism
Phenylalanine Metabolism
Primary Bile Acid Metabolism
Secondary Bile Acid Metabolism
Tryptophan Metabolism
Tyrosine Metabolism
Vitamin A Metabolism
Xanthine Metabolism

**Figure S8.** Heatmap of associations between variability of 24-hour BP-associated species and plasma metabolites.

Data were downloaded from the GUTSY Atlas (Supplementary Tables 4 and 6, https://gutsyatlas.serve.scilifelab.se/), which is based on the same metagenomics data as the current study. For each variability of 24-hour BP-associated species, the three strongest associations with known metabolites were selected based on their *P* value, and the Spearman's rank correlation coefficient of the unique subset is shown. Hierarchical clustering was performed based on the Euclidian distance. Column labeled as variability of 24-hour SBP and 24-hour DBP are the linear regression coefficient of the species with variability of 24-hour SBP and DBP, respectively. The color bar in the bottom depicts the annotated metabolite subclass of the metabolites.

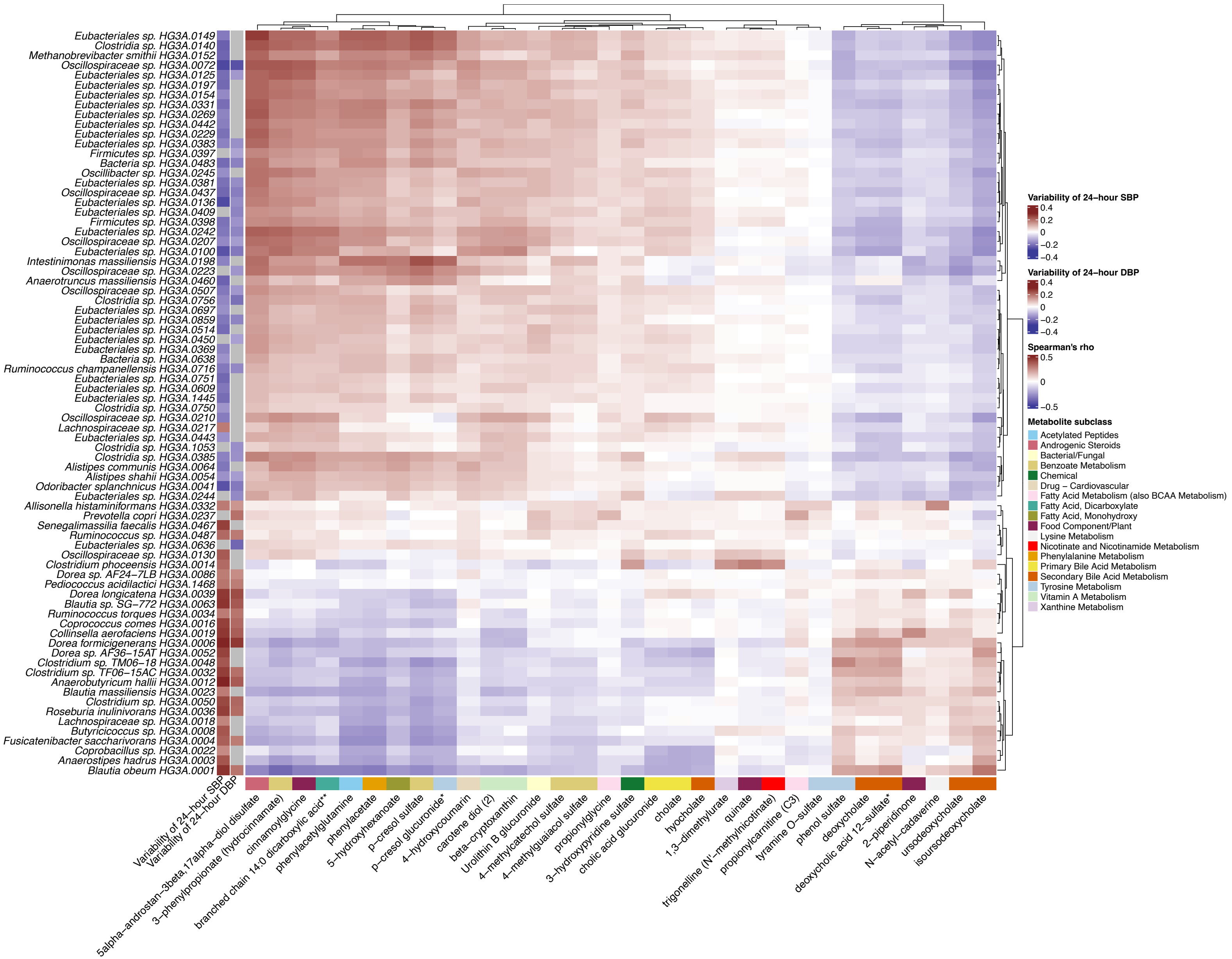

Eubacteriales sp. HG3A.0149
Clostridia sp. HG3A.0140
Methanobrevibacter smithii HG3A.0152
Oscillospiraceae sp. HG3A.0072
Eubacteriales sp. HG3A.0125
Eubacteriales sp. HG3A.0197
Eubacteriales sp. HG3A.0154
Eubacteriales sp. HG3A.0331
Eubacteriales sp. HG3A.0269
Eubacteriales sp. HG3A.0442
Eubacteriales sp. HG3A.0229
Eubacteriales sp. HG3A.0383
Firmicutes sp. HG3A.0397
Bacteria sp. HG3A.0483
Oscillibacter sp. HG3A.0245
Eubacteriales sp. HG3A.0381
Oscillospiraceae sp. HG3A.0437
Eubacteriales sp. HG3A.0136
Eubacteriales sp. HG3A.0409
Firmicutes sp. HG3A.0398
Eubacteriales sp. HG3A.0242
Oscillospiraceae sp. HG3A.0207
Eubacteriales sp. HG3A.0100
Intestinimonas massiliensis HG3A.0198
Oscillospiraceae sp. HG3A.0223
Anaerotruncus massiliensis HG3A.0460
Oscillospiraceae sp. HG3A.0507
Clostridia sp. HG3A.0756
Eubacteriales sp. HG3A.0697
Eubacteriales sp. HG3A.0859
Eubacteriales sp. HG3A.0514
Eubacteriales sp. HG3A.0450
Eubacteriales sp. HG3A.0369
Bacteria sp. HG3A.0638
Ruminococcus champanellensis HG3A.0716
Eubacteriales sp. HG3A.0751
Eubacteriales sp. HG3A.0609
Eubacteriales sp. HG3A.1445
Clostridia sp. HG3A.0750
Oscillospiraceae sp. HG3A.0210
Lachnospiraceae sp. HG3A.0217
Eubacteriales sp. HG3A.0443
Clostridia sp. HG3A.1053
Clostridia sp. HG3A.0385
Alistipes communis HG3A.0064
Alistipes shahii HG3A.0054
Odoribacter splanchnicus HG3A.0041
Eubacteriales sp. HG3A.0244
Allisonella histaminiformans HG3A.0332
Prevotella copri HG3A.0237
Senegalimassilia faecalis HG3A.0467
Ruminococcus sp. HG3A.0487
Eubacteriales sp. HG3A.0636
Oscillospiraceae sp. HG3A.0130
Clostridium phoceensis HG3A.0014
Dorea sp. AF24−7LB HG3A.0086
Pediococcus acidilactici HG3A.1468
Dorea longicatena HG3A.0039
Blautia sp. SG−772 HG3A.0063
Ruminococcus torques HG3A.0034
Coprococcus comes HG3A.0016
Collinsella aerofaciens HG3A.0019
Dorea formicigenerans HG3A.0006
Dorea sp. AF36−15AT HG3A.0052
Clostridium sp. TM06−18 HG3A.0048
Clostridium sp. TF06−15AC HG3A.0032
Anaerobutyricum hallii HG3A.0012
Blautia massiliensis HG3A.0023
Clostridium sp. HG3A.0050
Roseburia inulinivorans HG3A.0036
Lachnospiraceae sp. HG3A.0018
Butyricicoccus sp. HG3A.0008
Fusicatenibacter saccharivorans HG3A.0004
Coprobacillus sp. HG3A.0022
Anaerostipes hadrus HG3A.0003
Blautia obeum HG3A.0001
Variability of 24−hour SBP
Variability of 24−hour DBP
5alpha−androstan−3beta,17alpha−diol disulfate
3−phenylpropionate (hydrocinnamate)
cinnamoylglycine
branched chain 14:0 dicarboxylic acid**
phenylacetylglutamine
phenylacetate
5−hydroxyhexanoate
p−cresol sulfate
p−cresol glucuronide*
4−hydroxycoumarin
carotene diol (2)
beta−cryptoxanthin
Urolithin B glucuronide
4−methylcatechol sulfate
4−methylguaiacol sulfate
propionylglycine
3−hydroxypyridine sulfate
cholic acid glucuronide
cholate
hyocholate
1,3−dimethylurate
quinate
trigonelline (N'−methylnicotinate)
propionylcarnitine (C3)
tyramine O−sulfate
phenol sulfate
deoxycholate
deoxycholic acid 12−sulfate*
2−piperidinone
N−acetyl−cadaverine
ursodeoxycholate
isoursodeoxycholate
Variability of 24−hour SBP
0.4
0.2
0
−0.2
−0.4
Variability of 24−hour DBP
0.4
0.2
0
−0.2
−0.4
Spearman's rho
0.5
0
−0.5
Metabolite subclass
Acetylated Peptides
Androgenic Steroids
Bacterial/Fungal
Benzoate Metabolism
Chemical
Drug − Cardiovascular
Fatty Acid Metabolism (also BCAA Metabolism)
Fatty Acid, Dicarboxylate
Fatty Acid, Monohydroxy
Food Component/Plant
Lysine Metabolism
Nicotinate and Nicotinamide Metabolism
Phenylalanine Metabolism
Primary Bile Acid Metabolism
Secondary Bile Acid Metabolism
Tyrosine Metabolism
Vitamin A Metabolism
Xanthine Metabolism

#### Supplementary Tables

##### Table S1.

Associations between the metagenomic species (MGS) and 24-hour blood pressure outcomes from the 24-hour ABPM subsample in Model 1. Model 1: Adjusted for age, sex, country of birth, and technical source of variation. Name of MGS indicates the lowest taxonomic rank available to annotate the MGS; level of identification indicates the taxonomic rank; coef. indicates the regression coefficient of the linear model (a 1 SD increase in ln(relative abundance of MGS+1) implies a coef. point increase in the outcome variable); q-value is the adjusted *P*-value applying the Benjamini-Hochberg method on *P*-values from all four phenotypes together. SBP = systolic blood pressure; DBP = diastolic blood pressure; CI = confidence interval

##### Table S2.

Genera enriched for associations between metagenomic species (MGS) and 24-hour blood pressure outcomes in the 24-hour ABPM subsample in Model 1. Model 1 was adjusted for age, sex, country of birth, and technical source of variation. Taxon-set enrichment analysis was applied on the ranked *P*-values from Model 1 for positive and negative regression coefficients separately. 24-hour BP outcomes indicates the outcome variable in Model 1. Coef. indicates the normalized enrichment score; q-value is the adjusted enrichment *P*-value applying the Benjamini-Hochberg method on *P*-values from all four phenotypes together; and size indicates the number of species belonging to that genus in the analysis. DBP = diastolic blood pressure; SBP = systolic blood pressure

#### Table S3.

Gut metabolic modules (GMM) enriched for associations between metagenomic species (MGS) and 24-hour blood pressure outcomes in the 24-hour ABPM subsample in Model 1. Model 1 was adjusted for age, sex, country of birth, and technical source of variation. Taxon-set enrichment analysis was applied on the ranked *P*-values from Model 1 for positive and negative regression coefficients separately. GMM level 1 and GMM level 2 are the lowest and the highest hierarchical classification, respectively. 24-hour BP outcomes indicates the outcome variable in Model 1. Coef. indicates the normalized enrichment score; q-value is the adjusted enrichment *P*-value applying the Benjamini-Hochberg method on *P*-values from all four phenotypes together; and size indicates the number of species belonging to that module in the analysis. DBP = diastolic blood pressure; SBP = systolic blood pressure

#### Table S4.

Associations between the 140 unique metagenomic species (MGS) and 24-hour blood pressure outcomes from the ABPM subsample in Model 2. Model 2: adjusted for Model 1 covariates and additionally for smoking, fiber intake, total energy intake, sodium intake, usage of antidiabetic drugs, and usage of antihyperlipidemic drugs. These MGS were also investigated for influential observations. Influential observations were investigated by removing the observation with a highest absolute dfbeta. Name of MGS indicates the lowest taxonomic rank available to annotate the MGS; level of identification indicates the taxonomic rank; coef. indicates the regression coefficient of the linear model (a 1 SD increase in ln(relative abundance of MGS+1) implies a coef. point increase in the outcome variable); q-value is the adjusted *P*-value applying the Benjamini-Hochberg method on *P*-values from all four phenotypes together. SBP = systolic blood pressure; DBP = diastolic blood pressure; CI = confidence interval

#### Table S5.

Associations between the 140 unique metagenomic species (MGS) and 24-hour blood pressure outcomes in the 24-hour ABPM subsample in Model 3. Model 3: Adjusted for Model 2 covariates and additionally for BMI. These MGS were also investigated for influential observations. Influential observations were investigated by removing the observation with a highest absolute dfbeta. Name of MGS indicates the lowest taxonomic rank available to annotate the MGS; level of identification indicates the taxonomic rank; coef. indicates the regression coefficient of the linear model (a 1 SD increase in ln(relative abundance of MGS+1) implies a coef. point increase in the outcome variable); q-value is the adjusted *P*-value applying the Benjamini-Hochberg method on *P*-values from all four phenotypes together. SBP = systolic blood pressure; DBP = diastolic blood pressure; CI = confidence interval

#### Table S6.

Associations between the metagenomic species (MGS) and 24-hour blood pressure outcomes from the 24-hour ABPM subsample. Participants were excluded if they used antibiotics within half year, if they were diagnosed inflammatory bowel diseases, and if they used proton pump inhibitors. The significant results from Model 2 are presented. Model 2: adjusted for Model 1 covariates and additionally for smoking, fiber intake, total energy intake, sodium intake, usage of antidiabetic drugs, and usage of antihyperlipidemic drugs. Name of MGS indicates the lowest taxonomic rank available to annotate the MGS; 24-hour BP outcome indicates the outcome variable in the regression; coef. indicates the regression coefficient of the linear model (a 1 SD increase in ln(relative abundance of MGS+1) implies a coef. point increase in the outcome variable); q-value is the adjusted enrichment *P*-value applying the Benjamini-Hochberg method on *P*-values from all four phenotypes together. SBP = systolic blood pressure; DBP = diastolic blood pressure

#### Table S7.

Associations between gut microbial species (MGS) and plasma metabolites. Data were downloaded from the GUTSY Atlas ( https://gutsyatlas.serve.scilifelab.se/), which is based on the same metagenomics data as the current study. Shown are Spearman's rank correlations with q-value <0.05. n indicates the sample size in the analysis
