## Supplementary Figure S6 for "The association between the gut microbiome and 24-hour blood pressure measurements in the SCAPIS study"

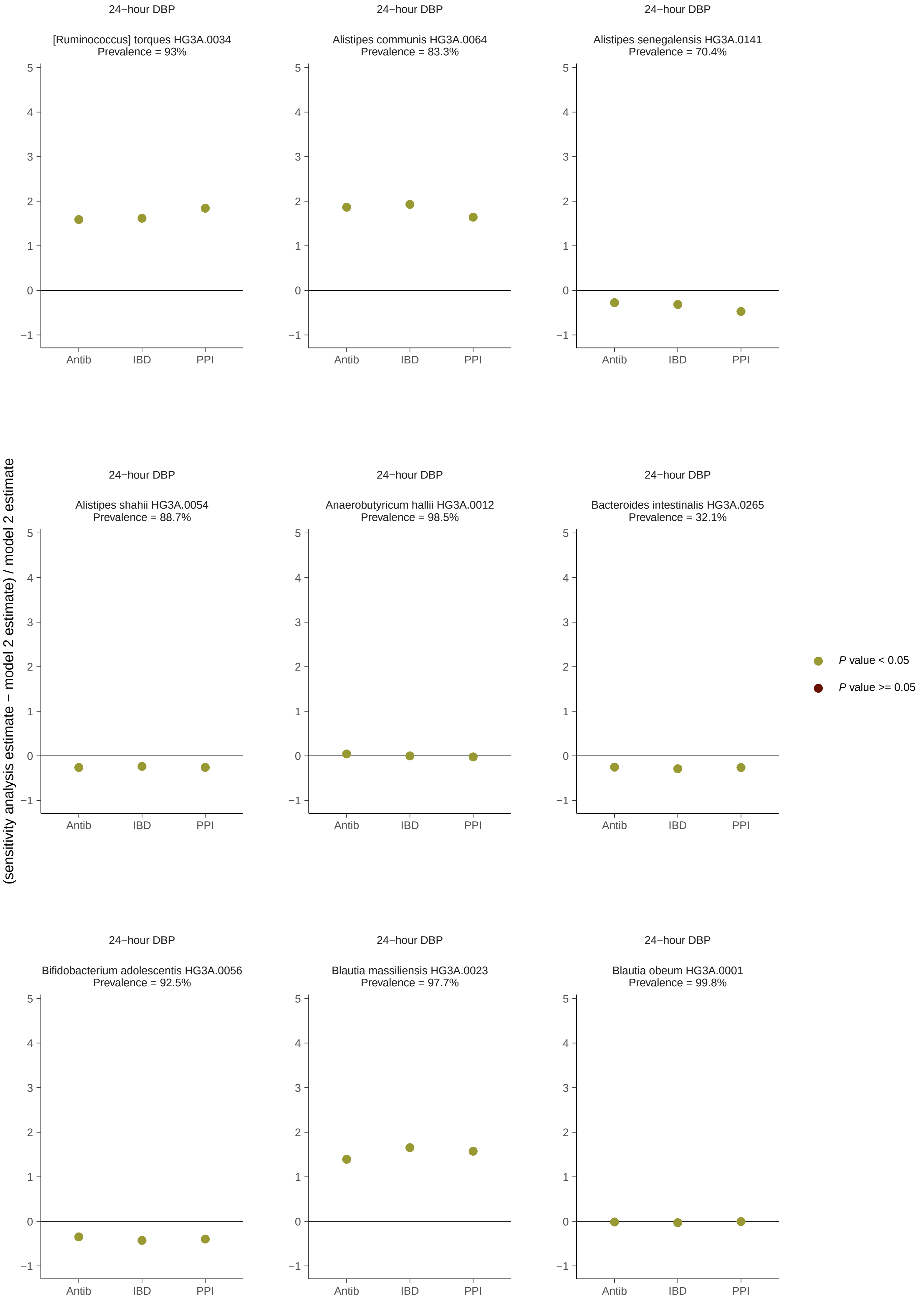
24−hour DBP
[Ruminococcus] torques HG3A.0034
Prevalence = 93%
24−hour DBP
Alistipes communis HG3A.0064
Prevalence = 83.3%
24−hour DBP
Alistipes senegalensis HG3A.0141
Prevalence = 70.4%
24−hour DBP
Alistipes shahii HG3A.0054
Prevalence = 88.7%
24−hour DBP
Anaerobutyricum hallii HG3A.0012
Prevalence = 98.5%
24−hour DBP
Bacteroides intestinalis HG3A.0265
Prevalence = 32.1%
24−hour DBP
Bifidobacterium adolescentis HG3A.0056
Prevalence = 92.5%
24−hour DBP
Blautia massiliensis HG3A.0023
Prevalence = 97.7%
24−hour DBP
Blautia obeum HG3A.0001
Prevalence = 99.8%
(sensitivity analysis estimate − model 2 estimate) / model 2 estimate
5
4
3
2
1
0
−1
Antib
IBD
PPI
P value < 0.05
P value >= 0.05

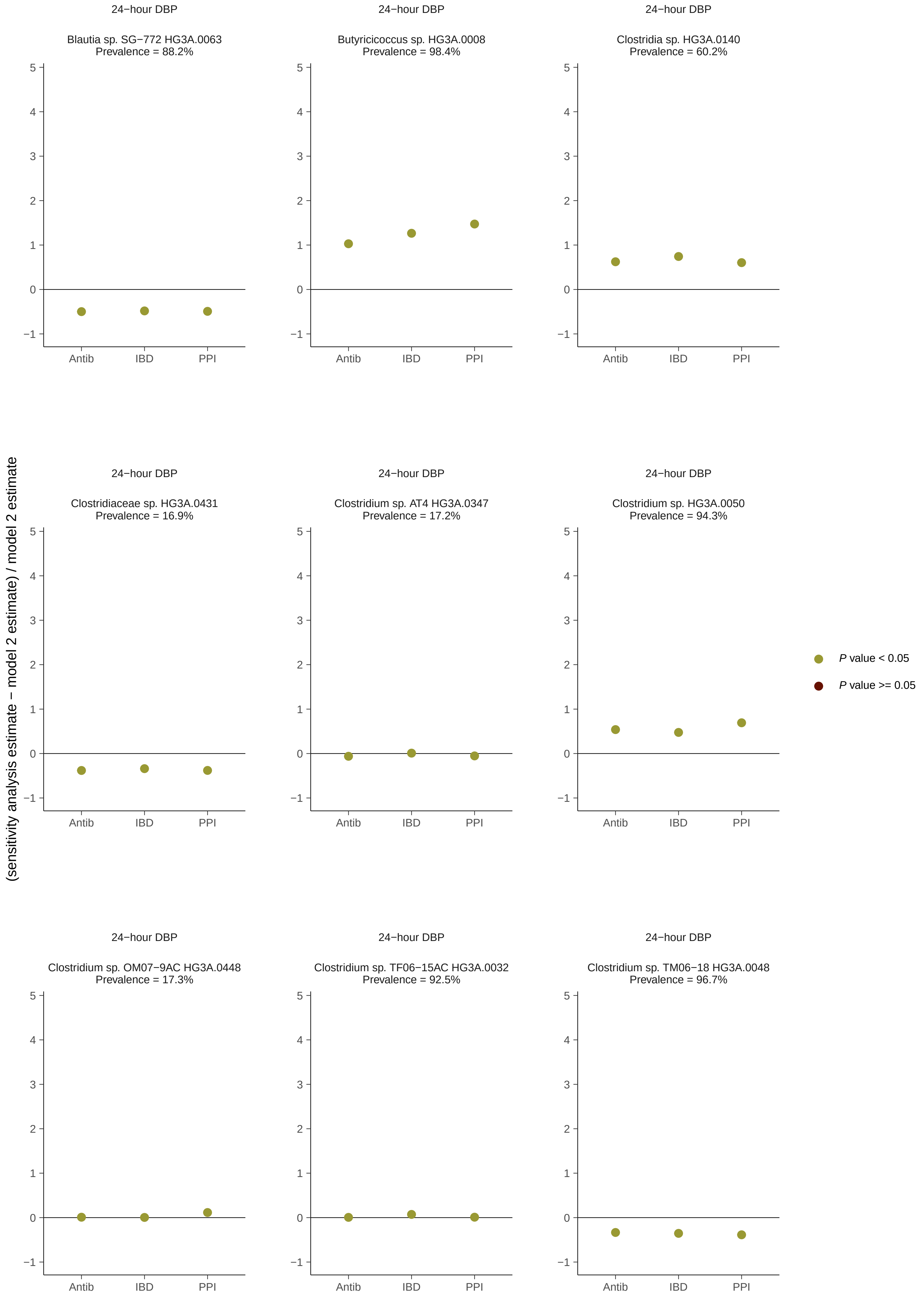

(sensitivity analysis estimate − model 2 estimate) / model 2 estimate
24−hour DBP
Blautia sp. SG−772 HG3A.0063
Prevalence = 88.2%
24−hour DBP
Butyricicoccus sp. HG3A.0008
Prevalence = 98.4%
24−hour DBP
Clostridia sp. HG3A.0140
Prevalence = 60.2%
24−hour DBP
Clostridiaceae sp. HG3A.0431
Prevalence = 16.9%
24−hour DBP
Clostridium sp. AT4 HG3A.0347
Prevalence = 17.2%
24−hour DBP
Clostridium sp. HG3A.0050
Prevalence = 94.3%
24−hour DBP
Clostridium sp. OM07−9AC HG3A.0448
Prevalence = 17.3%
24−hour DBP
Clostridium sp. TF06−15AC HG3A.0032
Prevalence = 92.5%
24−hour DBP
Clostridium sp. TM06−18 HG3A.0048
Prevalence = 96.7%
5
4
3
2
1
0
−1
Antib
IBD
PPI
P value < 0.05
P value >= 0.05

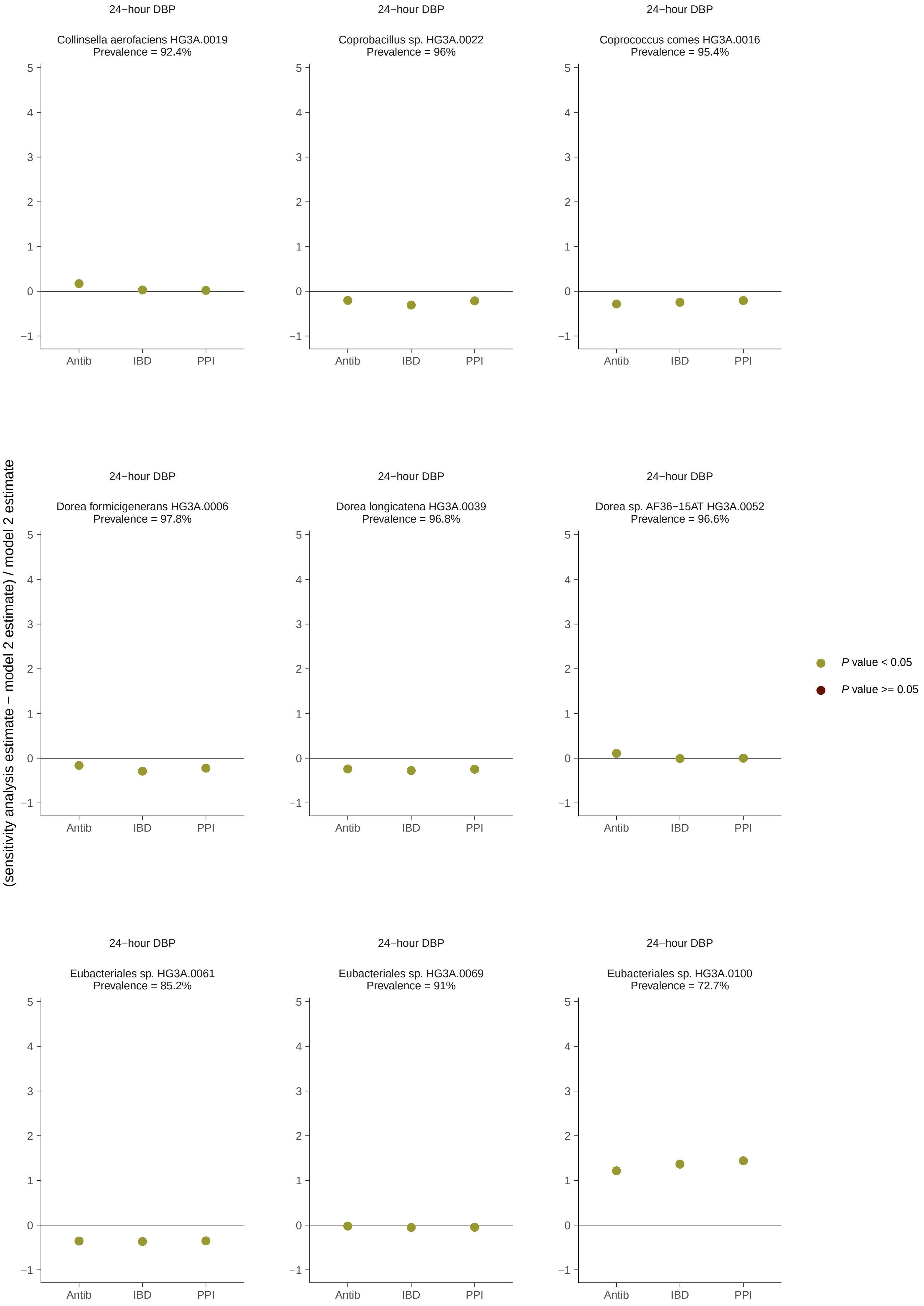

24−hour DBP
Collinsella aerofaciens HG3A.0019
Prevalence = 92.4%
24−hour DBP
Coprobacillus sp. HG3A.0022
Prevalence = 96%
24−hour DBP
Coprococcus comes HG3A.0016
Prevalence = 95.4%
24−hour DBP
Dorea formicigenerans HG3A.0006
Prevalence = 97.8%
24−hour DBP
Dorea longicatena HG3A.0039
Prevalence = 96.8%
24−hour DBP
Dorea sp. AF36−15AT HG3A.0052
Prevalence = 96.6%
24−hour DBP
Eubacteriales sp. HG3A.0061
Prevalence = 85.2%
24−hour DBP
Eubacteriales sp. HG3A.0069
Prevalence = 91%
24−hour DBP
Eubacteriales sp. HG3A.0100
Prevalence = 72.7%
Antib
IBD
PPI
(sensitivity analysis estimate − model 2 estimate) / model 2 estimate
P value < 0.05
P value >= 0.05

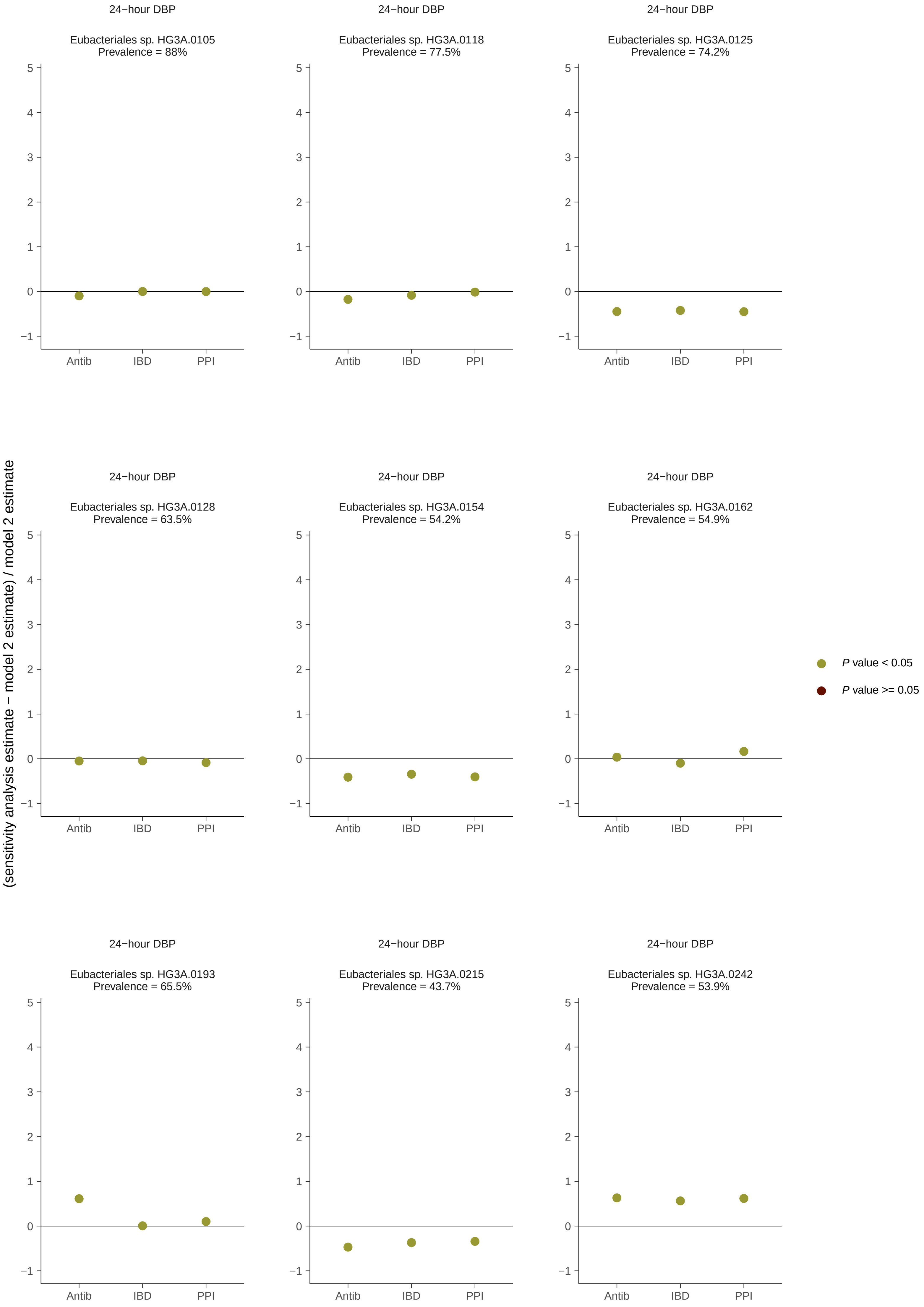
24−hour DBP
Eubacteriales sp. HG3A.0105
Prevalence = 88%
Eubacteriales sp. HG3A.0118
Prevalence = 77.5%
Eubacteriales sp. HG3A.0125
Prevalence = 74.2%
Eubacteriales sp. HG3A.0128
Prevalence = 63.5%
Eubacteriales sp. HG3A.0154
Prevalence = 54.2%
Eubacteriales sp. HG3A.0162
Prevalence = 54.9%
Eubacteriales sp. HG3A.0193
Prevalence = 65.5%
Eubacteriales sp. HG3A.0215
Prevalence = 43.7%
Eubacteriales sp. HG3A.0242
Prevalence = 53.9%
Antib
IBD
PPI
(sensitivity analysis estimate − model 2 estimate) / model 2 estimate
P value < 0.05
P value >= 0.05

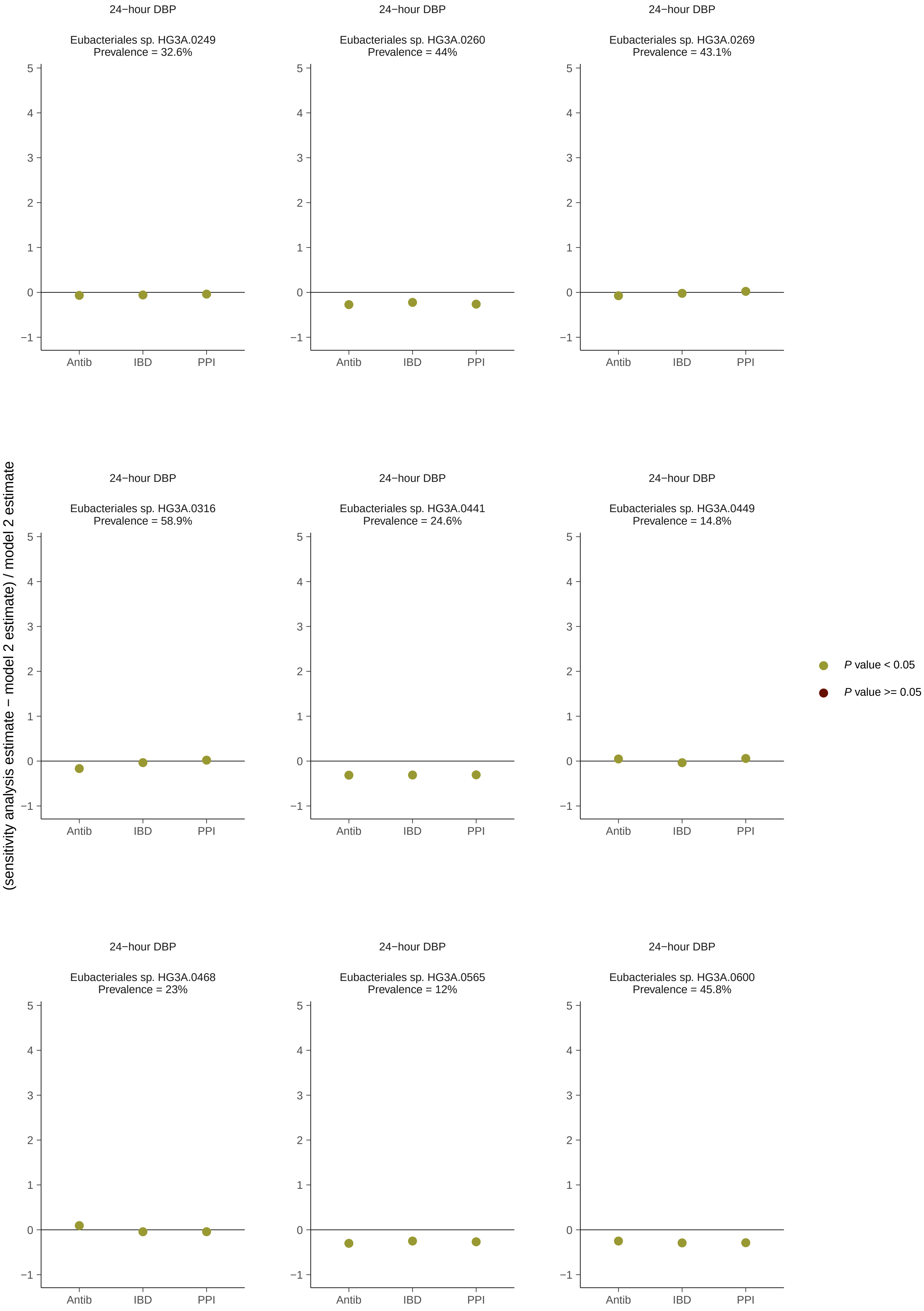

(sensitivity analysis estimate − model 2 estimate) / model 2 estimate
24−hour DBP
Eubacteriales sp. HG3A.0249
Prevalence = 32.6%
24−hour DBP
Eubacteriales sp. HG3A.0260
Prevalence = 44%
24−hour DBP
Eubacteriales sp. HG3A.0269
Prevalence = 43.1%
24−hour DBP
Eubacteriales sp. HG3A.0316
Prevalence = 58.9%
24−hour DBP
Eubacteriales sp. HG3A.0441
Prevalence = 24.6%
24−hour DBP
Eubacteriales sp. HG3A.0449
Prevalence = 14.8%
24−hour DBP
Eubacteriales sp. HG3A.0468
Prevalence = 23%
24−hour DBP
Eubacteriales sp. HG3A.0565
Prevalence = 12%
24−hour DBP
Eubacteriales sp. HG3A.0600
Prevalence = 45.8%
Antib
IBD
PPI
−1
0
1
2
3
4
5
P value < 0.05
P value >= 0.05

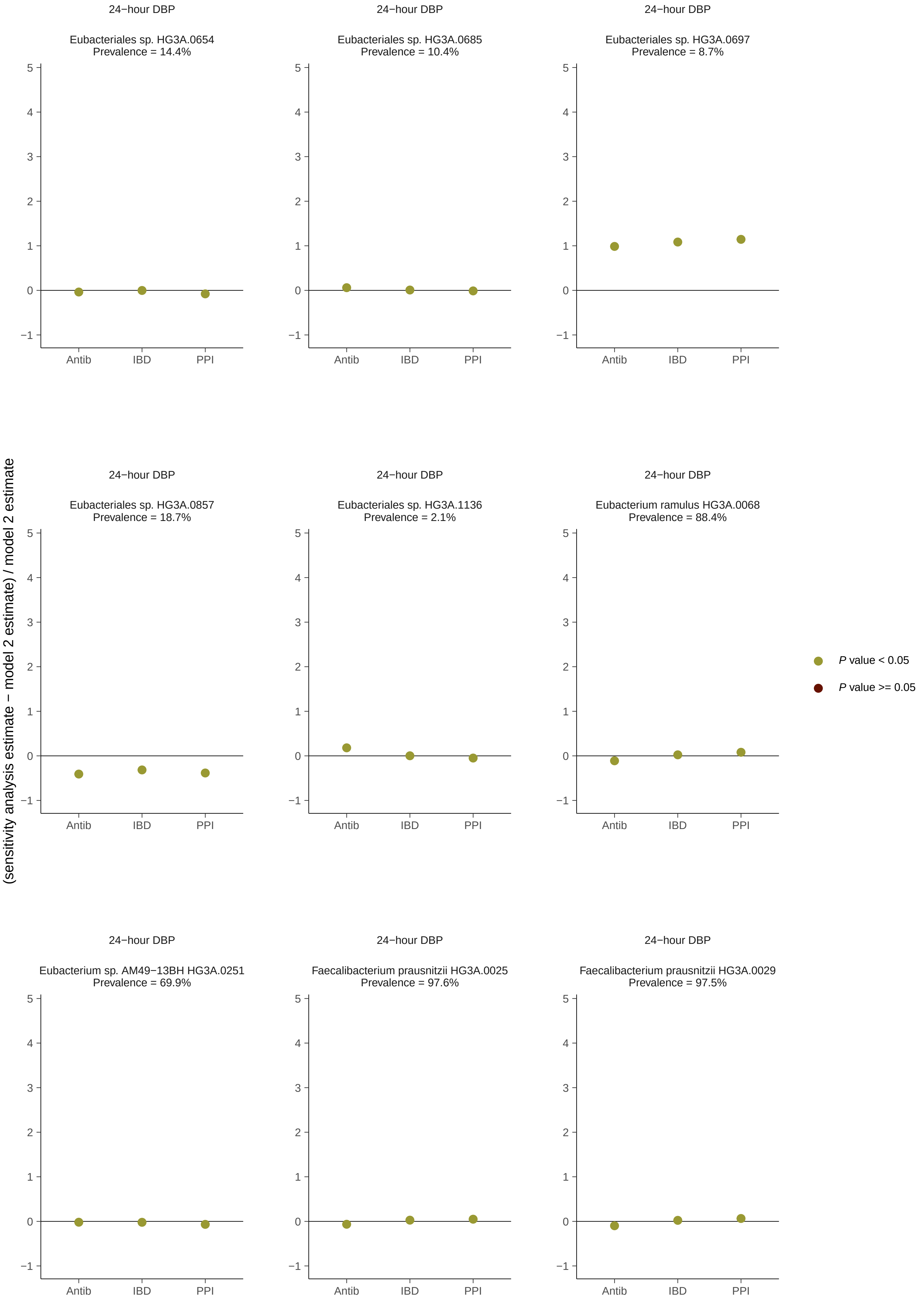
24−hour DBP
Eubacteriales sp. HG3A.0654
Prevalence = 14.4%
24−hour DBP
Eubacteriales sp. HG3A.0685
Prevalence = 10.4%
24−hour DBP
Eubacteriales sp. HG3A.0697
Prevalence = 8.7%
24−hour DBP
Eubacteriales sp. HG3A.0857
Prevalence = 18.7%
24−hour DBP
Eubacteriales sp. HG3A.1136
Prevalence = 2.1%
24−hour DBP
Eubacterium ramulus HG3A.0068
Prevalence = 88.4%
24−hour DBP
Eubacterium sp. AM49−13BH HG3A.0251
Prevalence = 69.9%
24−hour DBP
Faecalibacterium prausnitzii HG3A.0025
Prevalence = 97.6%
24−hour DBP
Faecalibacterium prausnitzii HG3A.0029
Prevalence = 97.5%
(sensitivity analysis estimate − model 2 estimate) / model 2 estimate
Antib
IBD
PPI
P value < 0.05
P value >= 0.05

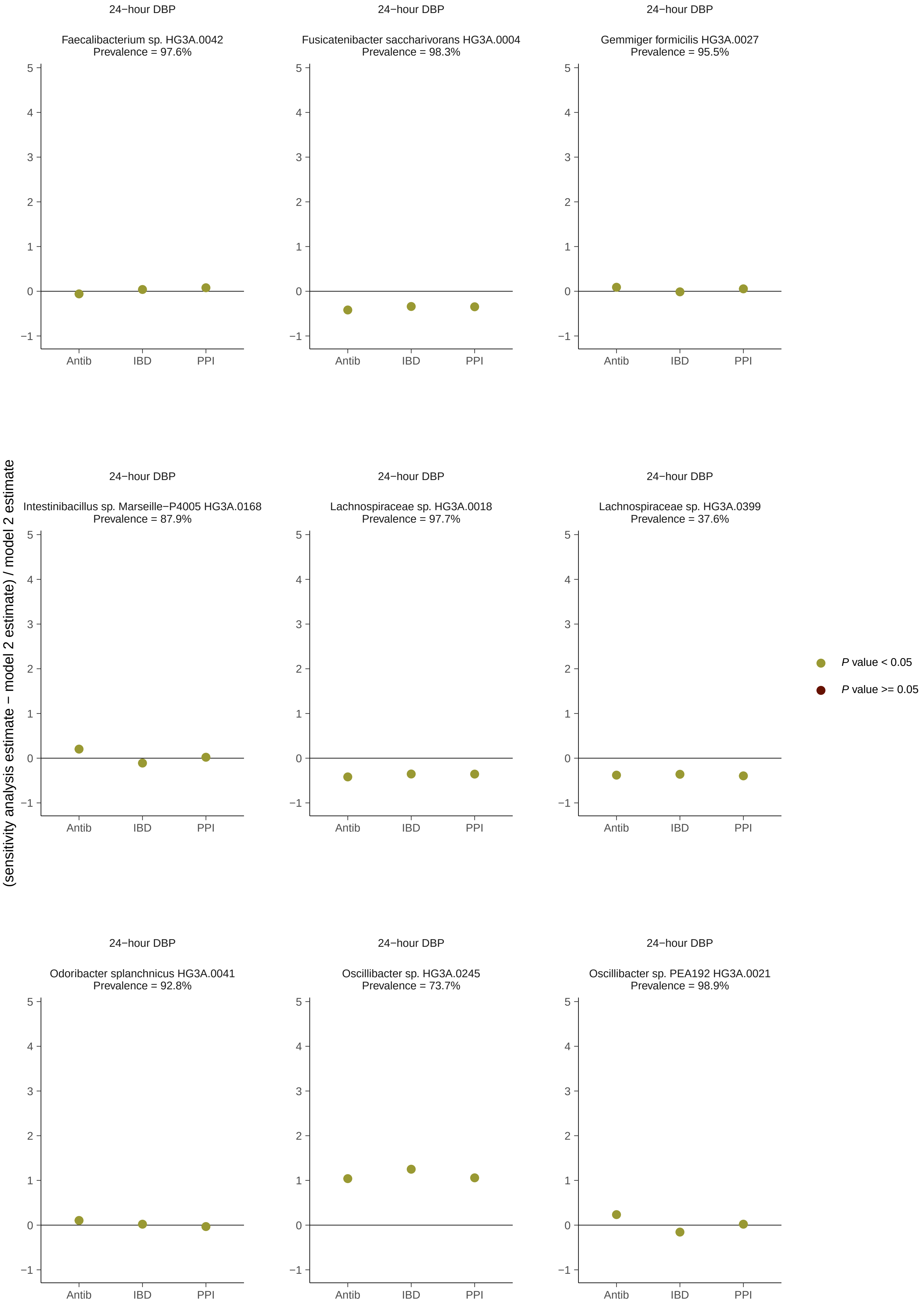
24−hour DBP
Faecalibacterium sp. HG3A.0042
Prevalence = 97.6%
24−hour DBP
Fusicatenibacter saccharivorans HG3A.0004
Prevalence = 98.3%
24−hour DBP
Gemmiger formicilis HG3A.0027
Prevalence = 95.5%
24−hour DBP
Intestinibacillus sp. Marseille−P4005 HG3A.0168
Prevalence = 87.9%
24−hour DBP
Lachnospiraceae sp. HG3A.0018
Prevalence = 97.7%
24−hour DBP
Lachnospiraceae sp. HG3A.0399
Prevalence = 37.6%
24−hour DBP
Odoribacter splanchnicus HG3A.0041
Prevalence = 92.8%
24−hour DBP
Oscillibacter sp. HG3A.0245
Prevalence = 73.7%
24−hour DBP
Oscillibacter sp. PEA192 HG3A.0021
Prevalence = 98.9%
(sensitivity analysis estimate − model 2 estimate) / model 2 estimate
−1
0
1
2
3
4
5
Antib
IBD
PPI
P value < 0.05
P value >= 0.05

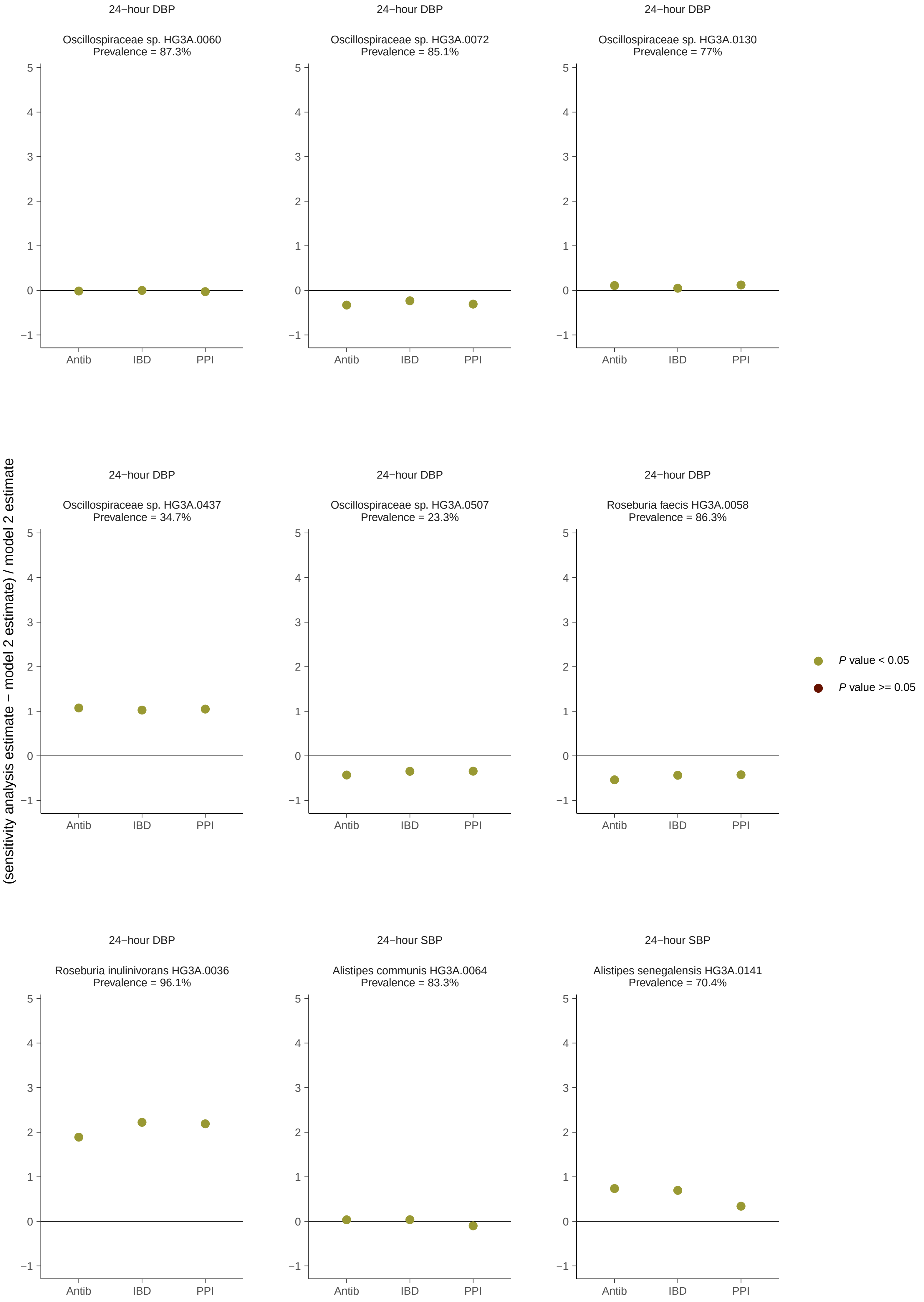
24−hour DBP
Oscillospiraceae sp. HG3A.0060
Prevalence = 87.3%
24−hour DBP
Oscillospiraceae sp. HG3A.0072
Prevalence = 85.1%
24−hour DBP
Oscillospiraceae sp. HG3A.0130
Prevalence = 77%
24−hour DBP
Oscillospiraceae sp. HG3A.0437
Prevalence = 34.7%
24−hour DBP
Oscillospiraceae sp. HG3A.0507
Prevalence = 23.3%
24−hour DBP
Roseburia faecis HG3A.0058
Prevalence = 86.3%
24−hour DBP
Roseburia inulinivorans HG3A.0036
Prevalence = 96.1%
24−hour SBP
Alistipes communis HG3A.0064
Prevalence = 83.3%
24−hour SBP
Alistipes senegalensis HG3A.0141
Prevalence = 70.4%
Antib
IBD
PPI
(sensitivity analysis estimate − model 2 estimate) / model 2 estimate
P value < 0.05
P value >= 0.05

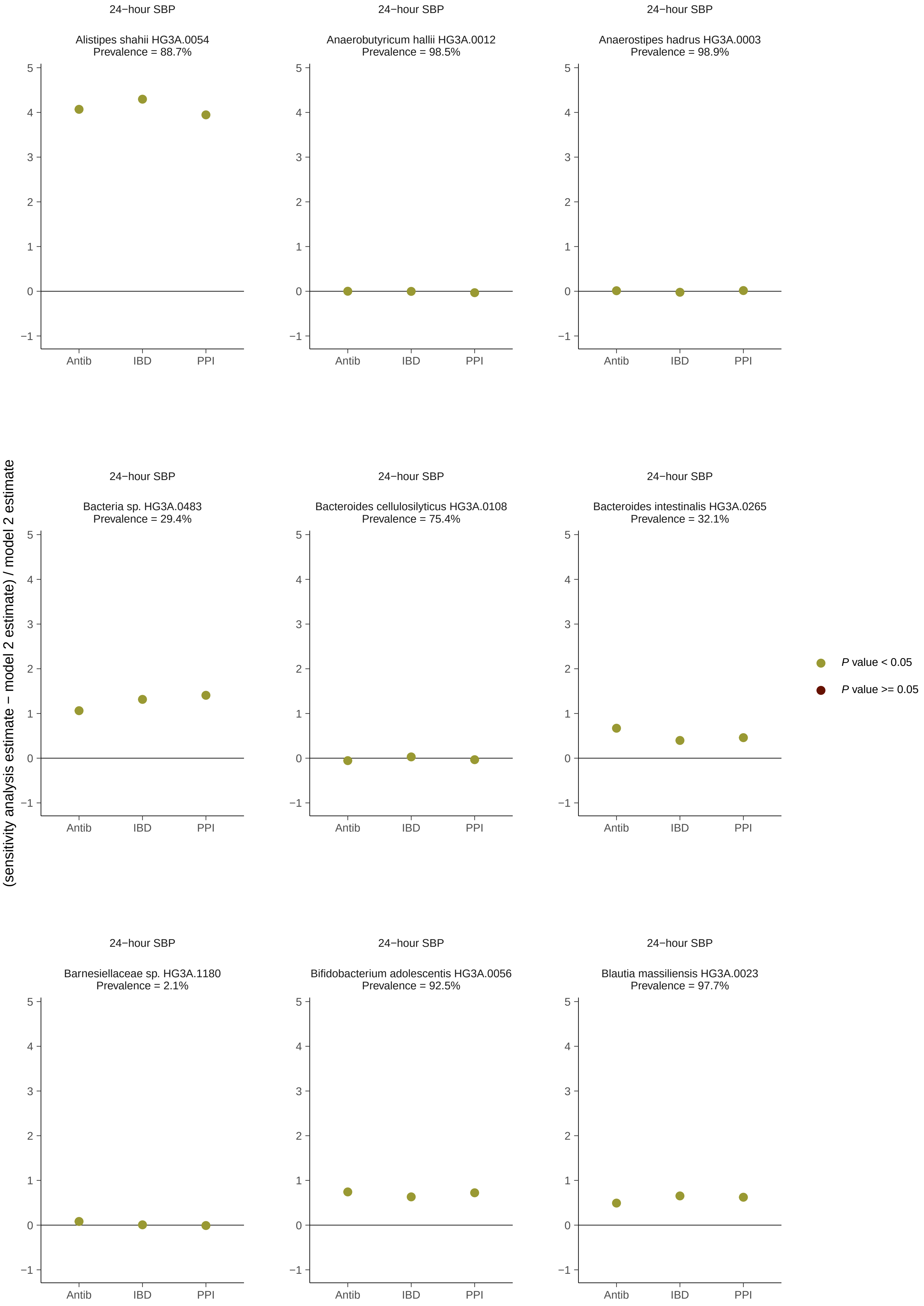
24−hour SBP
Alistipes shahii HG3A.0054
Prevalence = 88.7%
24−hour SBP
Anaerobutyricum hallii HG3A.0012
Prevalence = 98.5%
24−hour SBP
Anaerostipes hadrus HG3A.0003
Prevalence = 98.9%
24−hour SBP
Bacteria sp. HG3A.0483
Prevalence = 29.4%
24−hour SBP
Bacteroides cellulosilyticus HG3A.0108
Prevalence = 75.4%
24−hour SBP
Bacteroides intestinalis HG3A.0265
Prevalence = 32.1%
24−hour SBP
Barnesiellaceae sp. HG3A.1180
Prevalence = 2.1%
24−hour SBP
Bifidobacterium adolescentis HG3A.0056
Prevalence = 92.5%
24−hour SBP
Blautia massiliensis HG3A.0023
Prevalence = 97.7%
Antib
IBD
PPI
(sensitivity analysis estimate − model 2 estimate) / model 2 estimate
P value < 0.05
P value >= 0.05

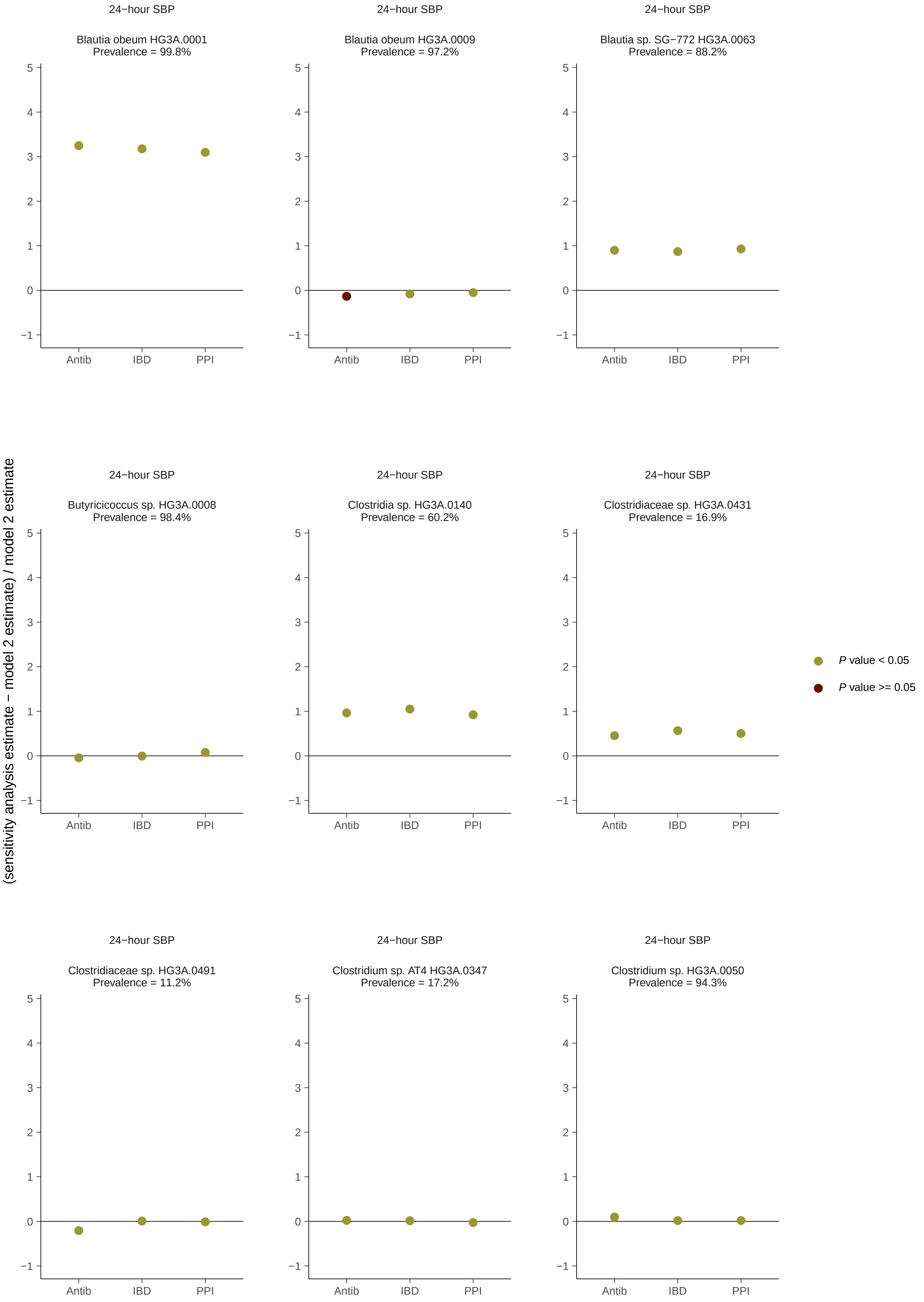
24−hour SBP
Blautia obeum HG3A.0001
Prevalence = 99.8%
24−hour SBP
Blautia obeum HG3A.0009
Prevalence = 97.2%
24−hour SBP
Blautia sp. SG−772 HG3A.0063
Prevalence = 88.2%
24−hour SBP
Butyricicoccus sp. HG3A.0008
Prevalence = 98.4%
24−hour SBP
Clostridia sp. HG3A.0140
Prevalence = 60.2%
24−hour SBP
Clostridiaceae sp. HG3A.0431
Prevalence = 16.9%
24−hour SBP
Clostridiaceae sp. HG3A.0491
Prevalence = 11.2%
24−hour SBP
Clostridium sp. AT4 HG3A.0347
Prevalence = 17.2%
24−hour SBP
Clostridium sp. HG3A.0050
Prevalence = 94.3%
Antib
IBD
PPI
(sensitivity analysis estimate − model 2 estimate) / model 2 estimate
P value < 0.05
P value >= 0.05

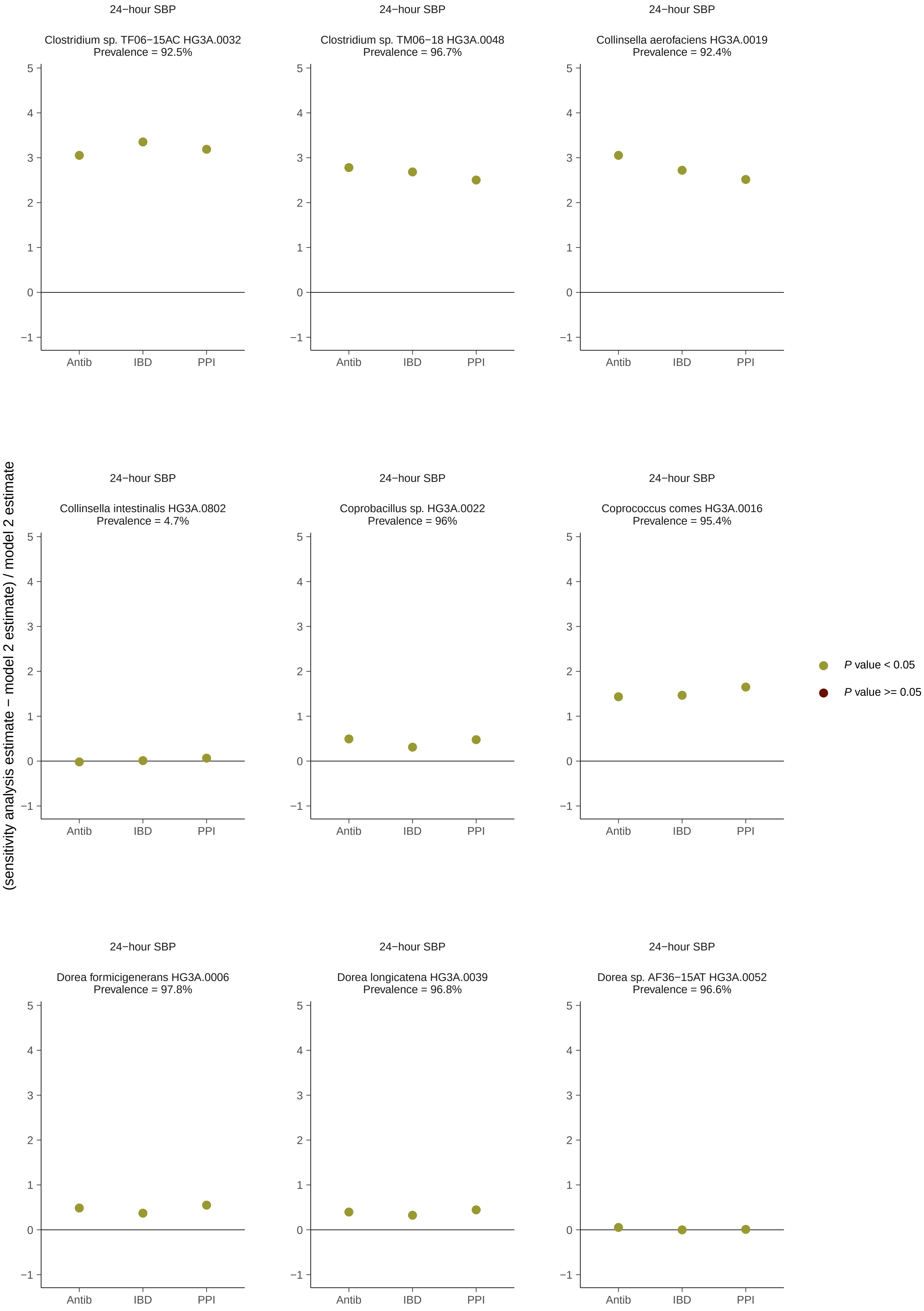
24−hour SBP
Clostridium sp. TF06−15AC HG3A.0032
Prevalence = 92.5%
24−hour SBP
Clostridium sp. TM06−18 HG3A.0048
Prevalence = 96.7%
24−hour SBP
Collinsella aerofaciens HG3A.0019
Prevalence = 92.4%
24−hour SBP
Collinsella intestinalis HG3A.0802
Prevalence = 4.7%
24−hour SBP
Coprobacillus sp. HG3A.0022
Prevalence = 96%
24−hour SBP
Coprococcus comes HG3A.0016
Prevalence = 95.4%
24−hour SBP
Dorea formicigenerans HG3A.0006
Prevalence = 97.8%
24−hour SBP
Dorea longicatena HG3A.0039
Prevalence = 96.8%
24−hour SBP
Dorea sp. AF36−15AT HG3A.0052
Prevalence = 96.6%
Antib
IBD
PPI
(sensitivity analysis estimate − model 2 estimate) / model 2 estimate
P value < 0.05
P value >= 0.05

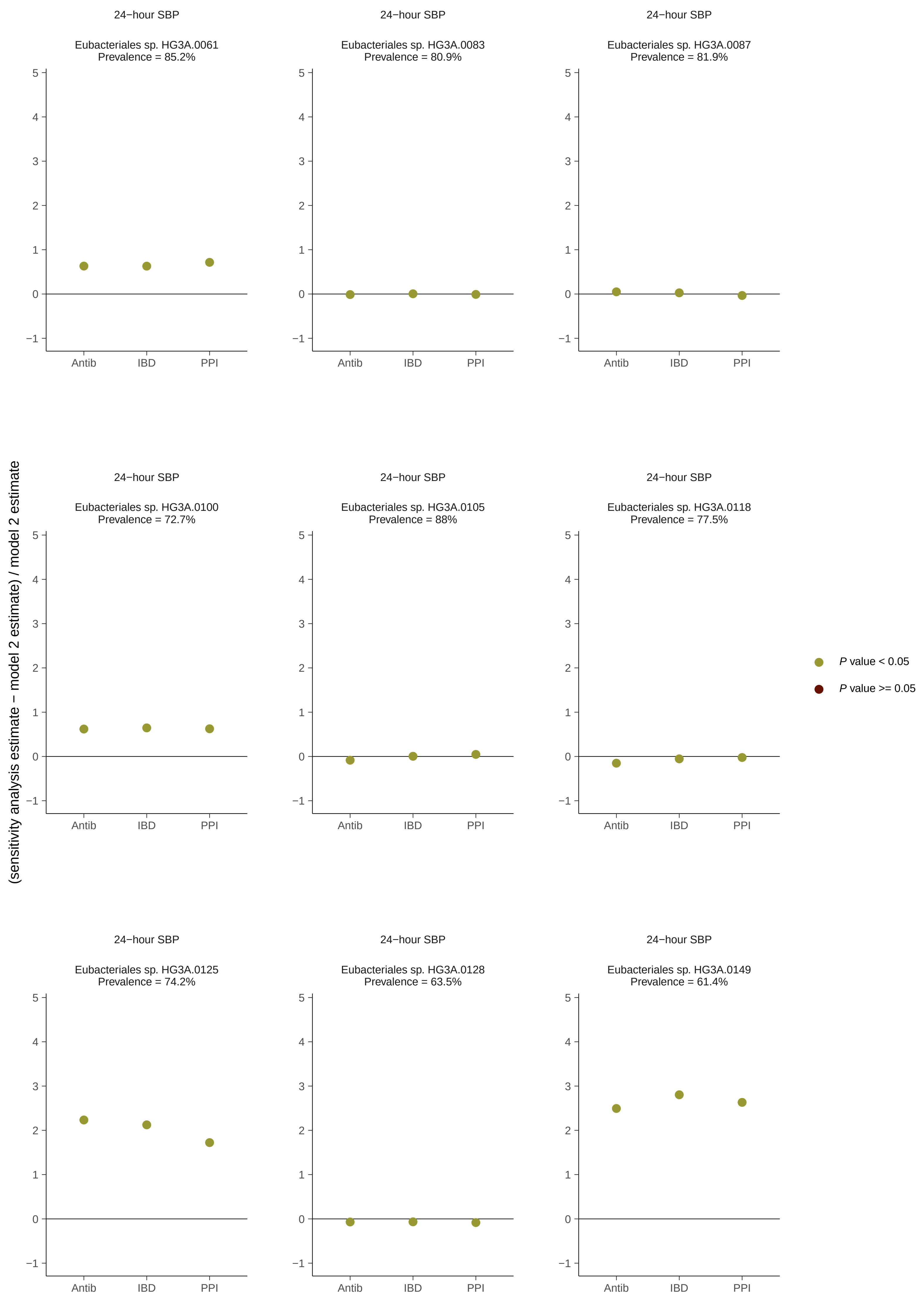
24−hour SBP
Eubacteriales sp. HG3A.0061
Prevalence = 85.2%
24−hour SBP
Eubacteriales sp. HG3A.0083
Prevalence = 80.9%
24−hour SBP
Eubacteriales sp. HG3A.0087
Prevalence = 81.9%
24−hour SBP
Eubacteriales sp. HG3A.0100
Prevalence = 72.7%
24−hour SBP
Eubacteriales sp. HG3A.0105
Prevalence = 88%
24−hour SBP
Eubacteriales sp. HG3A.0118
Prevalence = 77.5%
24−hour SBP
Eubacteriales sp. HG3A.0125
Prevalence = 74.2%
24−hour SBP
Eubacteriales sp. HG3A.0128
Prevalence = 63.5%
24−hour SBP
Eubacteriales sp. HG3A.0149
Prevalence = 61.4%
Antib
IBD
PPI
(sensitivity analysis estimate − model 2 estimate) / model 2 estimate
P value < 0.05
P value >= 0.05

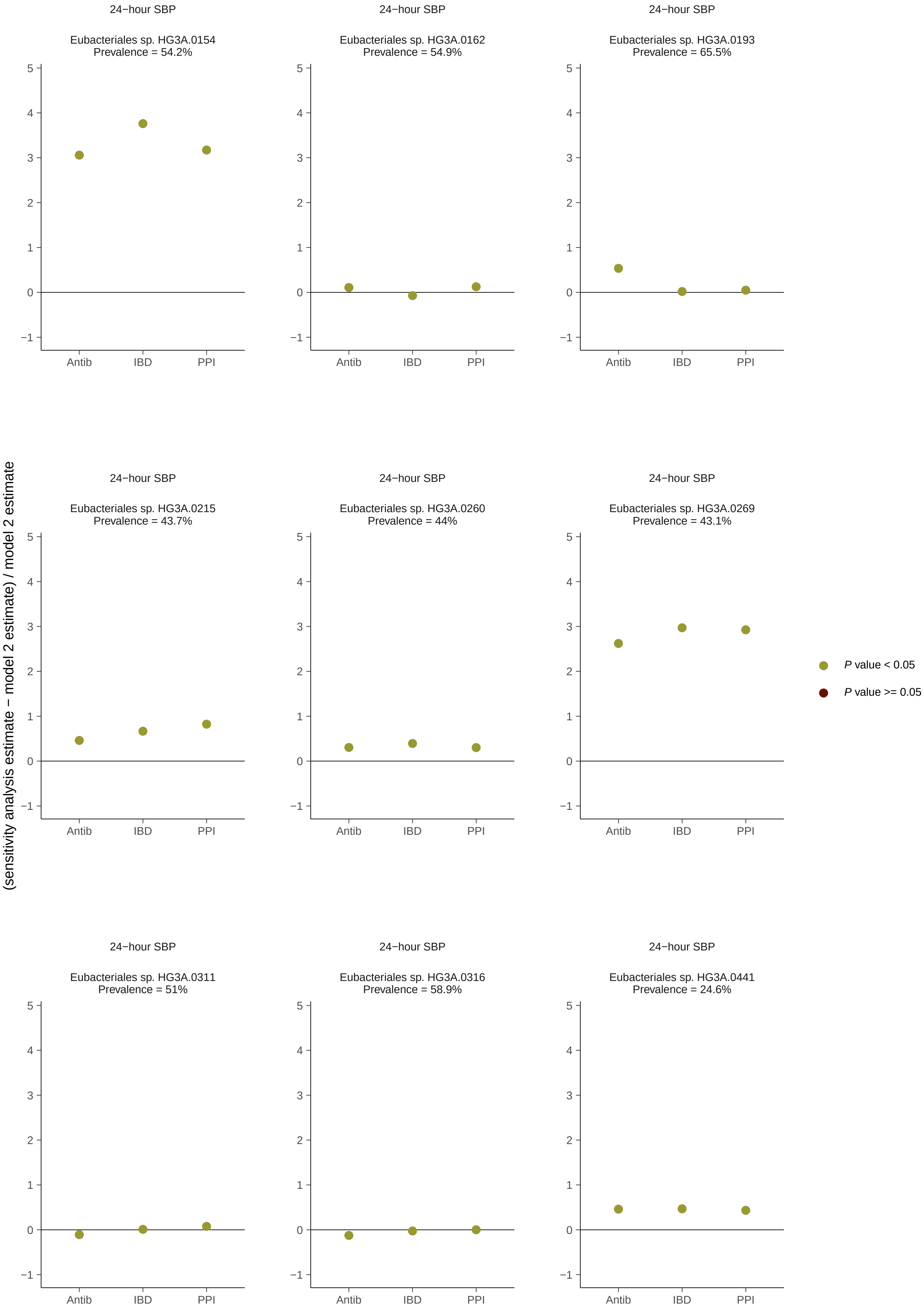
24−hour SBP
Eubacteriales sp. HG3A.0154
Prevalence = 54.2%
24−hour SBP
Eubacteriales sp. HG3A.0162
Prevalence = 54.9%
24−hour SBP
Eubacteriales sp. HG3A.0193
Prevalence = 65.5%
24−hour SBP
Eubacteriales sp. HG3A.0215
Prevalence = 43.7%
24−hour SBP
Eubacteriales sp. HG3A.0260
Prevalence = 44%
24−hour SBP
Eubacteriales sp. HG3A.0269
Prevalence = 43.1%
24−hour SBP
Eubacteriales sp. HG3A.0311
Prevalence = 51%
24−hour SBP
Eubacteriales sp. HG3A.0316
Prevalence = 58.9%
24−hour SBP
Eubacteriales sp. HG3A.0441
Prevalence = 24.6%
Antib
IBD
PPI
(sensitivity analysis estimate − model 2 estimate) / model 2 estimate
P value < 0.05
P value >= 0.05

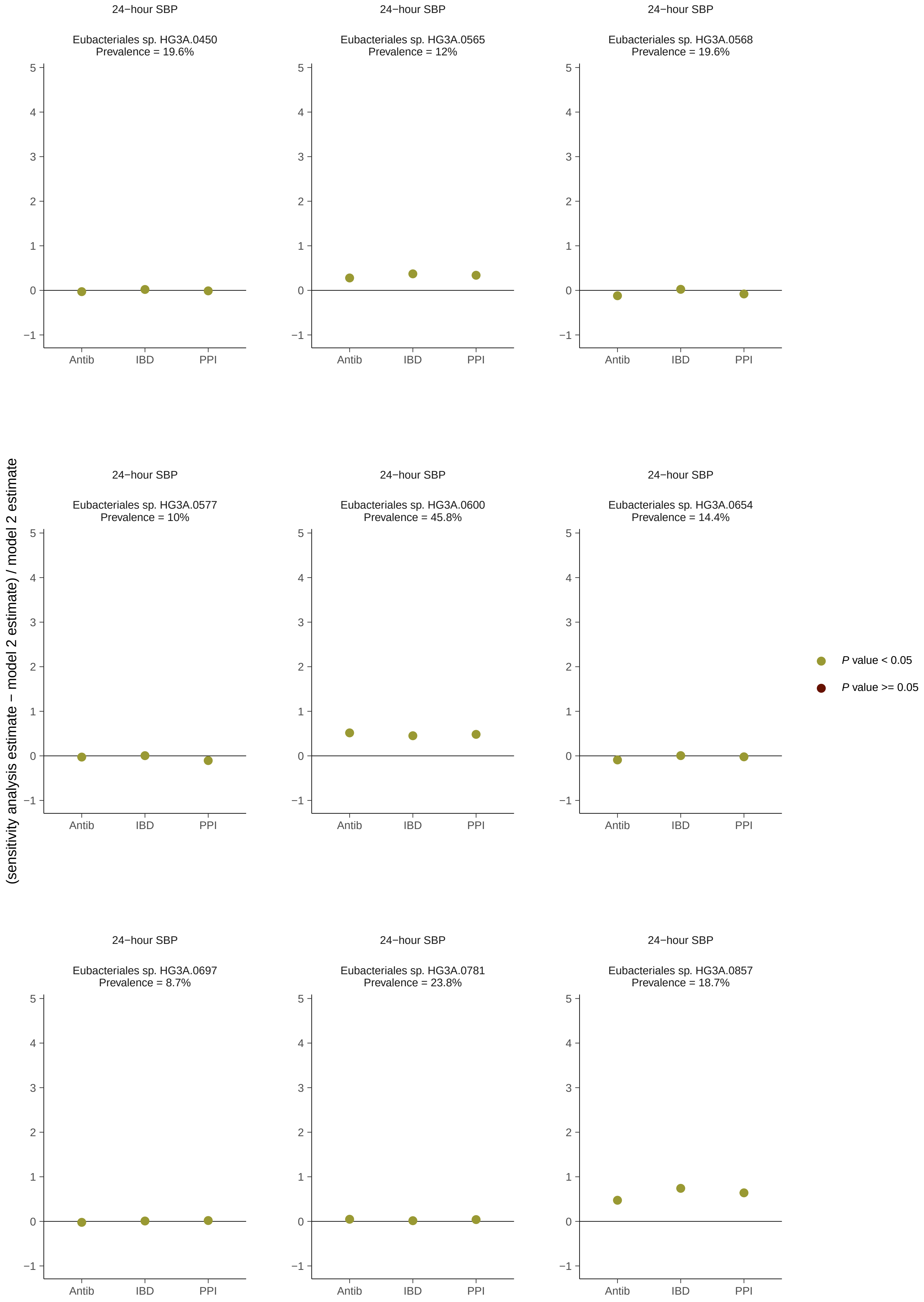
24−hour SBP
Eubacteriales sp. HG3A.0450
Prevalence = 19.6%
24−hour SBP
Eubacteriales sp. HG3A.0565
Prevalence = 12%
24−hour SBP
Eubacteriales sp. HG3A.0568
Prevalence = 19.6%
24−hour SBP
Eubacteriales sp. HG3A.0577
Prevalence = 10%
24−hour SBP
Eubacteriales sp. HG3A.0600
Prevalence = 45.8%
24−hour SBP
Eubacteriales sp. HG3A.0654
Prevalence = 14.4%
24−hour SBP
Eubacteriales sp. HG3A.0697
Prevalence = 8.7%
24−hour SBP
Eubacteriales sp. HG3A.0781
Prevalence = 23.8%
24−hour SBP
Eubacteriales sp. HG3A.0857
Prevalence = 18.7%
Antib
IBD
PPI
5
4
3
2
1
0
−1
(sensitivity analysis estimate − model 2 estimate) / model 2 estimate
P value < 0.05
P value >= 0.05

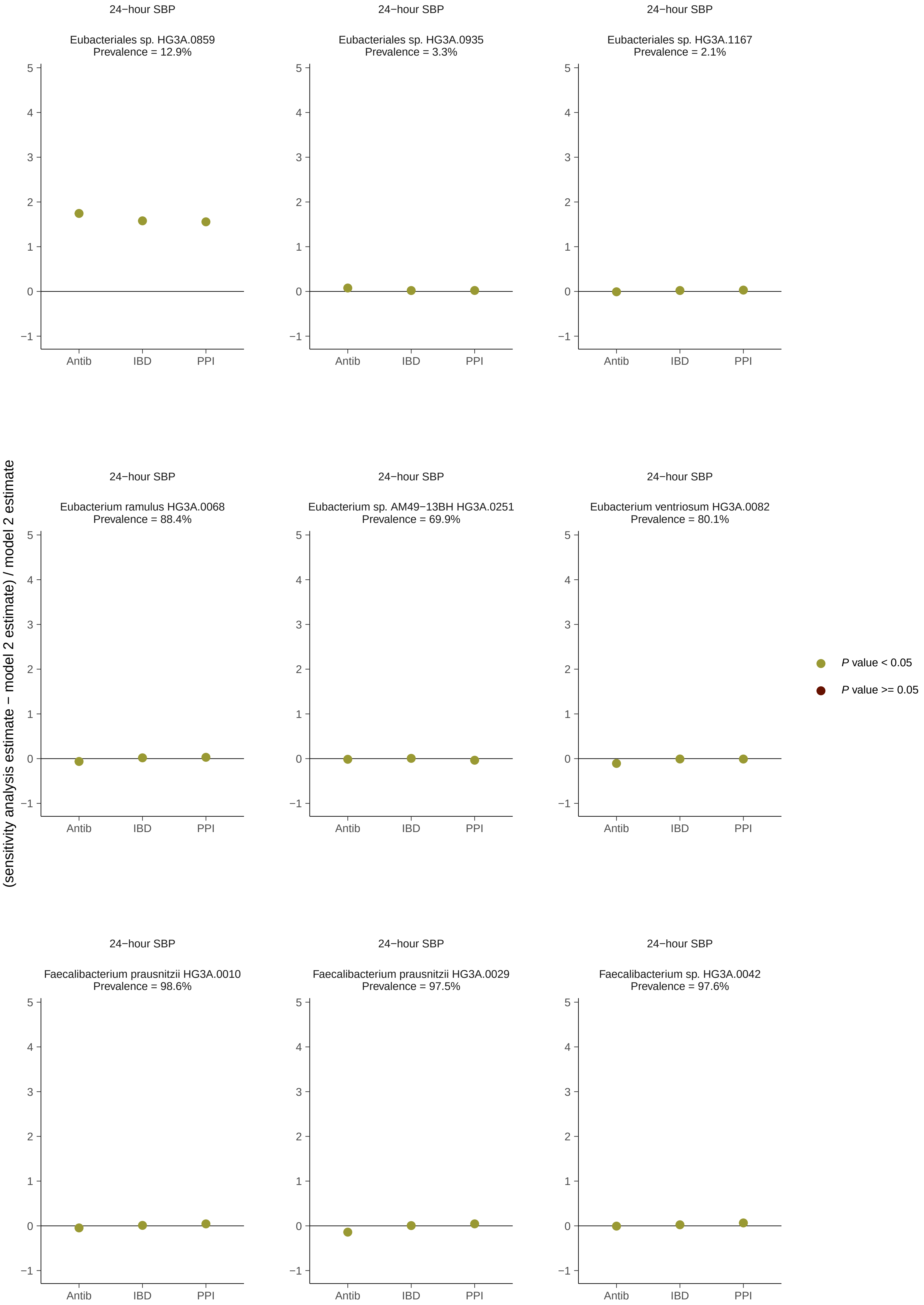
24−hour SBP
Eubacteriales sp. HG3A.0859
Prevalence = 12.9%
24−hour SBP
Eubacteriales sp. HG3A.0935
Prevalence = 3.3%
24−hour SBP
Eubacteriales sp. HG3A.1167
Prevalence = 2.1%
24−hour SBP
Eubacterium ramulus HG3A.0068
Prevalence = 88.4%
24−hour SBP
Eubacterium sp. AM49−13BH HG3A.0251
Prevalence = 69.9%
24−hour SBP
Eubacterium ventriosum HG3A.0082
Prevalence = 80.1%
24−hour SBP
Faecalibacterium prausnitzii HG3A.0010
Prevalence = 98.6%
24−hour SBP
Faecalibacterium prausnitzii HG3A.0029
Prevalence = 97.5%
24−hour SBP
Faecalibacterium sp. HG3A.0042
Prevalence = 97.6%
Antib
IBD
PPI
(sensitivity analysis estimate − model 2 estimate) / model 2 estimate
P value < 0.05
P value >= 0.05

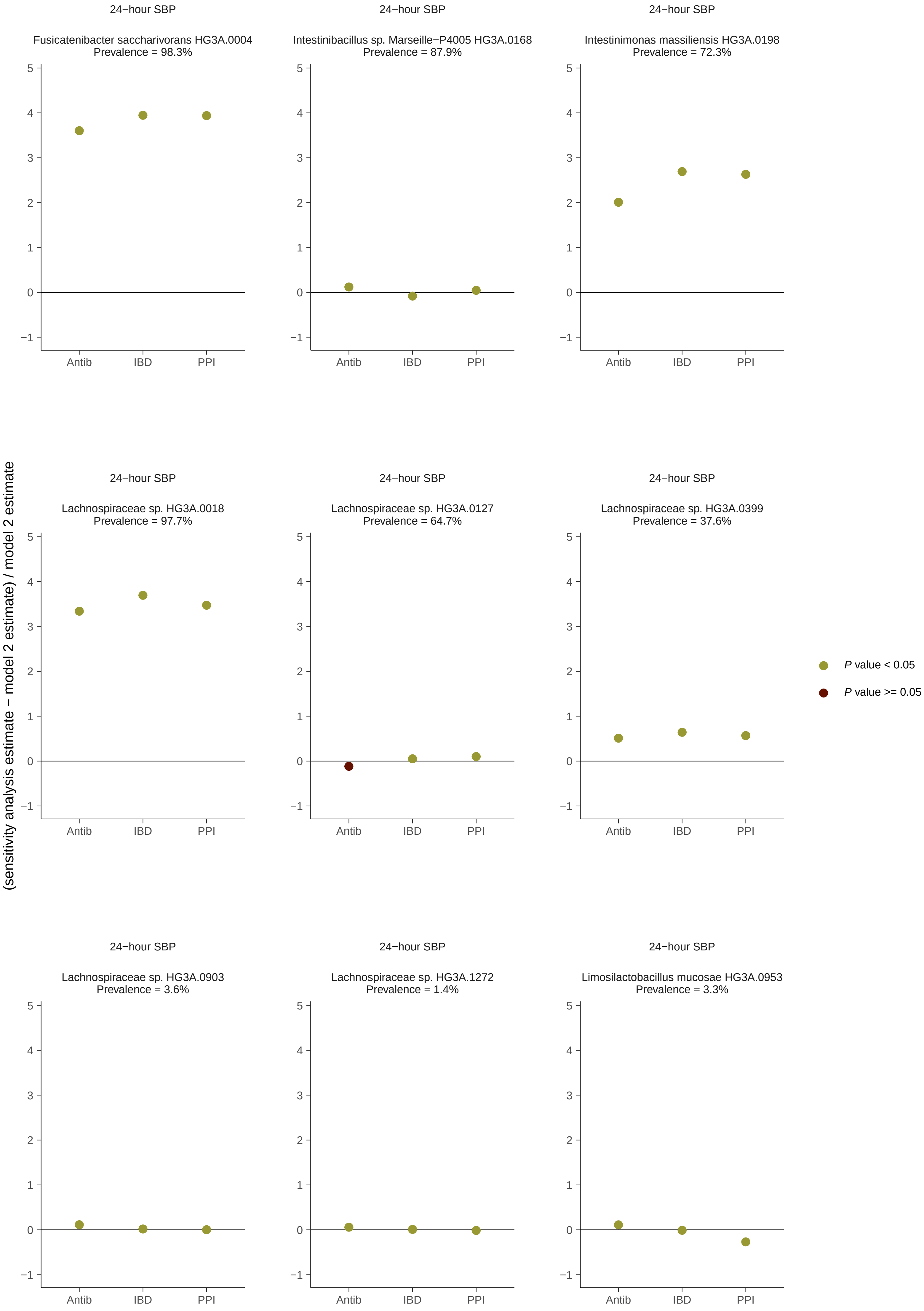
24−hour SBP
Fusicatenibacter saccharivorans HG3A.0004
Prevalence = 98.3%
24−hour SBP
Intestinibacillus sp. Marseille−P4005 HG3A.0168
Prevalence = 87.9%
24−hour SBP
Intestinimonas massiliensis HG3A.0198
Prevalence = 72.3%
24−hour SBP
Lachnospiraceae sp. HG3A.0018
Prevalence = 97.7%
24−hour SBP
Lachnospiraceae sp. HG3A.0127
Prevalence = 64.7%
24−hour SBP
Lachnospiraceae sp. HG3A.0399
Prevalence = 37.6%
24−hour SBP
Lachnospiraceae sp. HG3A.0903
Prevalence = 3.6%
24−hour SBP
Lachnospiraceae sp. HG3A.1272
Prevalence = 1.4%
24−hour SBP
Limosilactobacillus mucosae HG3A.0953
Prevalence = 3.3%
Antib
IBD
PPI
(sensitivity analysis estimate − model 2 estimate) / model 2 estimate
P value < 0.05
P value >= 0.05

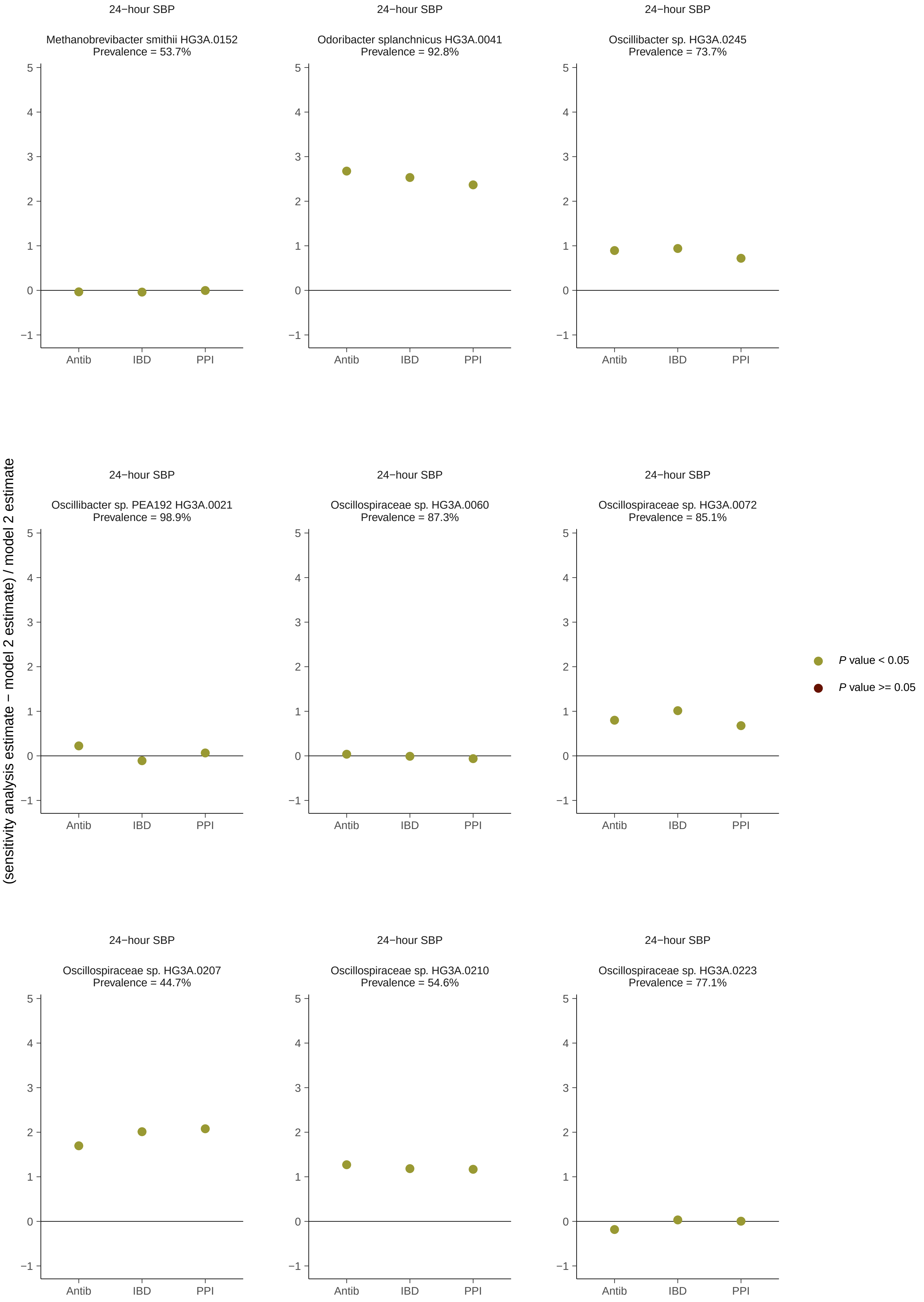
24−hour SBP
Methanobrevibacter smithii HG3A.0152
Prevalence = 53.7%
24−hour SBP
Odoribacter splanchnicus HG3A.0041
Prevalence = 92.8%
24−hour SBP
Oscillibacter sp. HG3A.0245
Prevalence = 73.7%
24−hour SBP
Oscillibacter sp. PEA192 HG3A.0021
Prevalence = 98.9%
24−hour SBP
Oscillospiraceae sp. HG3A.0060
Prevalence = 87.3%
24−hour SBP
Oscillospiraceae sp. HG3A.0072
Prevalence = 85.1%
24−hour SBP
Oscillospiraceae sp. HG3A.0207
Prevalence = 44.7%
24−hour SBP
Oscillospiraceae sp. HG3A.0210
Prevalence = 54.6%
24−hour SBP
Oscillospiraceae sp. HG3A.0223
Prevalence = 77.1%
Antib
IBD
PPI
(sensitivity analysis estimate − model 2 estimate) / model 2 estimate
P value < 0.05
P value >= 0.05

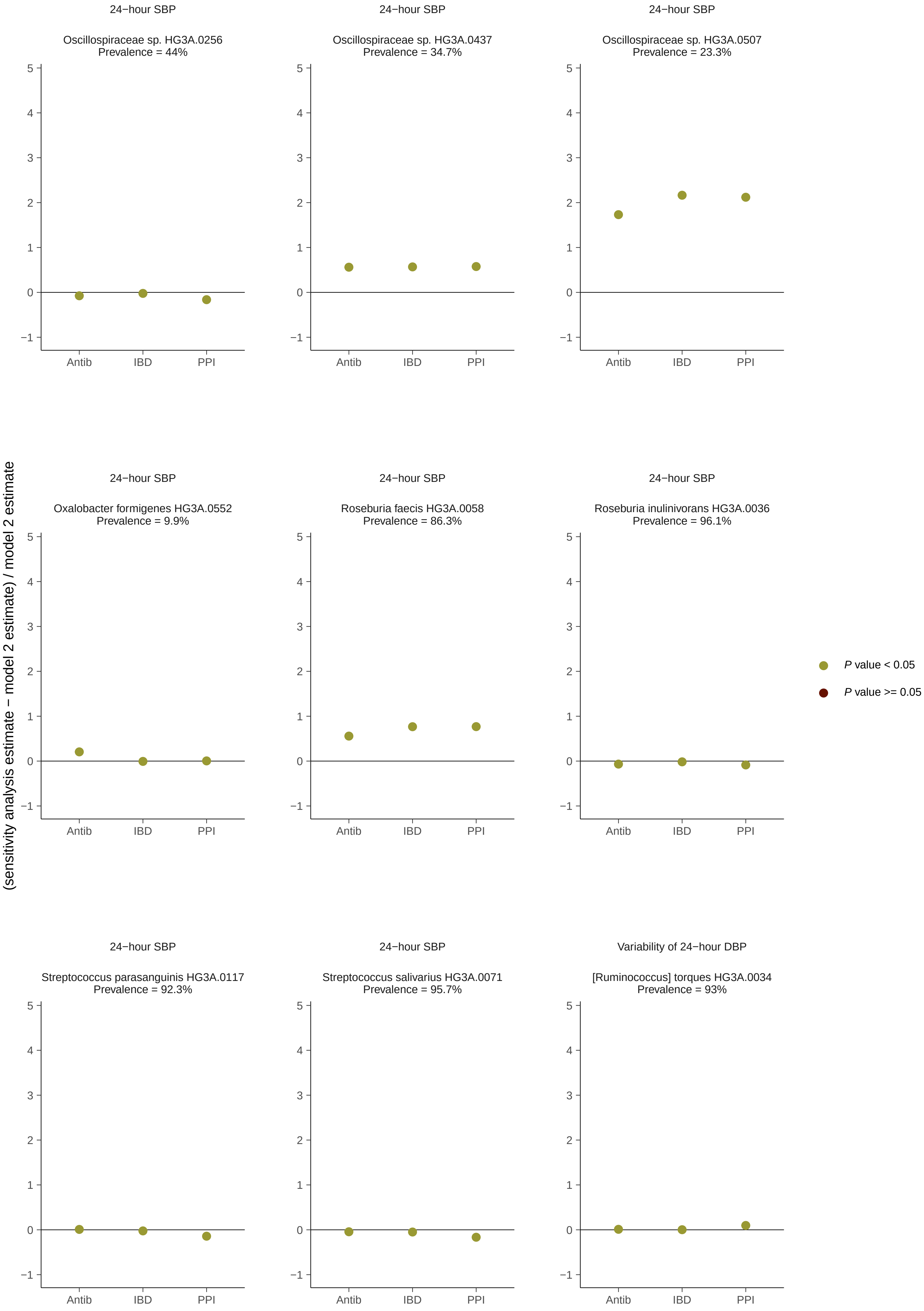
24−hour SBP
Oscillospiraceae sp. HG3A.0256
Prevalence = 44%
24−hour SBP
Oscillospiraceae sp. HG3A.0437
Prevalence = 34.7%
24−hour SBP
Oscillospiraceae sp. HG3A.0507
Prevalence = 23.3%
24−hour SBP
Oxalobacter formigenes HG3A.0552
Prevalence = 9.9%
24−hour SBP
Roseburia faecis HG3A.0058
Prevalence = 86.3%
24−hour SBP
Roseburia inulinivorans HG3A.0036
Prevalence = 96.1%
24−hour SBP
Streptococcus parasanguinis HG3A.0117
Prevalence = 92.3%
24−hour SBP
Streptococcus salivarius HG3A.0071
Prevalence = 95.7%
Variability of 24−hour DBP
[Ruminococcus] torques HG3A.0034
Prevalence = 93%
Antib
IBD
PPI
(sensitivity analysis estimate − model 2 estimate) / model 2 estimate
P value < 0.05
P value >= 0.05

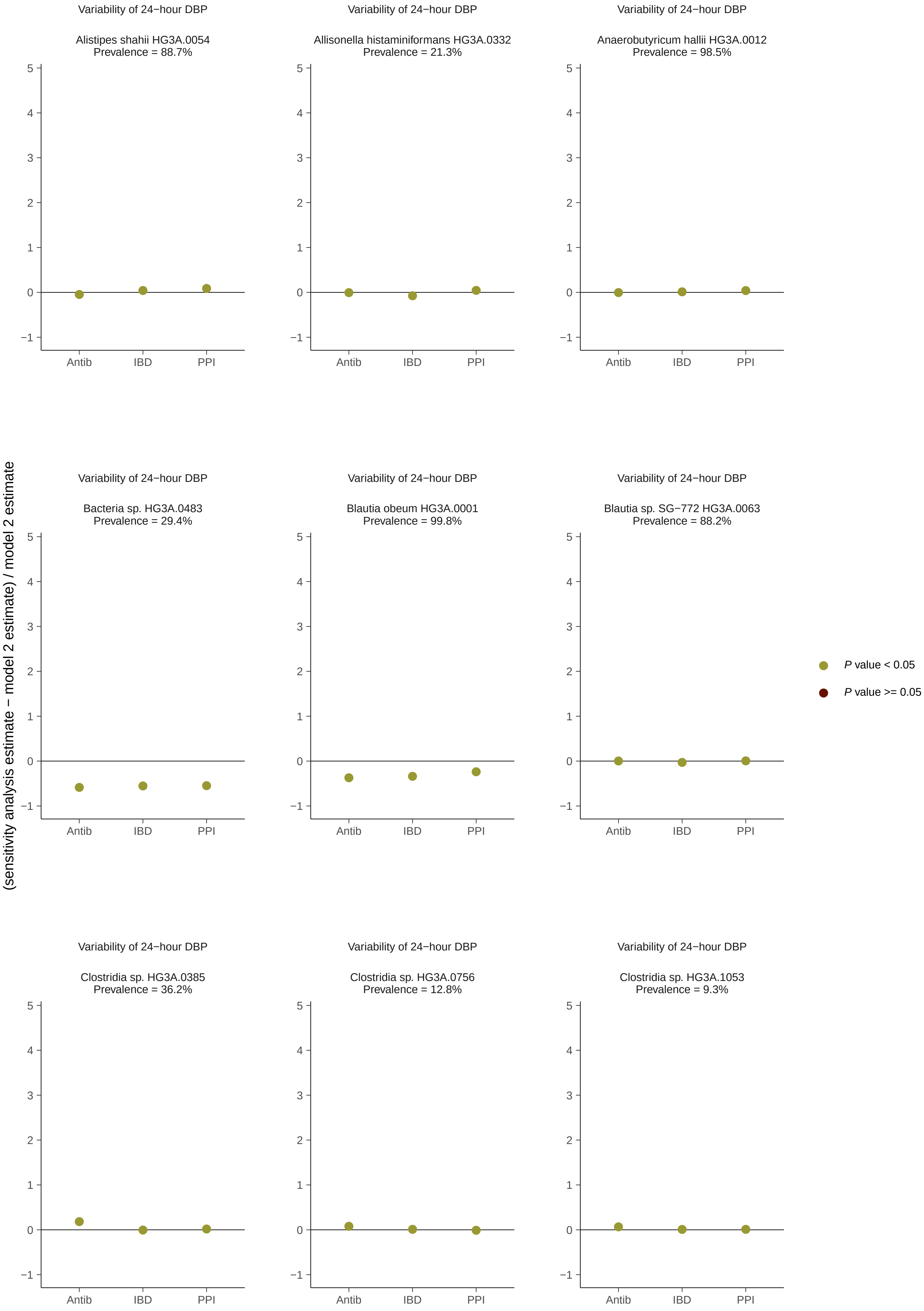

Variability of 24−hour DBP
Alistipes shahii HG3A.0054
Prevalence = 88.7%
Variability of 24−hour DBP
Allisonella histaminiformans HG3A.0332
Prevalence = 21.3%
Variability of 24−hour DBP
Anaerobutyricum hallii HG3A.0012
Prevalence = 98.5%
Variability of 24−hour DBP
Bacteria sp. HG3A.0483
Prevalence = 29.4%
Variability of 24−hour DBP
Blautia obeum HG3A.0001
Prevalence = 99.8%
Variability of 24−hour DBP
Blautia sp. SG−772 HG3A.0063
Prevalence = 88.2%
Variability of 24−hour DBP
Clostridia sp. HG3A.0385
Prevalence = 36.2%
Variability of 24−hour DBP
Clostridia sp. HG3A.0756
Prevalence = 12.8%
Variability of 24−hour DBP
Clostridia sp. HG3A.1053
Prevalence = 9.3%
Antib
IBD
PPI
5
4
3
2
1
0
−1
(sensitivity analysis estimate − model 2 estimate) / model 2 estimate
P value < 0.05
P value >= 0.05

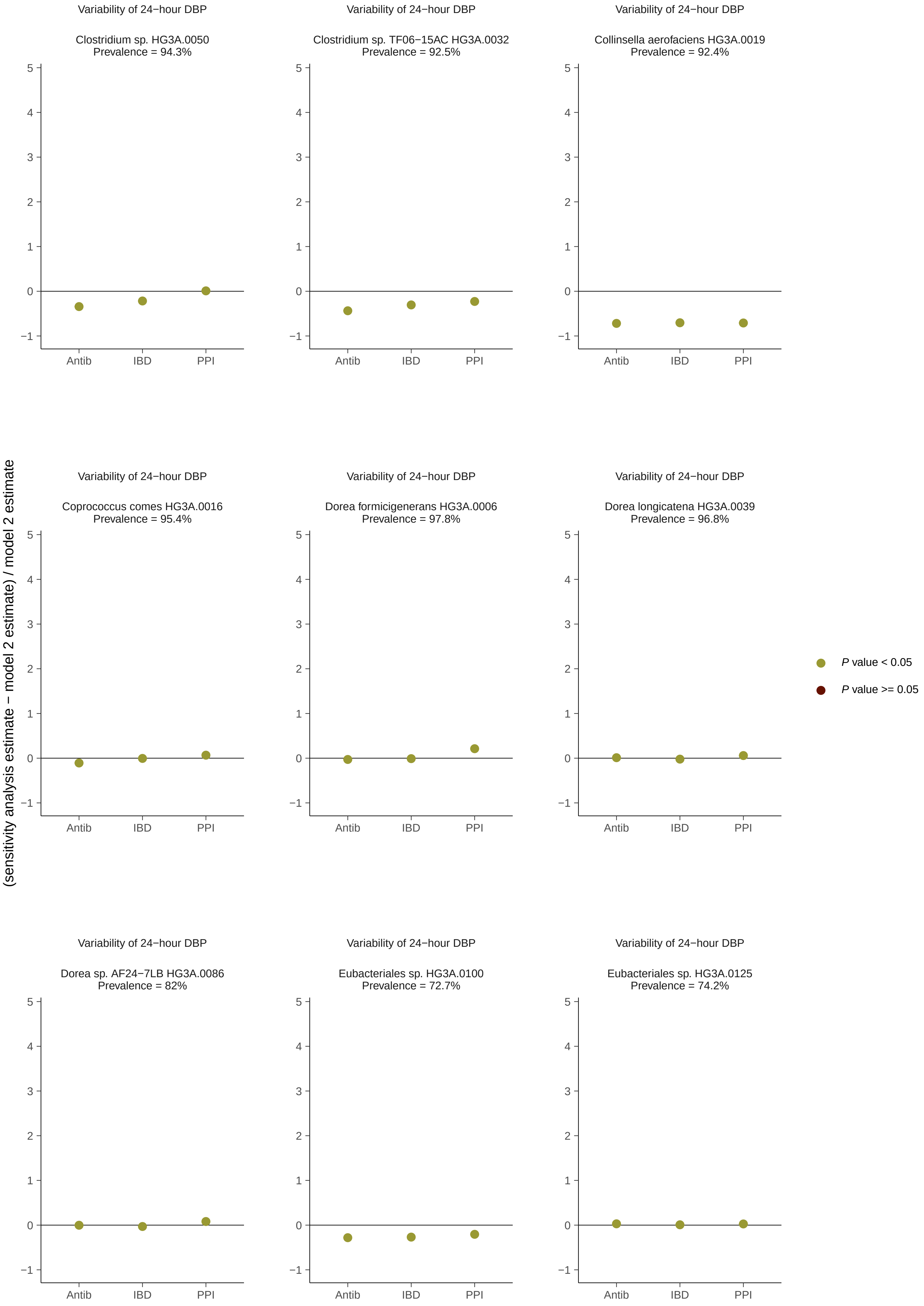
Variability of 24−hour DBP
Clostridium sp. HG3A.0050
Prevalence = 94.3%
Variability of 24−hour DBP
Clostridium sp. TF06−15AC HG3A.0032
Prevalence = 92.5%
Variability of 24−hour DBP
Collinsella aerofaciens HG3A.0019
Prevalence = 92.4%
Variability of 24−hour DBP
Coprococcus comes HG3A.0016
Prevalence = 95.4%
Variability of 24−hour DBP
Dorea formicigenerans HG3A.0006
Prevalence = 97.8%
Variability of 24−hour DBP
Dorea longicatena HG3A.0039
Prevalence = 96.8%
Variability of 24−hour DBP
Dorea sp. AF24−7LB HG3A.0086
Prevalence = 82%
Variability of 24−hour DBP
Eubacteriales sp. HG3A.0100
Prevalence = 72.7%
Variability of 24−hour DBP
Eubacteriales sp. HG3A.0125
Prevalence = 74.2%
5
4
3
2
1
0
−1
Antib
IBD
PPI
(sensitivity analysis estimate − model 2 estimate) / model 2 estimate
P value < 0.05
P value >= 0.05

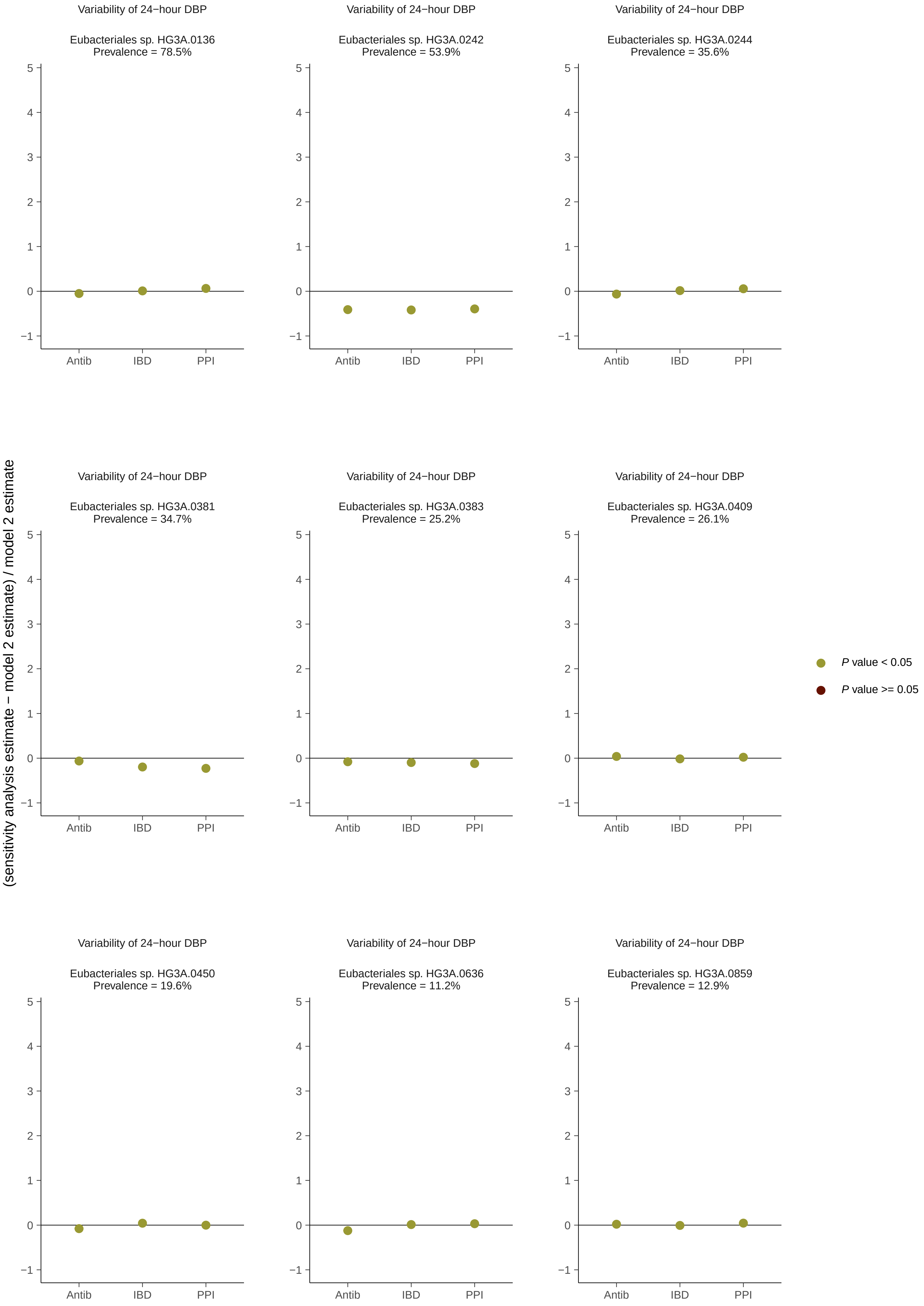
Variability of 24−hour DBP
Eubacteriales sp. HG3A.0136
Prevalence = 78.5%
Variability of 24−hour DBP
Eubacteriales sp. HG3A.0242
Prevalence = 53.9%
Variability of 24−hour DBP
Eubacteriales sp. HG3A.0244
Prevalence = 35.6%
Variability of 24−hour DBP
Eubacteriales sp. HG3A.0381
Prevalence = 34.7%
Variability of 24−hour DBP
Eubacteriales sp. HG3A.0383
Prevalence = 25.2%
Variability of 24−hour DBP
Eubacteriales sp. HG3A.0409
Prevalence = 26.1%
Variability of 24−hour DBP
Eubacteriales sp. HG3A.0450
Prevalence = 19.6%
Variability of 24−hour DBP
Eubacteriales sp. HG3A.0636
Prevalence = 11.2%
Variability of 24−hour DBP
Eubacteriales sp. HG3A.0859
Prevalence = 12.9%
(sensitivity analysis estimate − model 2 estimate) / model 2 estimate
Antib
IBD
PPI
P value < 0.05
P value >= 0.05

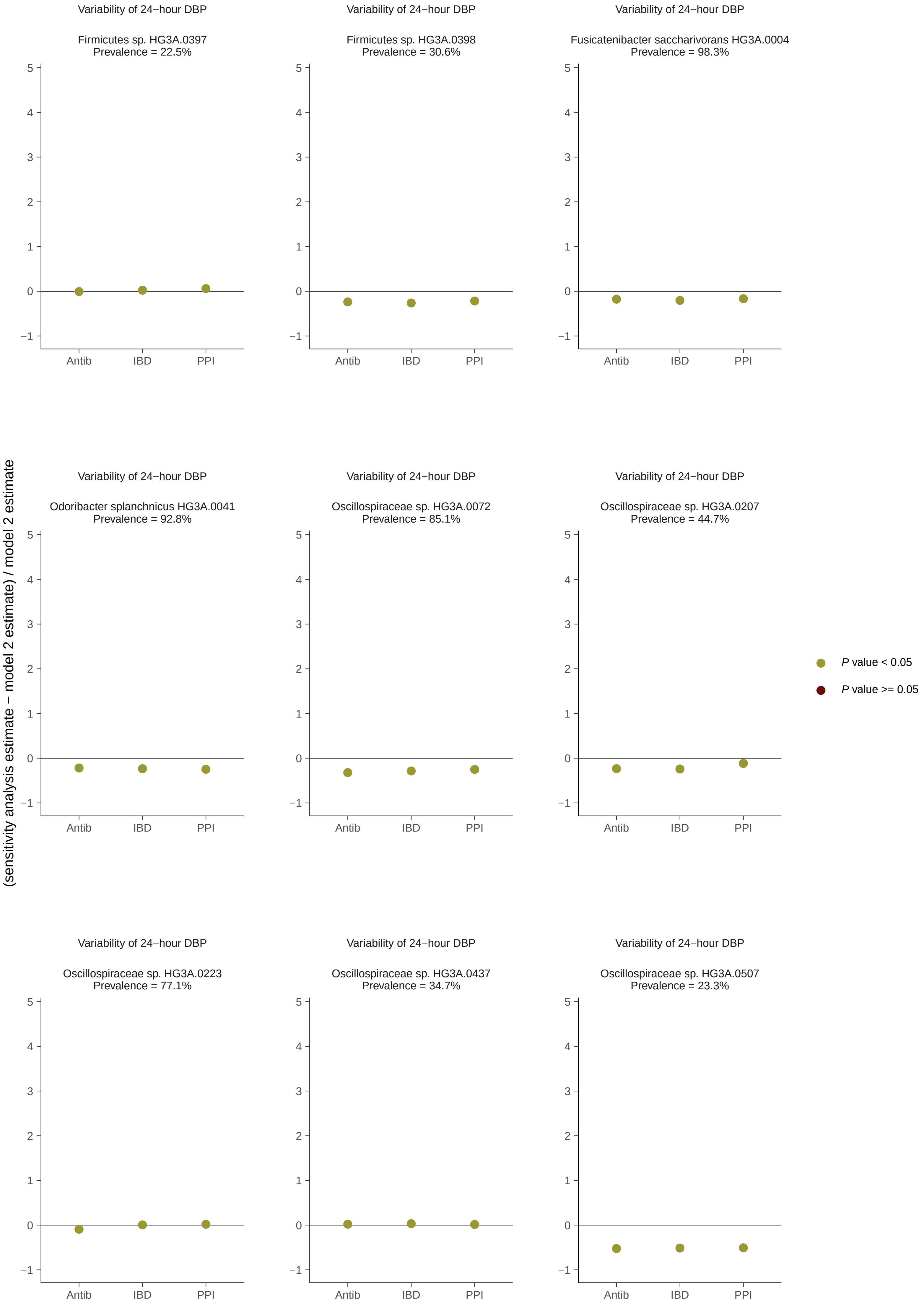
Variability of 24−hour DBP
Firmicutes sp. HG3A.0397
Prevalence = 22.5%
Variability of 24−hour DBP
Firmicutes sp. HG3A.0398
Prevalence = 30.6%
Variability of 24−hour DBP
Fusicatenibacter saccharivorans HG3A.0004
Prevalence = 98.3%
Variability of 24−hour DBP
Odoribacter splanchnicus HG3A.0041
Prevalence = 92.8%
Variability of 24−hour DBP
Oscillospiraceae sp. HG3A.0072
Prevalence = 85.1%
Variability of 24−hour DBP
Oscillospiraceae sp. HG3A.0207
Prevalence = 44.7%
Variability of 24−hour DBP
Oscillospiraceae sp. HG3A.0223
Prevalence = 77.1%
Variability of 24−hour DBP
Oscillospiraceae sp. HG3A.0437
Prevalence = 34.7%
Variability of 24−hour DBP
Oscillospiraceae sp. HG3A.0507
Prevalence = 23.3%
(sensitivity analysis estimate − model 2 estimate) / model 2 estimate
Antib
IBD
PPI
5
4
3
2
1
0
−1
P value < 0.05
P value >= 0.05

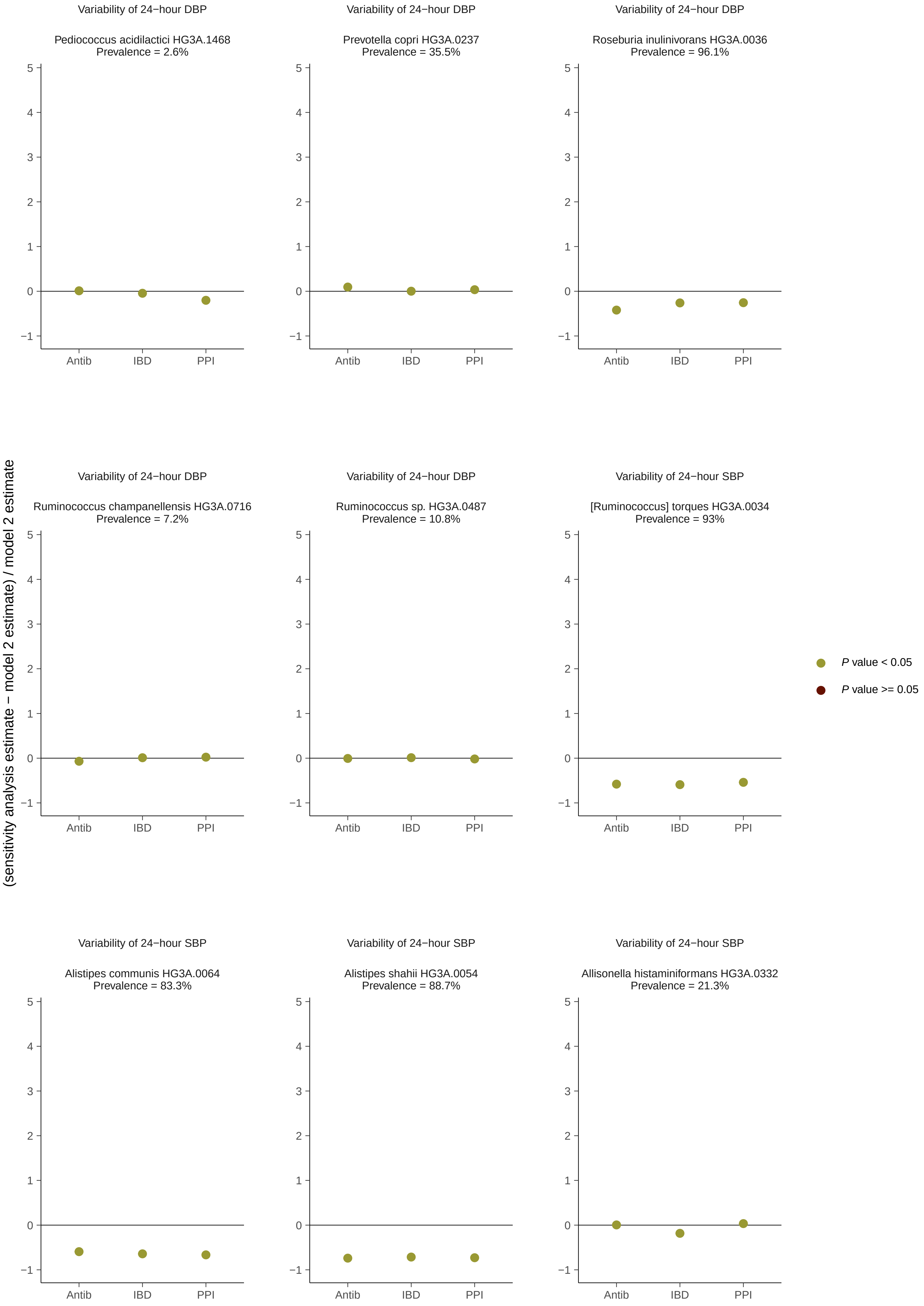
Variability of 24−hour DBP
Pediococcus acidilactici HG3A.1468
Prevalence = 2.6%
Variability of 24−hour DBP
Prevotella copri HG3A.0237
Prevalence = 35.5%
Variability of 24−hour DBP
Roseburia inulinivorans HG3A.0036
Prevalence = 96.1%
Variability of 24−hour DBP
Ruminococcus champanellensis HG3A.0716
Prevalence = 7.2%
Variability of 24−hour DBP
Ruminococcus sp. HG3A.0487
Prevalence = 10.8%
Variability of 24−hour SBP
[Ruminococcus] torques HG3A.0034
Prevalence = 93%
Variability of 24−hour SBP
Alistipes communis HG3A.0064
Prevalence = 83.3%
Variability of 24−hour SBP
Alistipes shahii HG3A.0054
Prevalence = 88.7%
Variability of 24−hour SBP
Allisonella histaminiformans HG3A.0332
Prevalence = 21.3%
Antib
IBD
PPI
(sensitivity analysis estimate − model 2 estimate) / model 2 estimate
P value < 0.05
P value >= 0.05

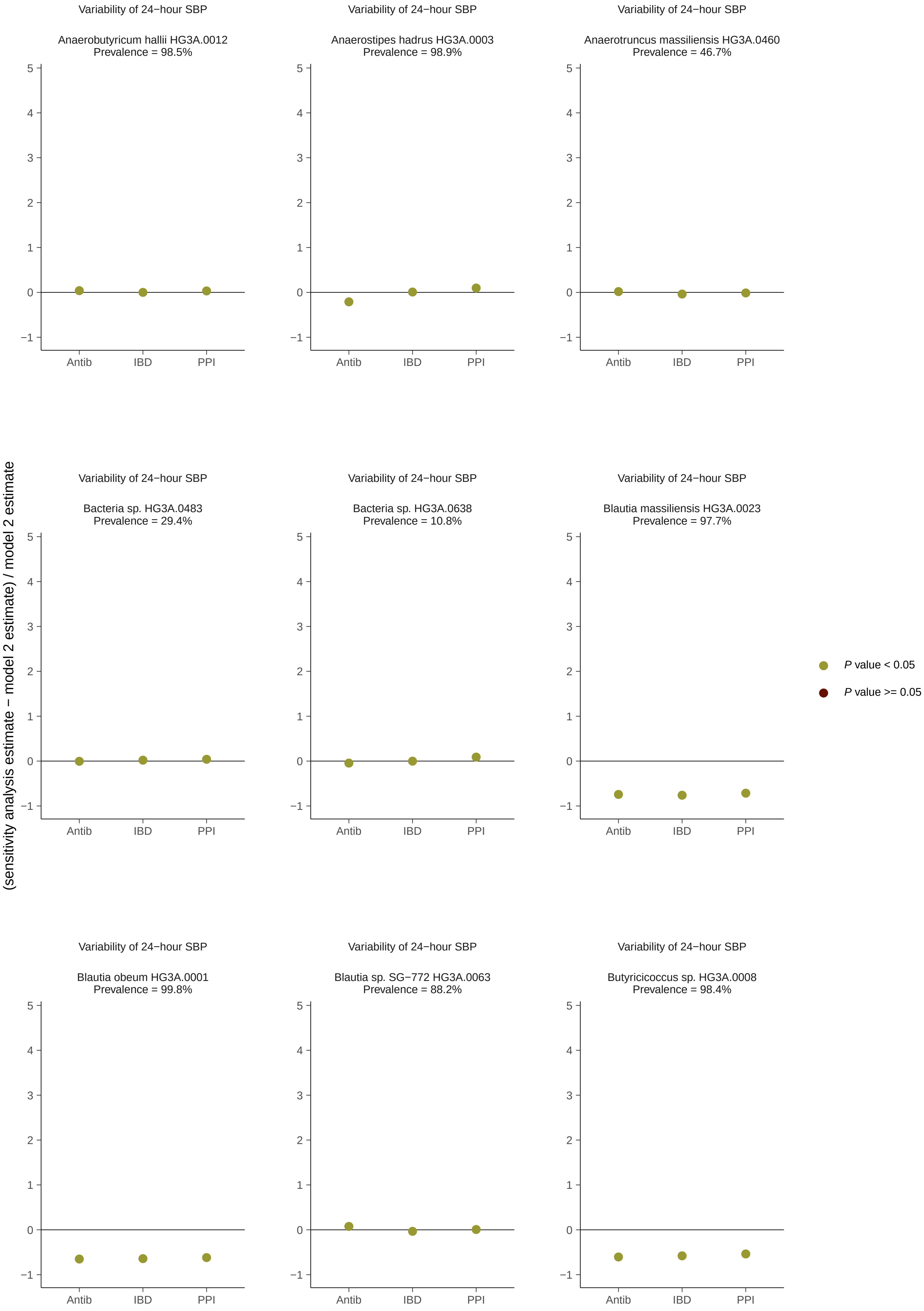
Variability of 24−hour SBP
Anaerobutyricum hallii HG3A.0012
Prevalence = 98.5%
Variability of 24−hour SBP
Anaerostipes hadrus HG3A.0003
Prevalence = 98.9%
Variability of 24−hour SBP
Anaerotruncus massiliensis HG3A.0460
Prevalence = 46.7%
Variability of 24−hour SBP
Bacteria sp. HG3A.0483
Prevalence = 29.4%
Variability of 24−hour SBP
Bacteria sp. HG3A.0638
Prevalence = 10.8%
Variability of 24−hour SBP
Blautia massiliensis HG3A.0023
Prevalence = 97.7%
Variability of 24−hour SBP
Blautia obeum HG3A.0001
Prevalence = 99.8%
Variability of 24−hour SBP
Blautia sp. SG−772 HG3A.0063
Prevalence = 88.2%
Variability of 24−hour SBP
Butyricicoccus sp. HG3A.0008
Prevalence = 98.4%
(sensitivity analysis estimate − model 2 estimate) / model 2 estimate
Antib
IBD
PPI
5
4
3
2
1
0
−1
P value < 0.05
P value >= 0.05

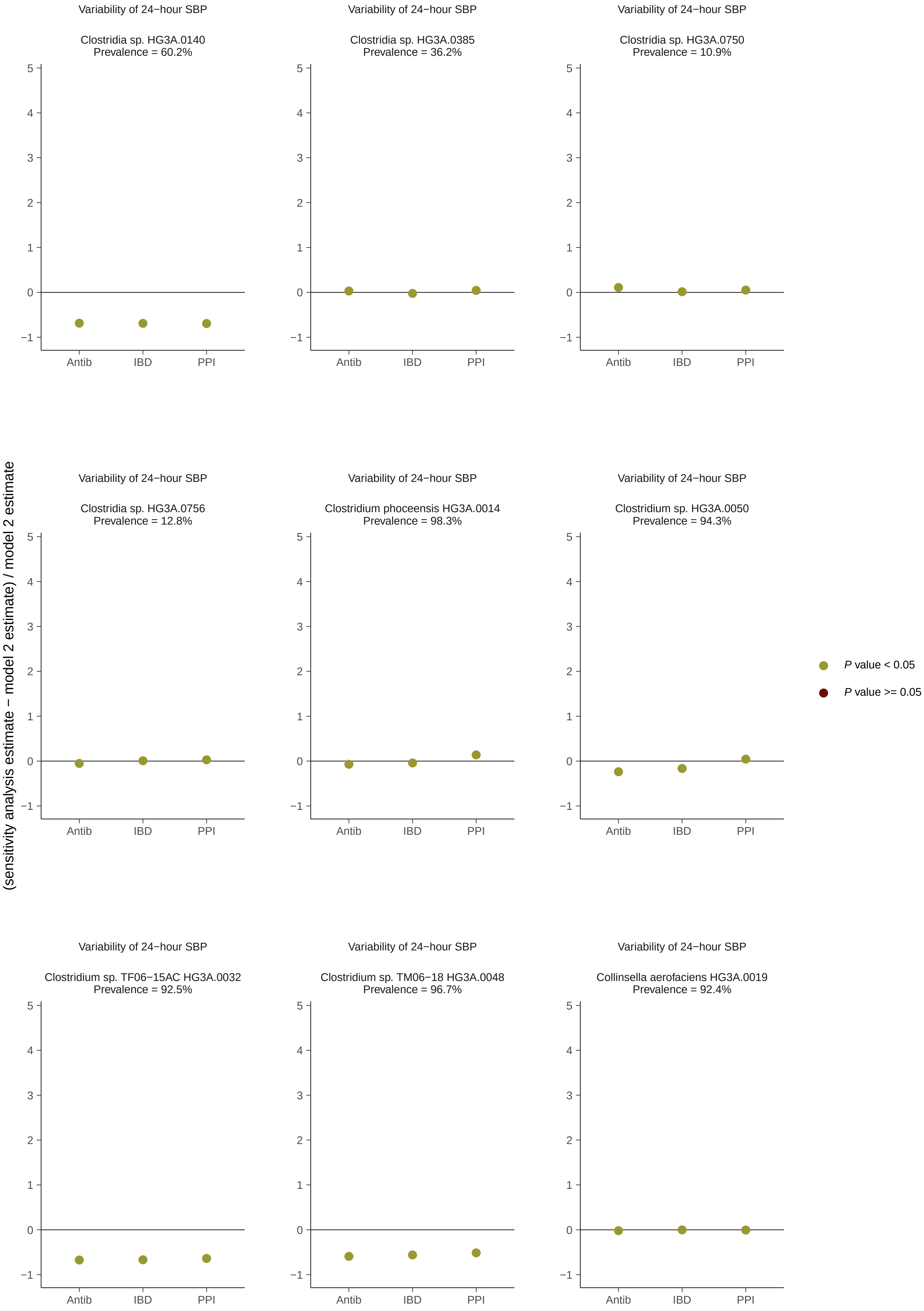

Variability of 24−hour SBP
Clostridia sp. HG3A.0140
Prevalence = 60.2%
Clostridia sp. HG3A.0385
Prevalence = 36.2%
Clostridia sp. HG3A.0750
Prevalence = 10.9%
Clostridia sp. HG3A.0756
Prevalence = 12.8%
Clostridium phoceensis HG3A.0014
Prevalence = 98.3%
Clostridium sp. HG3A.0050
Prevalence = 94.3%
Clostridium sp. TF06−15AC HG3A.0032
Prevalence = 92.5%
Clostridium sp. TM06−18 HG3A.0048
Prevalence = 96.7%
Collinsella aerofaciens HG3A.0019
Prevalence = 92.4%
Antib
IBD
PPI
(sensitivity analysis estimate − model 2 estimate) / model 2 estimate
P value < 0.05
P value >= 0.05

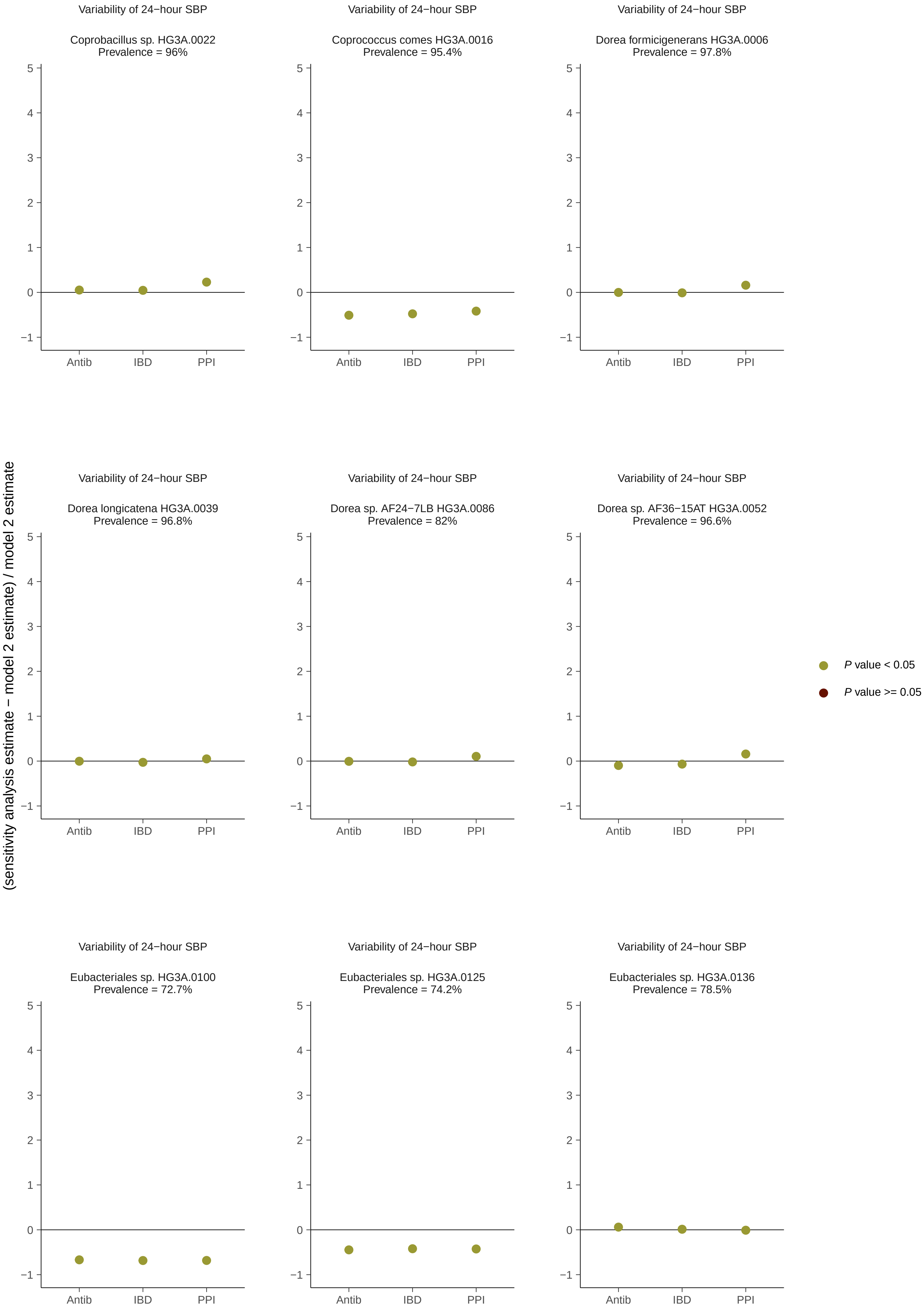

Variability of 24−hour SBP
Coprobacillus sp. HG3A.0022
Prevalence = 96%
Variability of 24−hour SBP
Coprococcus comes HG3A.0016
Prevalence = 95.4%
Variability of 24−hour SBP
Dorea formicigenerans HG3A.0006
Prevalence = 97.8%
Variability of 24−hour SBP
Dorea longicatena HG3A.0039
Prevalence = 96.8%
Variability of 24−hour SBP
Dorea sp. AF24−7LB HG3A.0086
Prevalence = 82%
Variability of 24−hour SBP
Dorea sp. AF36−15AT HG3A.0052
Prevalence = 96.6%
Variability of 24−hour SBP
Eubacteriales sp. HG3A.0100
Prevalence = 72.7%
Variability of 24−hour SBP
Eubacteriales sp. HG3A.0125
Prevalence = 74.2%
Variability of 24−hour SBP
Eubacteriales sp. HG3A.0136
Prevalence = 78.5%
Antib
IBD
PPI
(sensitivity analysis estimate − model 2 estimate) / model 2 estimate
P value < 0.05
P value >= 0.05

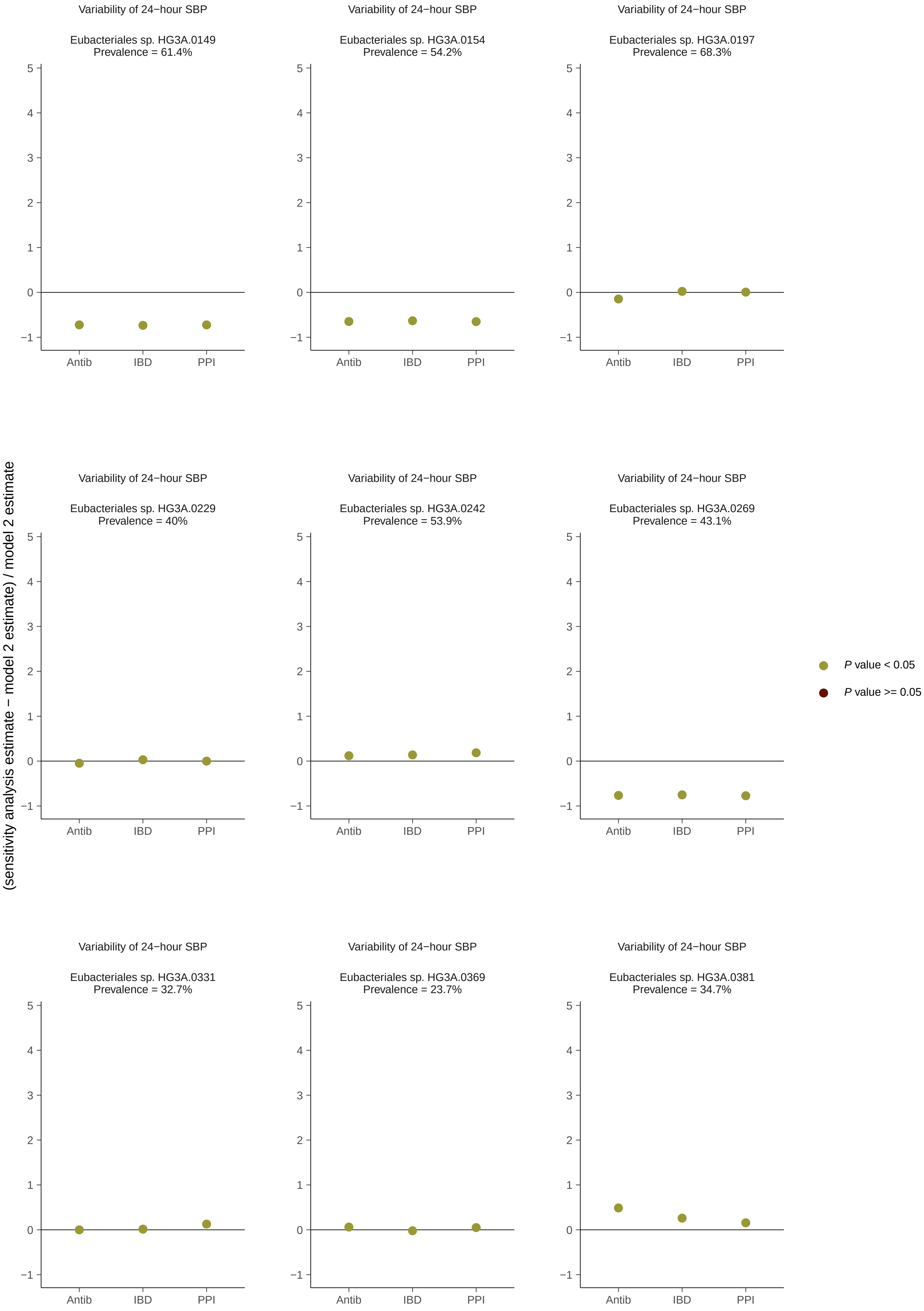

Variability of 24−hour SBP
Eubacteriales sp. HG3A.0149
Prevalence = 61.4%
Eubacteriales sp. HG3A.0154
Prevalence = 54.2%
Eubacteriales sp. HG3A.0197
Prevalence = 68.3%
Eubacteriales sp. HG3A.0229
Prevalence = 40%
Eubacteriales sp. HG3A.0242
Prevalence = 53.9%
Eubacteriales sp. HG3A.0269
Prevalence = 43.1%
Eubacteriales sp. HG3A.0331
Prevalence = 32.7%
Eubacteriales sp. HG3A.0369
Prevalence = 23.7%
Eubacteriales sp. HG3A.0381
Prevalence = 34.7%
Antib
IBD
PPI
(sensitivity analysis estimate − model 2 estimate) / model 2 estimate
P value < 0.05
P value >= 0.05

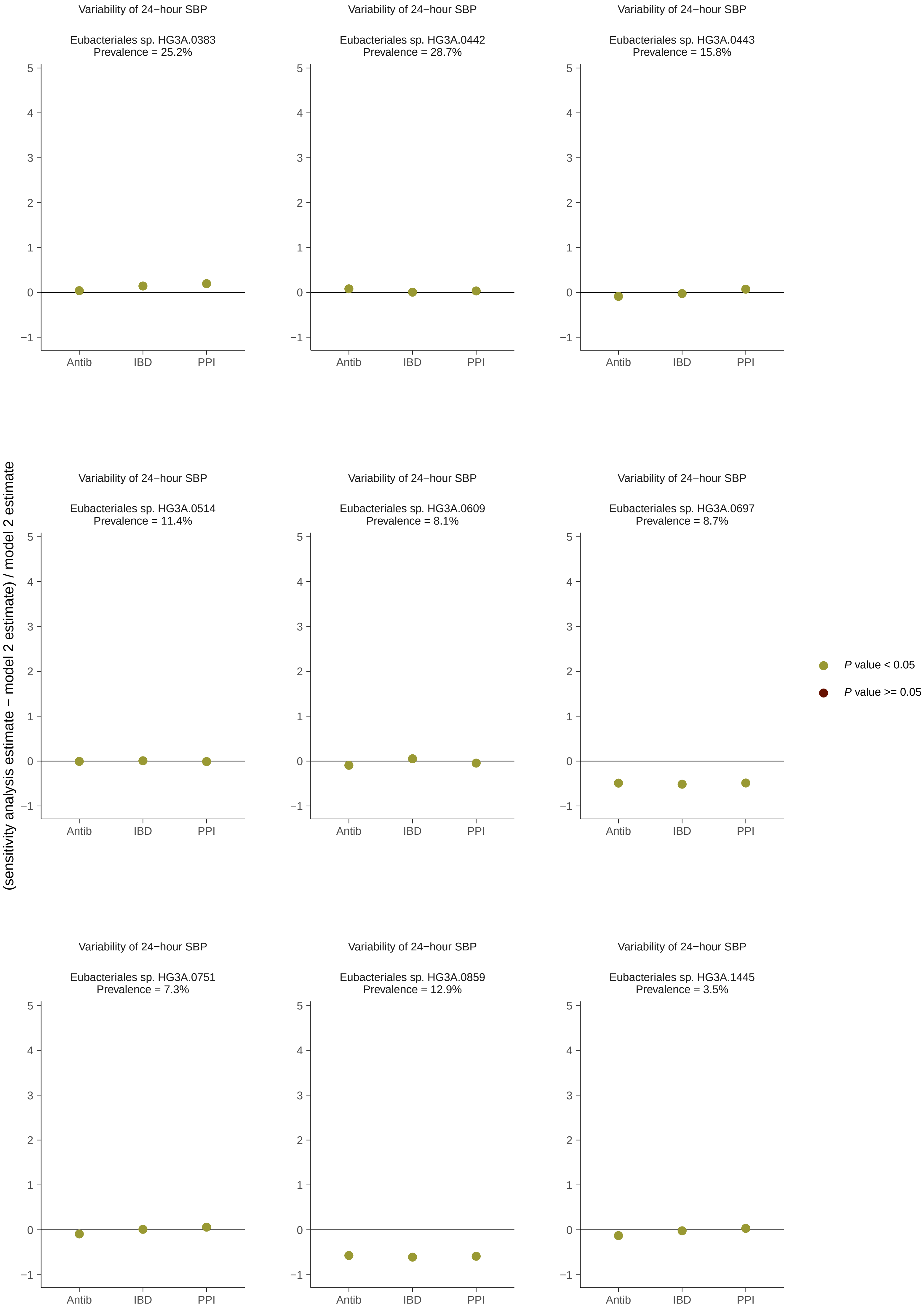

Variability of 24−hour SBP
Eubacteriales sp. HG3A.0383
Prevalence = 25.2%
Variability of 24−hour SBP
Eubacteriales sp. HG3A.0442
Prevalence = 28.7%
Variability of 24−hour SBP
Eubacteriales sp. HG3A.0443
Prevalence = 15.8%
Variability of 24−hour SBP
Eubacteriales sp. HG3A.0514
Prevalence = 11.4%
Variability of 24−hour SBP
Eubacteriales sp. HG3A.0609
Prevalence = 8.1%
Variability of 24−hour SBP
Eubacteriales sp. HG3A.0697
Prevalence = 8.7%
Variability of 24−hour SBP
Eubacteriales sp. HG3A.0751
Prevalence = 7.3%
Variability of 24−hour SBP
Eubacteriales sp. HG3A.0859
Prevalence = 12.9%
Variability of 24−hour SBP
Eubacteriales sp. HG3A.1445
Prevalence = 3.5%
5
4
3
2
1
0
−1
Antib
IBD
PPI
(sensitivity analysis estimate − model 2 estimate) / model 2 estimate
P value < 0.05
P value >= 0.05

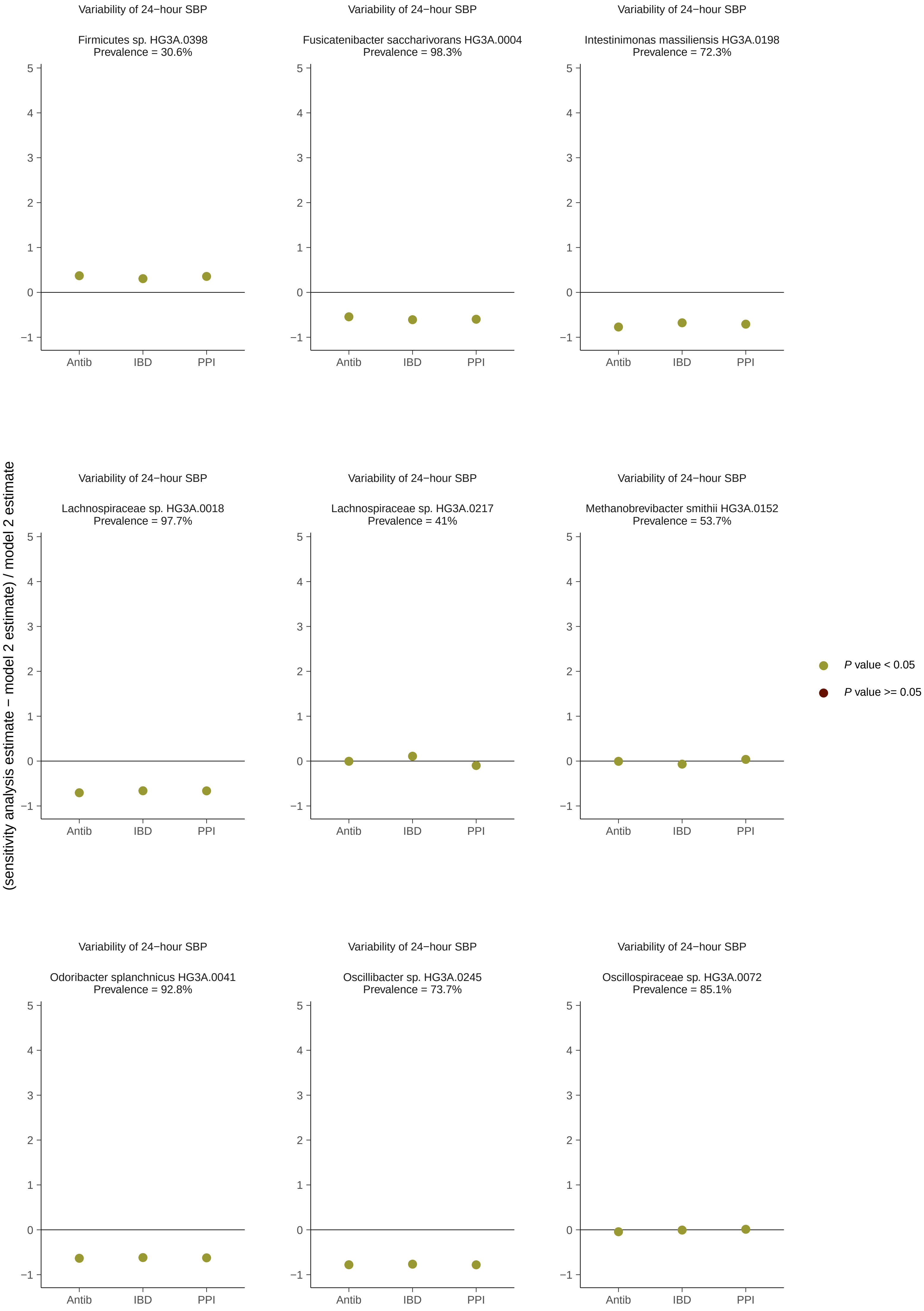
Variability of 24−hour SBP
Firmicutes sp. HG3A.0398
Prevalence = 30.6%
Variability of 24−hour SBP
Fusicatenibacter saccharivorans HG3A.0004
Prevalence = 98.3%
Variability of 24−hour SBP
Intestinimonas massiliensis HG3A.0198
Prevalence = 72.3%
Variability of 24−hour SBP
Lachnospiraceae sp. HG3A.0018
Prevalence = 97.7%
Variability of 24−hour SBP
Lachnospiraceae sp. HG3A.0217
Prevalence = 41%
Variability of 24−hour SBP
Methanobrevibacter smithii HG3A.0152
Prevalence = 53.7%
Variability of 24−hour SBP
Odoribacter splanchnicus HG3A.0041
Prevalence = 92.8%
Variability of 24−hour SBP
Oscillibacter sp. HG3A.0245
Prevalence = 73.7%
Variability of 24−hour SBP
Oscillospiraceae sp. HG3A.0072
Prevalence = 85.1%
Antib
IBD
PPI
(sensitivity analysis estimate − model 2 estimate) / model 2 estimate
P value < 0.05
P value >= 0.05

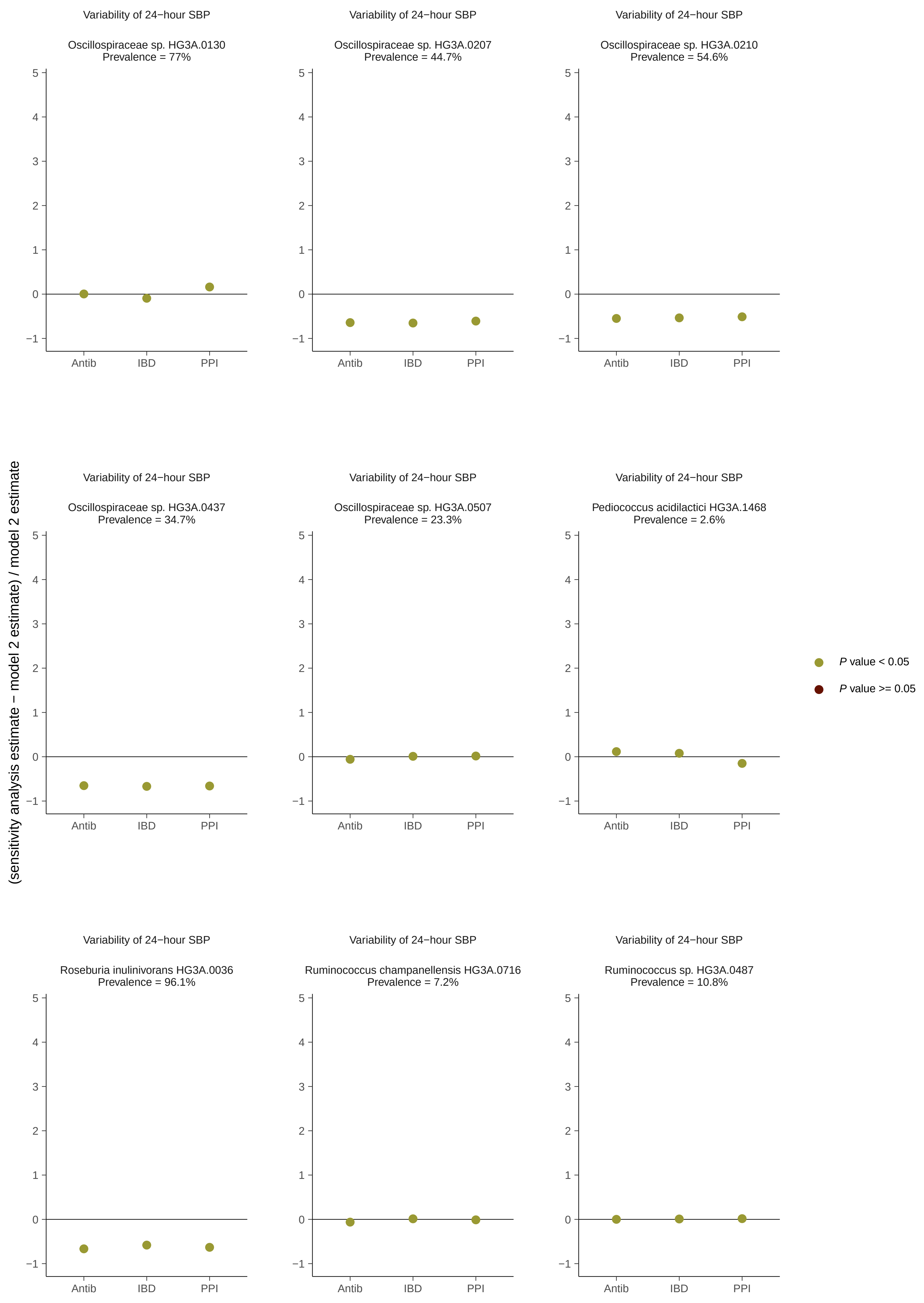

(sensitivity analysis estimate − model 2 estimate) / model 2 estimate
Variability of 24−hour SBP
Oscillospiraceae sp. HG3A.0130
Prevalence = 77%
Variability of 24−hour SBP
Oscillospiraceae sp. HG3A.0207
Prevalence = 44.7%
Variability of 24−hour SBP
Oscillospiraceae sp. HG3A.0210
Prevalence = 54.6%
Variability of 24−hour SBP
Oscillospiraceae sp. HG3A.0437
Prevalence = 34.7%
Variability of 24−hour SBP
Oscillospiraceae sp. HG3A.0507
Prevalence = 23.3%
Variability of 24−hour SBP
Pediococcus acidilactici HG3A.1468
Prevalence = 2.6%
Variability of 24−hour SBP
Roseburia inulinivorans HG3A.0036
Prevalence = 96.1%
Variability of 24−hour SBP
Ruminococcus champanellensis HG3A.0716
Prevalence = 7.2%
Variability of 24−hour SBP
Ruminococcus sp. HG3A.0487
Prevalence = 10.8%
5
4
3
2
1
0
−1
Antib
IBD
PPI
P value < 0.05
P value >= 0.05

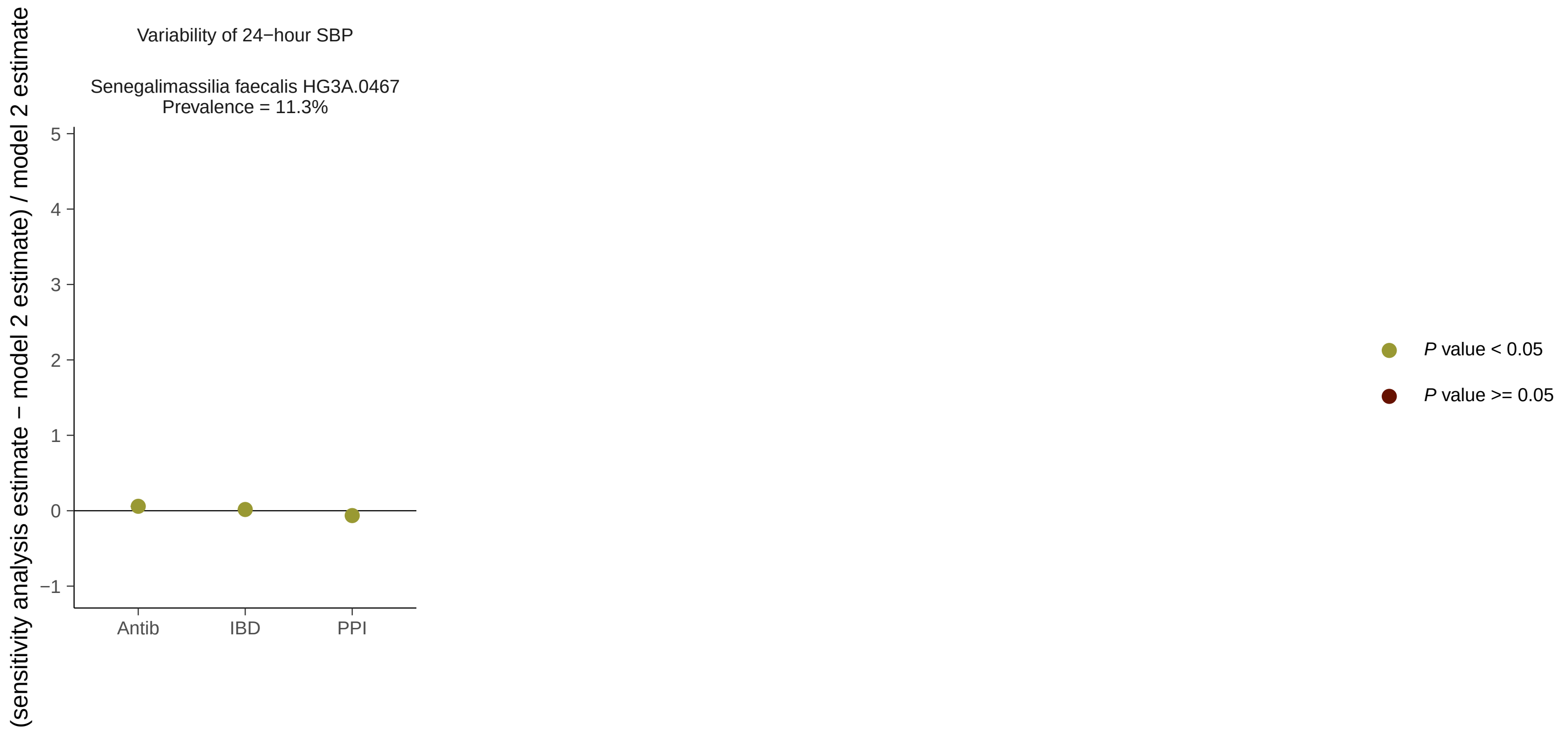

Variability of 24−hour SBP
Senegalimassilia faecalis HG3A.0467
Prevalence = 11.3%
(sensitivity analysis estimate − model 2 estimate) / model 2 estimate
5
4
3
2
1
0
−1
Antib
IBD
PPI
P value < 0.05
P value >= 0.05
